# Genomic and biological panoramas of non-muscle actinopathies

**DOI:** 10.1101/2024.08.21.24310320

**Authors:** Nataliya Di Donato, NMA Consortium, Andrew Thom, Andreas Rump, Johannes N. Greve, Marcus Kropp, Juan Cadiñanos, Salvatore Calabro, Sara Cathey, Brian Chung, Heidi Cope, Maria Costales, Sara Cuvertino, Philine Dinkel, Kalliopi Erripi, Andrew E. Fry, Livia Garavelli, Kaomei Guan, Sabine Hoffjan, Wibke G. Janzarik, Matti Koenig, Insa Kreimer, Karolina Kuenzel, Grazia Mancini, Purificacion Marin-Reina, Andrea Meinhardt, Indra Niehaus, Daniela Pilz, Ivana Ricca, Fernando Santos Simarro, Evelin Schrock, Anja Marquardt, Manuel H. Taft, Kamer Tezcan, Sofia Thunström, Judith Verhagen, Alain Verloes, Bernd Wollnik, Peter Krawitz, Tzung-Chien Hsieh, Leo Zeef, Michael Seifert, Michael Heide, Catherine B. Lawrence, Neil Roberts, Dietmar Manstein, Adrian S. Woolf, Siddharth Banka

## Abstract

**Background:** Cytoskeletal non-muscle actin isoforms are the most abundant intracellular proteins and extensively interact with other molecules. Biological consequences and genotype-phenotype correlations of the variants in genes encoding these isoforms, ACTB and ACTG1, are not delineated.

**Methods:** Clinical data analysis from 290 individuals with pathogenic ACTB/ACTG1 variants; characterization of patient cells, mutant proteins, patient-derived iPSC-based models and mutant mice.

**Results:** We show that ACTB and ACTG1 variants have distinct clinical profiles. ACTB nonsense, frameshift and missense variants that lead to rapid protein degradation result in milder phenotypes. Heterozygous Actb knockout causes altered neuronal cell morphology and abnormal expression of actin-related genes in newborn mouse brains. Truncating ACTG1 variants are likely to be non-pathogenic, but chromosomal deletions encompassing ACTG1 and flanking genes may result in susceptibility to neurodevelopmental phenotypes. Subsets of disease-causing ACTB missense variants (MVs) result in more severe Type 1 Baraitser-Winter Cerebrofrontofacial (BWCFF1) or Deafness Dystonia syndromes. Pathogenic ACTG1 MVs cause BWCFF2 or isolated hearing loss. These amino acid substitutions are associated with dramatically dysregulated actin polymerization and depolymerization dynamics and, in induced pluripotent stem cells (iPSC) derived models, lead to neuronal migration defects. A significant subset of MVs result in disorders that cannot yet be classified into recognizable groups.

**Conclusions:** ACTB or ACTG1 variants and result in minimum eight mechanistically diverse non-muscle actinopathies. These results will improve their diagnosis and management, and pave the way for new treatment strategies. This study reflects the scale of collaborative clinical studies and multi-modal mechanistic studies required to dissect rare allelic disorders.

## Introduction

The ability to make accurate diagnoses, reliably estimate prognoses, and having a deep understanding of underpinning molecular and disease mechanisms are prerequisites to developing precision medicine approaches for genetic disorders. Recent advances in Mendelian genomics has dismantled the ‘one-gene one-disorder paradigm’(1). This means that many genetic disorders will likely require characterisation at the variants level, and not just at gene level. We attempted dissection of a complex set of allelic disorders caused by constitutional variants in two genes encoding for actin isoforms which we here refer to collectively as non-muscle actinopathies (NMAs).

Actins are the most abundant intracellular proteins, highly versatile and play crucial roles in multiple cellular processes. The mammalian actin gene family consists of four muscle-specific isoforms (*ACTA1*, *ACTA2*, *ACTC1*, and *ACTG2*) and two ubiquitously expressed genes, *ACTB* and *ACTG1*, respectively coding for the highly conserved b- and g-cytoplasmic actins (CYAs). Human bCYA and gCYA differ only in four out of 375 amino acids, but possess different polymerisation properties, localize in different parts of the cell, and display preferred interactions with different subsets of actin binding proteins(2).

NMAs listed in OMIM include *ACTB*-related Baraitser-Winter cerebrofrontofacial syndrome (BWCFF) 1 (#243310), Dystonia-deafness syndrome 1 (DDS1) (#607371)(3), Thrombocytopenia 8 with dysmorphic features and developmental delay (#620475)(4), *ACTG1*-related BWCFF2 (#614583)(5, 6) and dominant deafness 20/26 (#604717)(7, 8). An additional documented NMA, *ACTB*-associated isolated ocular coloboma(9), is not listed in OMIM. A neurodevelopmental-congenital malformation disorder due to loss-of-function *ACTB* variants(10) is included within BWCFF1. Furthermore, postzygotic somatic mosaic *ACTB* variants have been detected in Becker’s nevus, segmental odontomaxillary dysplasia, and congenital smooth muscle hamartoma with or without hemihypertrophy(11–13). Nevertheless, the full mutational and clinical spectrum of human monogenic disorders caused by *ACTB* or *ACTG1* variants has yet to be systematically explored. Challenges in assigning pathogenicity and in linking individual variants to specific NMAs hamper accurate diagnosis. Even after a specific NMA is diagnosed, clinical management and providing prognosis is challenging due to limited data. Moreover, the pathobiology of NMAs is poorly understood. For example, it is not known whether disease-causing missense variants (MVs) in NMAs are hypomorphic or dominant-negative or gain-of-function. Because of these limitations, no specific treatments for these disorders exist.

Here, we undertake detailed studies of disease-causing variants in *ACTB* and *ACTG1*. We describe a cohort of 290 individuals with pathogenic or likely pathogenic (P/LP) variants and propose a variant- and phenotype-based classification system that delineates NMAs and defines their clinical spectrum. Using *in vitro* cellular and biochemical studies, we provide variant level insights into the molecular mechanisms of NMAs. Finally, using a mutant mouse, a mouse neuronal cell line, and patient-derived induced pluripotent stem cell models, we provide insights into the pathogenesis of neural defects in NMAs.

## Results

### *ACTB* and *ACTG1* variants have dissimilar population profiles

To study the profile and consequences of the variants in the NMA-genes, we simulated all possible *ACTB* and *ACTG1* single nucleotide substitutions and compared them with known variants from population databases, representing ~431,130 individuals(14–16). Simulated variant counts per gene were overall similar, but we observed several differences between the non-synonymous, synonymous and non-coding population variant counts of the two genes, which are unlikely to be the result of differences in mutation acquisition potential **(Suppl. Fig. S1-2, Suppl. Table 1)**. For example, the population dataset included only one individual with pLOF (predicted loss-of-function) *ACTB* variant (pLI=0.99), but >20 individuals with truncating *ACTG1* variants (pLI=0)(17) **(Fig. 1A, Suppl. Fig. S1A)**. Both genes are highly constrained for population MVs (Suppl. Table S1A), but *ACTB* is more intolerant (43 distinct MVs, 0.02% population frequency) than *ACTG1* (149 MVs, 0.05% population frequency) **(Suppl. Table S1)**(17).

**Fig. 1.**
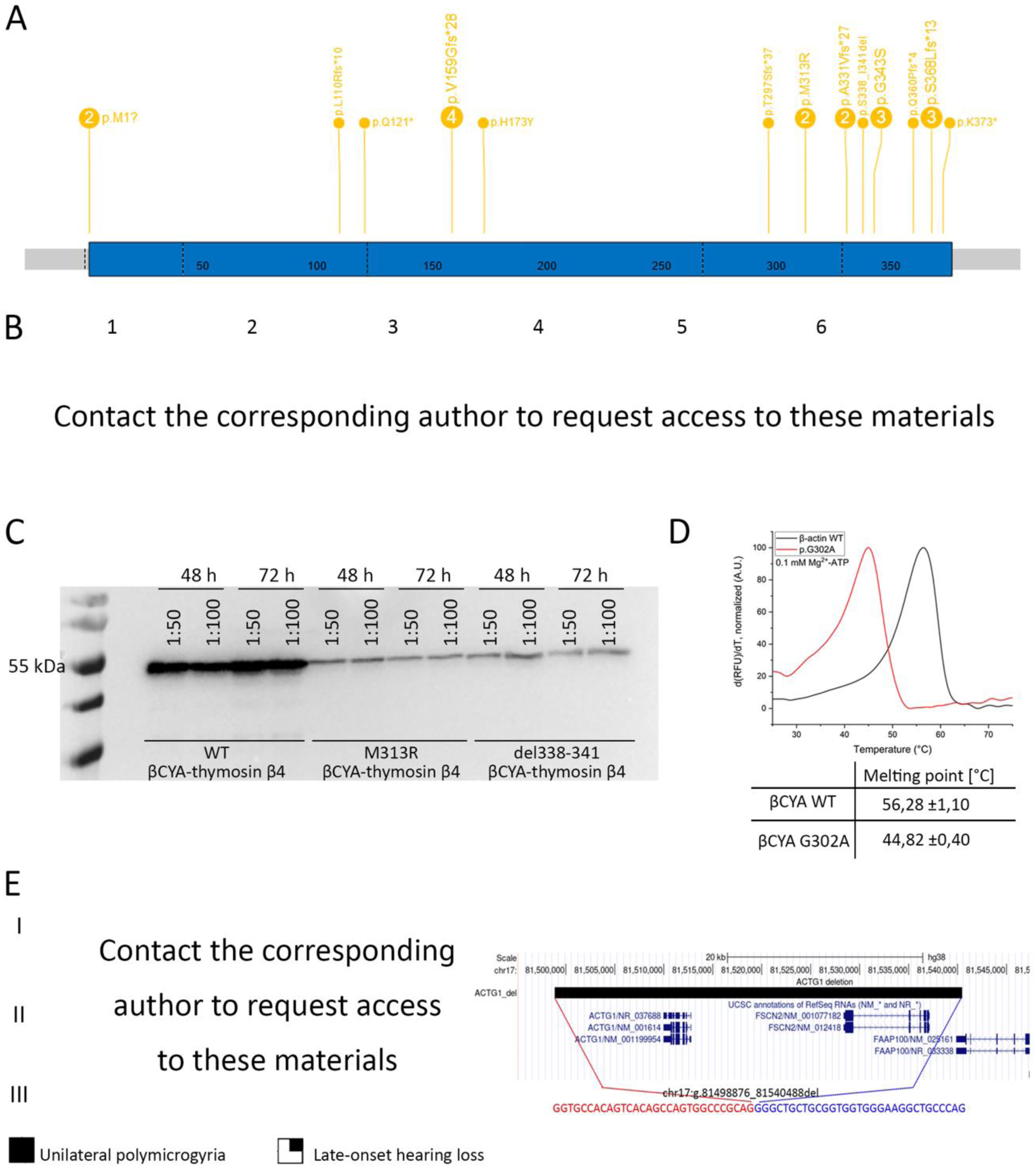
Definition of the functional haploinsufficiency of *ACTB* and *ACTG1*. **(A)** Number and distribution of truncating variants (start loss, stop gain, frameshift) as well as missense variants in *ACTB* in patients with identical phenotype **(B)** Facial gestalt in patients with the *ACTB* whole gene deletions (1,2), *ACTB* missense variant (3), and familial presentation of the *ACTB* stop gain (4–6) **(C)** Immunoblot of Sf9 insect cell lysate revealing only small amounts of mutated actin-thymosin β4 fusion constructs in the cell lysate. This amount was not sufficient to purify mutant constructs as they could not be eluted from NiNTA column. Cells were transfected with different titres of baculovirus encoding for the respective actin-thymosin β4 construct (1:50, 1:100). Samples were taken 48 hours and 72 hours after transfection. Blot was developed using the anti-Penta-His antibody (Qiagen, Hilden, Germany) and the goat anti-mouse IgG-HRP secondary antibody (Thermo, Waltham, USA) **(D)** Thermofluor-assay monitoring the thermal stability of Mg^2+^-ATP-G-actin. Representative traces of melting experiments with β-actin WT and G302A in the presence of 0.1 µM Mg^2+^-ATP. Pictured are the first derivatives of the melting curves. The melting point is determined from the peak of the function. Final concentration of actin is 0.2 mg/mL. 5 independent experiments were performed for β-actin WT and G302A. Means and SE are shown in the table below the graph. **(E)** Segregation and intrafamilial variability of microdeletion of *ACTG1* with the pedigree with the affected relatives (II.4 and III.4) with unilateral polymicrogyria on the left; +/− indicates the carrier status; and the UCSC genome hg38 browser view demonstrating a microdeletion encompassing *ACTG1* and *FSCB2*, coordinates of the last normal probes are shown in red with the precise breakpoint mapping in blue (first and last missing bases) including junction sequence shown below.

Next, we compiled clinical information from 290 individuals (125 new) with *ACTB* or *ACTG1* variants classified as P/LP(18). This ‘NMA cohort’ comprised 275 individuals with SNVs (single nucleotide variants) in *ACTB* (N=145) or *ACTG1* (N=130) and 15 individuals with small deletions encompassing *ACTB* or *ACTG1* **(Suppl. Table S2)**. The cohort’s age range spanned 20 weeks gestation to 60 years; 54% were males and 46% females. The phenotype profiles of individuals in the NMA cohort, as expected, were highly variable.

*ACTB* and *ACTG1* variants, therefore, have remarkably different population profiles. These observations support their different biological roles in humans. Our initial examination of the clinical data revealed impressive heterogeneity, indicating the need for systematic in-depth analysis.

### *ACTB* and *ACTG1* loss of function alleles have distinct clinical consequences

To study the clinical consequences of variations in the two genes, we devised a variant and phenotypic-led approach **(Suppl. Fig. S3)**. We first analysed pLOF variants in our NMA cohort. Previously, two conditions have been described to result from *ACTB* pLOF variants i.e. a ‘pleiotropic developmental disorder’ due to 7p22.1 chromosomal microdeletions or point variants(10), and syndromic thrombocytopenia caused by point variants in the last exon(4). We compared the phenotypes of individuals with either microdeletions encompassing *ACTB* or with individuals *ACTB* pLOF variants expected to undergo or escape nonsense mediated decay **(Fig. 1B, Suppl. Table S2)**. The phenotypes of individuals in these three groups were highly overlapping and characterised by mild/moderate neurodevelopmental delay or borderline/mild intellectual difficulties (ID), behaviour anomalies, mild microcephaly, short stature, brain anomalies (including heterotopias, but not pachygyria), and combinations of other congenital malformations and variable thrombocytopenia **(Fig. 1B, Suppl. Note S1)**.

Next, we hypothesised that some *ACTB* MVs may result in simple loss of function. Utilising our clinical delineation of the ‘*ACTB* pLOF disorder’, we identified eight individuals with five *ACTB* MVs/inframe indels and features similar to those of individuals with *ACTB* pLOF variants **(Fig. 1B–3)**. None of these MVs are predicted to significantly alter *ACTB* splicing. Therefore, we generated recombinant mutant bCYA proteins for *in-vitro* characterization of a selection of these variants. Protein purification could not be achieved for bCYA with two of these variants (p.Met313Arg or p.Ser338_Ile341del) indicating a possible defect with protein folding and/or stability **(Fig. 1C)**. For one variant (p.Gly302Ala) the stability of bCYA was significantly reduced **(Fig. 1D)**. We did not test two variants (p.His173Tyr and p.Gly343Ser). In contrast, the NMA cohort did not have any P/LP *ACTG1* pLOF point variants. However, we identified seven individuals with chromosome 17q25 deletions shorter than 1 Mb that encompassed *ACTG1*. Deletion carriers showed variable phenotypes ranging from normal intelligence to borderline intellectual disability (ID) with additional structural anomalies **(Suppl. Table 2 – request from corresponding author)**. This included a 41.6kb deletion including *ACTG1* and *FSCN2* segregating through three generations in mildly affected and healthy relatives **(Fig. 1E)**. *FSCN2* encodes the actin-bundling protein fascin-2 that crosslinks actin filaments into bundles within dynamic membrane protrusions and its pLI score is 0, indicating that its haploinsufficiency alone is unlikely to cause a monogenic disorder(14).

These data indicate that the *ACTB*-related ‘pleiotropic developmental disorder’(10) is distinct from BWCFF1 **(see next section)** and that the ‘syndromic thrombocytopenia syndrome’(4) is the same as the *ACTB*-related ‘pleiotropic developmental disorder. This condition also includes some missense/in-frame *ACTB* variants that lead to translation of unstable CYA and have clinical consequences that overlap with those of pLoF *ACTB* variants. Importantly, *ACTG1* pLOF point variants are unlikely to be pathogenic. Chromosome 17q25.3 deletions involving *ACTG1* and flanking genes could cause a contiguous genes deletion syndrome with variable penetrance, which requires further studies.

### *ACTB* or *ACTG1* missense and inframe variants result in multiple NMAs

Next, applying our classification strategy **(Suppl. Fig. S3)** on individuals with missense or inframe variants in the NMA cohort showed that 73 patients could be categorised into *ACTB-*BWCFF1, 60 with *ACTG1-*isolated hearing loss (HL), 40 with *ACTG1-*BWCFF2 and 13 with *ACTB-*DDS1 **(Fig. 2A-B)**. Detailed inspection of the clinical data of the remaining 65 patients revealed heterogeneous phenotypes not compatible with any of the previously mentioned actinopathies and absence of recognizable facial gestalt (grouped as unspecified non-muscle actinopathy or unNMA) **(Fig. 2D)**. This classification revealed detailed clinical characteristics and variant spectrum for each NMA sub-type **(Suppl. notes S1-5)**. Apart from rare exceptions, we found that individuals with the same variants generally had highly overlapping clinical features and were classified into the same NMA sub-type. Notably, however, we detected substantial differences in severity within the same sub-type. We found the variant spectra to be much larger than previously recognised for all NMAs, apart from *ACTB-*DDS1, which seems to be caused only by *ACTB*:p.Arg183Trp. Interestingly, we observed that for certain positions, the same substitution in *ACTB* or in *ACTG1* can cause different NMAs.

**Fig. 2.**
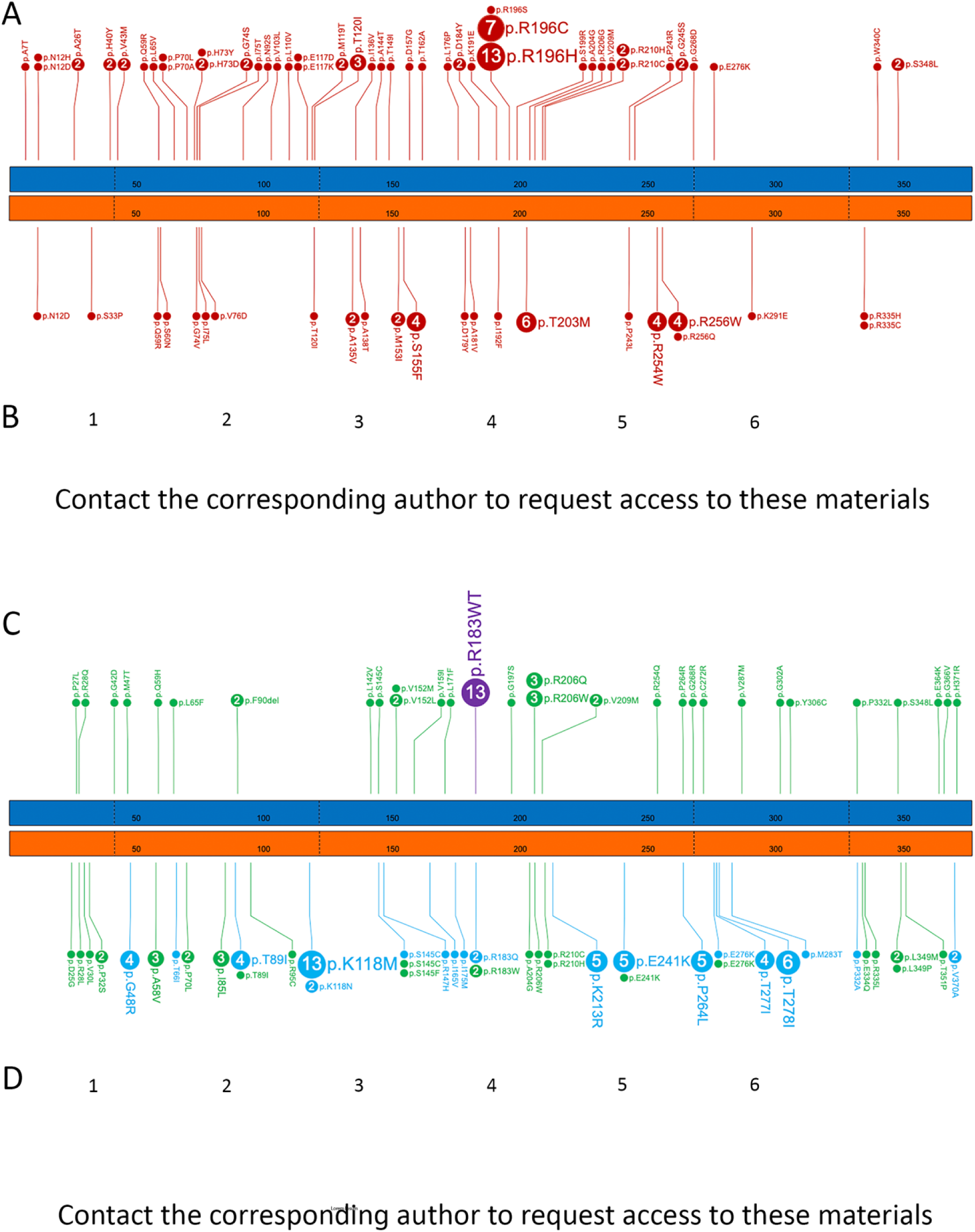
Missense variants in *ACTB* and *ACTG1* result in several distinct disorders. **(A)** Number and distribution of missense variants in *ACTB* and *ACTG1* resulting in the Baraitser-Winter cerebrofrontofacial syndrome (BWCFF); *ACTB* (blue bar) and *ACTG1* (orange bar) with every 50^th^ amino acid numbered in both gene models, the borders of coding exons are marked with dashed lines. **(B)** Representative facial gestalt in patients with the BWCFF syndrome, note that patients on the images 5 and 6 carry the same *ACTB* hotspot variant at the position Arg196 **(C)** Number, distribution, and clinical consequence of the missense variants in *ACTB* and *ACTG1* not resulting in BWC, gene models and amino acid numbering as described above **(D)** Representative facial gestalt in patients with unspecified Non-muscle actinopathies (unNMA) due to variants in *ACTG1* (1–4) and *ACTB* (5,6); note that images 3 and 4 show affected individuals from the same family.

These data show that *ACTB* or *ACTG1* protein altering variants (PAVs) result in multiple NMAs. The consistency of variant specific genotype-phenotype correlations suggests that in most cases the variant, rather than other factors such as genetic background, specifies the resulting NMA sub-type. Importantly, the broad spread of PAVs across all of the NMAs suggests that these variants (with a possible exception of *ACTB*:p.Arg183Trp) are unlikely to be gain-of-function(5). The differences of phenotypes of *ACTB*/*ACTG1* PAVs versus LoF variants indicate that these missense/in-frame variants are unlikely to result in simple loss of function. Hence, most of these variants possibly act in dominant negative manner or result in loss of different sub-functions(19). The heterogeneity of associated clinical features suggests that their cellular consequences could be context-dependent.

### Genetic and clinically led classification of NMAs is accurate and has important clinical implications

To test our clinically led classification **(Fig. 3A)** we subjected our conclusions to objective validation. Firstly, objective analysis of the craniofacial features using the photographs available to us using GestaltMatcher(20, 21) confirmed our classification orthogonally **(Fig. 3B, Suppl. Fig. S4)**. Next, we compared the clinical features of all the NMAs **(Fig. 3C)** and mapped the summarised trends in the severity of intellectual/neurological and the number of congenital anomalies or affected organs **(Fig. 3D)**. This revealed an extraordinarily broad clinical spectrum, ranging from individuals who appear to be unaffected (e.g. small chromosomal deletions across *ACTG1*) or relatively mildly affected (e.g. *ACTB* pLOF variants) to individuals who do not survive till birth.

**Fig. 3.**
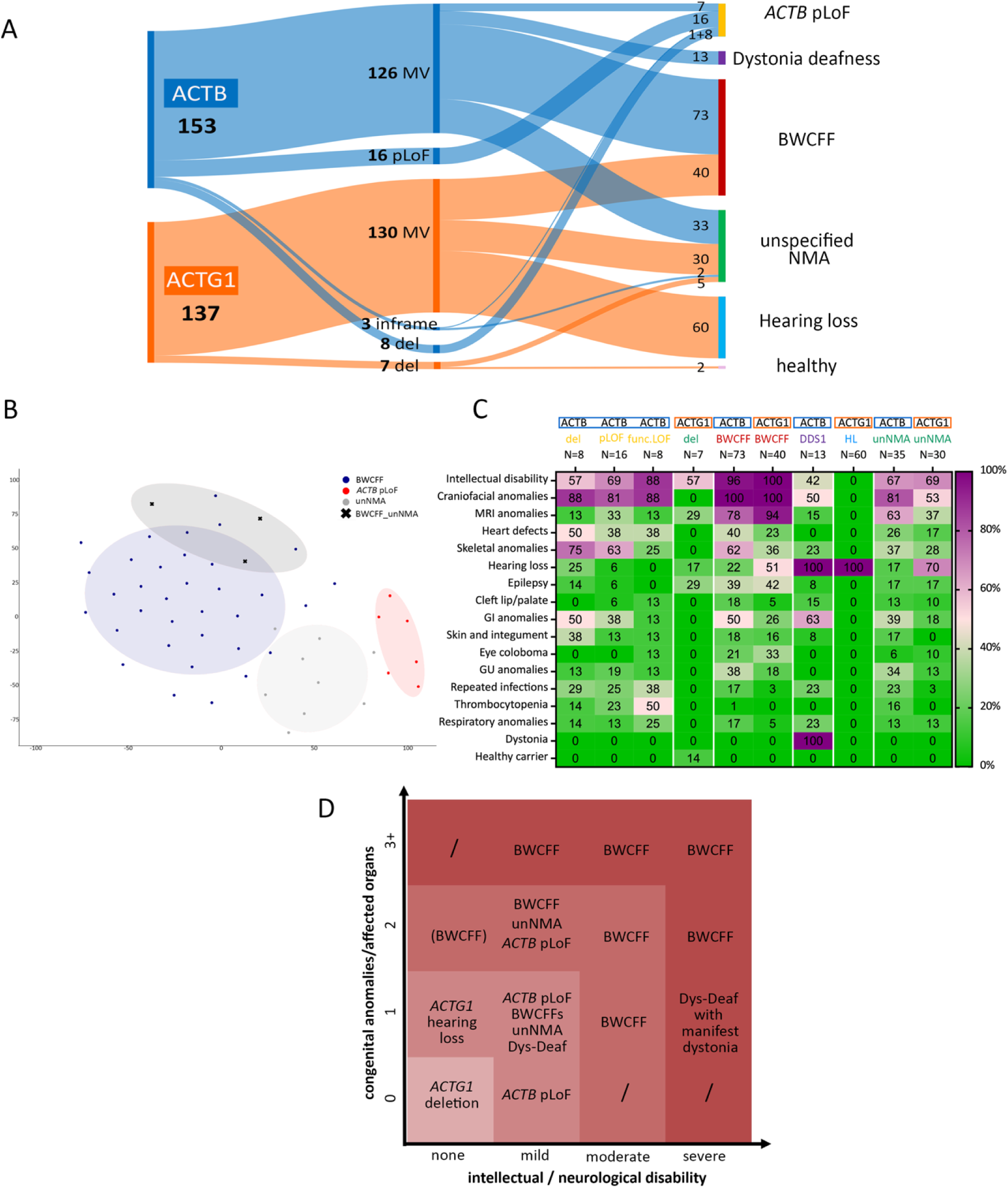
Broad clinical spectrum of Non-muscle actinopathies. **(A)** Distribution of 290 pathogenic variants in *ACTB* and *ACTG1* within different disorders of NMA spectrum; BWCFF Baraitser-Winter-Cerebrofrontofacial syndrome, del whole gene deletion, MV missense variant, pLoF putative loss-of-function **(B)** tSNE visualization of three major NMA phenotypes in GestaltMatcher analysis; NMA-BWCFF indicates three patients that were not reliably classified as BWCFF or unNMA based on the facial gestalt, note that GestaltMatcher analysis allocated all three patients within the BWCFF group **(C)** Phenogrid demonstrating frequencies of selected clinical features in patients with pLoF in *ACTB*, microdeletions encompassing *ACTB* (*ACTB* del) or *ACTG1* (*ACTG1* del) as well as non-truncating *ACTB* variants resulting in unstable protein (*ACTB* func.LoF), BWCFF, dystonia-deafness syndrome (DD); hearing loss (HL) and unspecified non-muscle actinopathy (unNMA); patients with the incomplete clinical information were counted as clinical feature was not present except for GI anomalies, for GI anomalies patients with unknown status were excluded from the calculation; the presence of dystonia was analysed in patients older than 18 years **(D)** Severity grading of the disorders within the non-muscle actinopathy spectrum based on the number of congenital anomalies and/or affected organs and the severity of the intellectual impairment.

These results show that our genetic and clinically led classification of NMAs is reasonably accurate, and that the clinical differences between NMAs have implications for their accurate diagnosis, management, prognosis, and surveillance. Potentially, the grading of severity also opens up new approaches for treatment. For example, since *ACTG1* haploinsufficiency is likely to be benign, gene silencing aimed at inactivating the variant allele could be used to treat some features of *ACTG1*-BWCFF2. Similarly, some features of *ACTB* MV-associated severe NMAs could be attenuated into an “*ACTB* pLOF phenotype”.

### Actin polymerization and depolymerization dynamics are significantly altered in BWCFF syndrome variants

Next, we set out to understand the mechanistic basis of NMAs. Using dermal fibroblasts from individuals with *ACTB*-pLOF disorder, BWCFF1/2, DDS1, and unNMA, we performed transcriptome analysis and immunoblotting to determine amounts of bCYA, gCYA and total actin. No significant differences were found between these groups **(Fig. 4A, Suppl. Fig. S5-S7)**. Examination of actin cytoskeletal patterns within fibroblast with pLOF or BWCFF variants revealed a variety of defects but no specific pattern **(Fig. 4B, Suppl. Fig. S10-11)**.

**Fig. 4.**
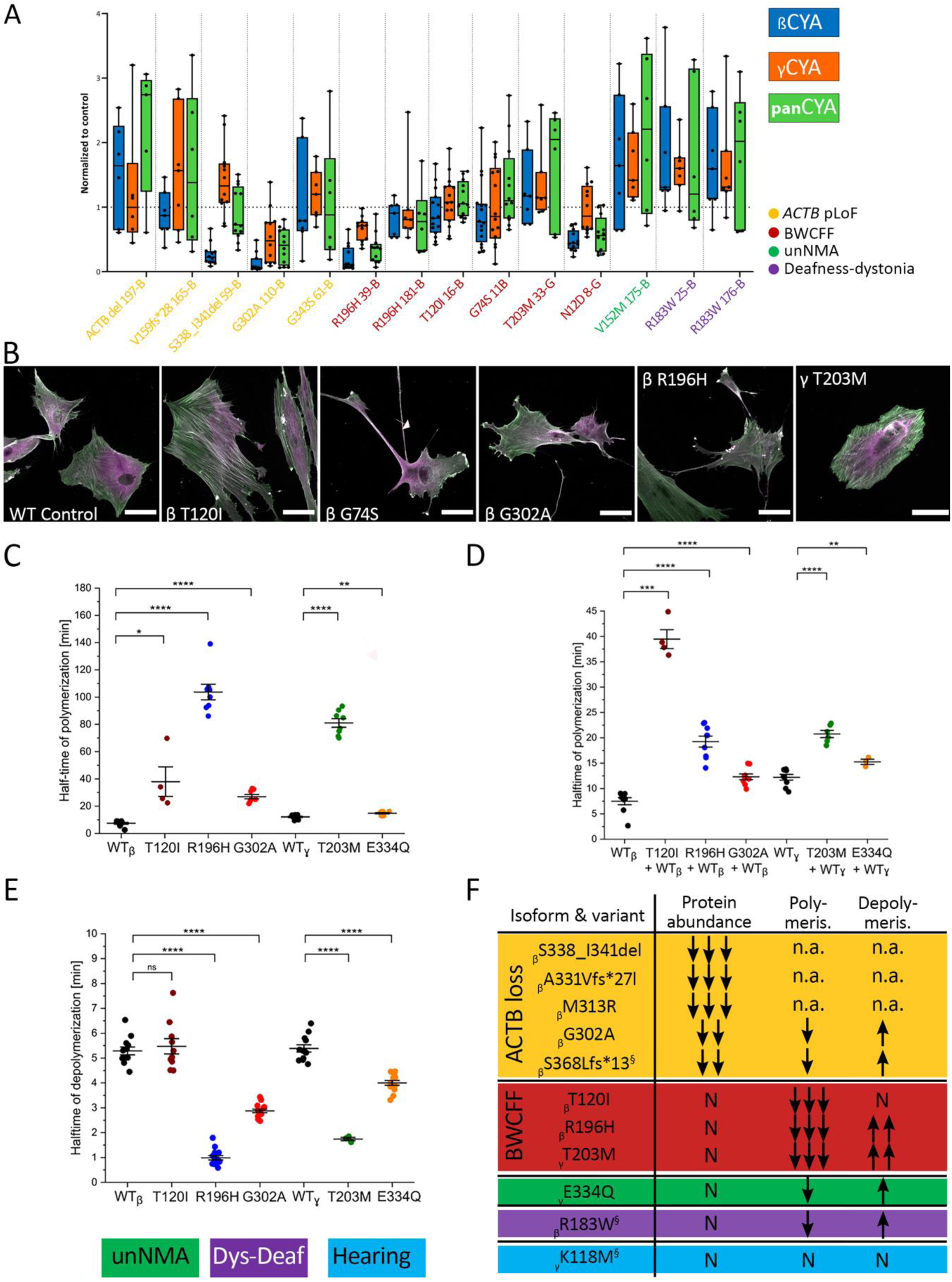
NMAs variants result in diverse abundance, morphology and polymerization anomalies. **(A)** Analysis of bCYA, gCYA, and panactin protein abundance in patient-derived fibroblasts by western blot; data is presented using the box-and-whiskers plot where box contains the 25^th^ to 75^th^ percentiles of the data set and central line indicates the median signal intensity in immunoblots normalized to the total protein amount relative to the control; note that samples were analysed in two different experimental setups using chemiluminescent (samples 59-B, 11B, 110-B, 39-B, 16-B and 8-G)) and fluorescent detections (samples 61-B, 25-B, 176-B, 165-B, 175-B, 181-B and 33-G); representative immunoblots are in shown in the Suppl. Fig. S7 and S9; p-values computed with Kruskal-Wallis test in GraphPad Prism 9 where patients’ derived lines were compared to the actin protein amount normalized to the total protein amount in the individual experiments; **(B)** Immunohistochemical staining with the specific antibodies for bCYA (green) and gCYA (magenta), scale bar 50 µm; note prominent stress fibres co-expressing both actin isoforms in the bT120I fibroblasts, together with the non-filamentous aggregates (arrows) that were also present in the bR196H culture; gT203M culture showed pronounced actin bundles predominantly built by bCYA and underrepresented actin meshwork. **(C)** Salt-induced actin polymerization assessed by pyrene assay: scatter plot demonstrates distribution of the measured half-times from the individual experiments with 2µM bCYA WT, gCYA WT and mutant isoforms, means and SE are indicated; representative traces are shown in the Suppl. Fig. 14 **(D)** same experiment repeated in the 1:1 mixture or the WT and mutants **(E)** Dilution-induced actin filament depolymerization assessed by pyrene assay: scatter plot demonstrates distribution of the measured half-times from the individual experiments with 20 µM F-actin that is rapidly diluted to 0,2 µM with the monitoring of the decrease in fluorescence; representative traces are shown in the Suppl. Fig. 14. **(F)** Summary of the actin mutants characterized in the current and previous studies; previously published mutants are marked with silcrow with the following references βS368Lfs*13(61), βR183W(37) and gK118M(36); n.a. not assessable.

Using selected recombinant proteins, we investigated the potential impacts of CYA variants on actin dynamics. *ACTB*:T120I (BWCFF1), *ACTB*:R196H (BWCFF1) and *ACTG1*:T203M (BWCFF2) showed a striking decrease in filament polymerization rates, and faster depolymerization in the cases of *ACTB*:R196H and *ACTG1*:T203M **(Fig. 4C-E, Suppl. Fig. S12)**, consistent with changes in actin meshwork organization and density **(Fig. 4B)**. All tested mutants formed co-filaments with WT protein. The presence of 50% WT protein in the polymerization experiments attenuated the observed defect in all but one mutant **(Fig. 4D)**. Actin filaments containing both *ACTB*:T120I and WT proteins showed the same polymerisation rate as the pure *ACTB*:T120I mutant. We did not have access to dermal fibroblasts from any patients with unNMA, but we produced and characterized *ACTG1:*E334Q (unNMA)(22). We observed only minor alterations in the polymerization and depolymerization kinetics of this unNMA-associated variant **(Fig. 4C-E)**.

Collectively, these results show that several *ACTB* and *ACTG1* MVs result in highly diverse cellular morphologies and distinct patterns of actin isoform distribution. Moreover, we observed a correlation between the BWCFF variants and the striking changes in polymerization and depolymerization dynamics **(Fig. 4F)**.

### *Actb* knockdown alters morphology and migration in mouse neuronal cells and alters the transcriptional landscape in newborn mouse brains

To investigated the mechanistic basis of the neurological phenotype of *ACTB*-pLOF disorder we performed siRNA-mediated transient knockdown of *Actb* in a mouse neuroblastoma cell line, N2a **(Suppl. Fig. S13-S14)**. This resulted in significant reductions in cell areas and perimeters, as well as significantly increased numbers of neurite-like projections per cell body **(Fig. 5A-C)**. *Actb* siRNA N2a cells showed significant decreased migration after wounding in a scratch assay **(Fig. 5D)** but knock-down did not affect proliferation (assessed by BrdU incorporation), or cell viability (assessed by LDH assay) **(Suppl. Fig. S15)**.

**Fig. 5.**
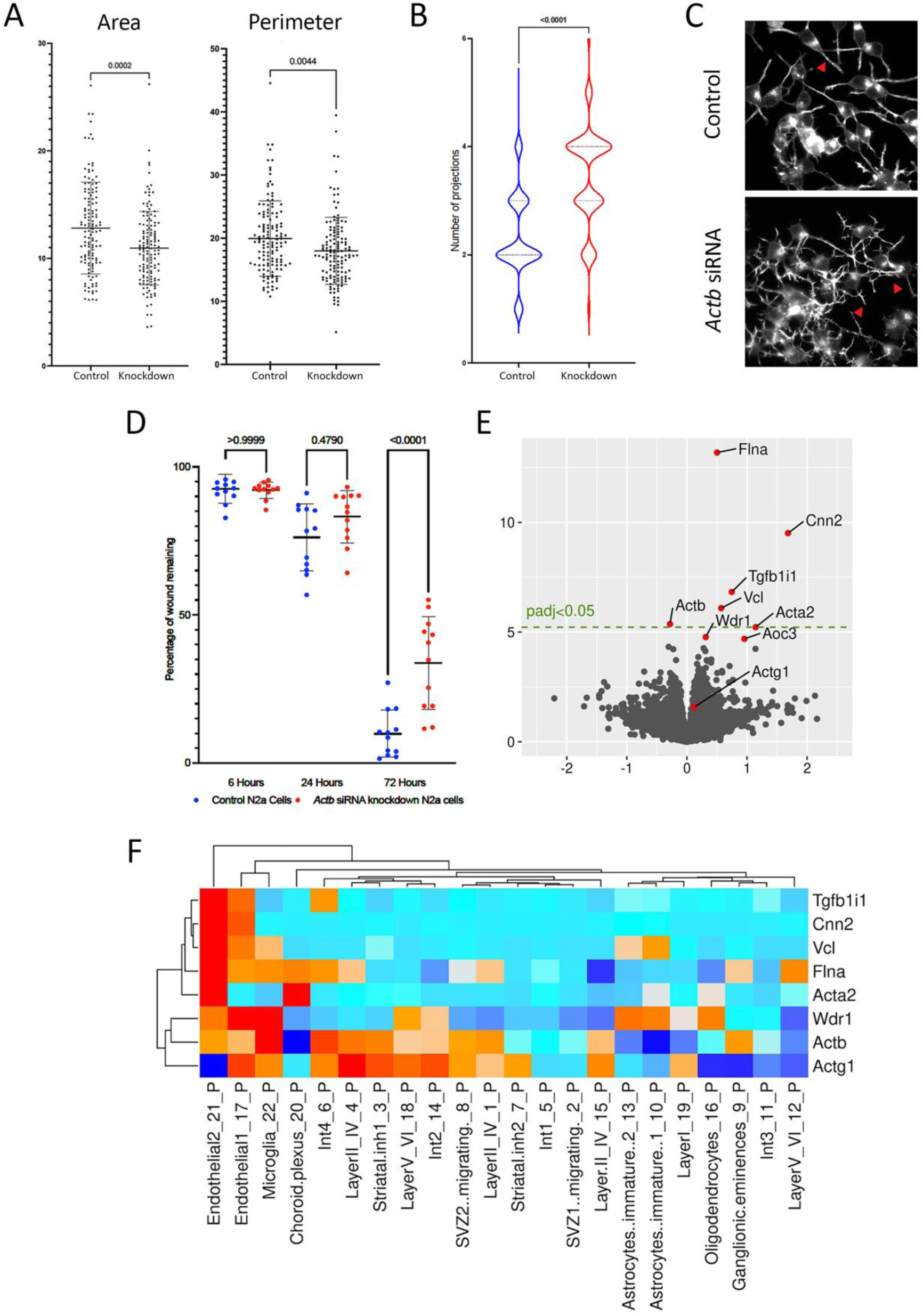
Decreased Actb RNA levels result in abnormalities *in vitro* and *in vivo*. **(A)** N2a morphology analysis showing *Actb* knockdown cells have smaller areas and perimeters. **(B)** The number of neurite-like projections was increased after *Actb* knockdown. **(C)** Note the increased complexity of primary neurite-like extensions, and secondary branches (arrowheads) in N2a *Actb* knockdown cells. **(D)** The motility of N2a cells, assessed with a scratch wound assay, was decreased in N2a cells after *Actb* knockdown. **(E)** RNAseq of male neonatal brains highlighted eight differentially expressed genes. **(F)** The seven transcripts upregulated in *Actb*+/− brains are expressed predominantly in endothelia and microglia.

To explore the *in vivo* consequences of *ACTB*-haploinsufficiency in the maturing brain, we crossed *Actb^Lox^* male mice(23) with female *K14-Cre* mice expressing Cre recombinase in oocytes(24), thus generating offspring with heterozygous intragenic *Actb* deletion (referred to as *Actb*^+/−^) **(Suppl. Fig. S16)**. Compared with non-deleted newborn (D0) male mice, *Actb*^+/−^ males showed no significant differences in body weights and lengths, or brain weights (absolute or normalised by body weight) **(Suppl. Fig. S17)**. Bulk RNAseq **(Suppl. Fig. S18)** on whole neonatal brains showed downregulation of *Actb* in heterozygous mice. The overall reduction was, however, <50%, suggesting upregulation of the intact allele **(Fig. 5E)**. We observed 227 differentially expressed genes (DEGs) at unadjusted P<0.01, but only eight DEGs, including seven upregulated ones, at a P<0.05 adjusted for multiple comparisons. The most marked upregulation was in *Flna* and *Cnn2*, respectively, encoding actin-binding proteins filamin A and calponin 2. While the other CYA gene, *Actg1,* was unchanged in *Actb* knock-down brains, a modest upregulation was noted for the smooth muscle actin *gene Acta2*. qPCR of whole brain RNA from neonates (n=6 from non-deleted mice of each sex, and from deleted mice of each sex) confirmed the trends in altered levels of the transcripts **(Suppl. Fig. S19)**. Next, we utilised pseudo-bulk gene expression profiles from neonatal non-mutant mouse cerebral cortex single-cell RNA-seq(25), noting that all seven transcripts upregulated in *Actb*^+/−^ brains are typically expressed predominantly in endothelia cells and microglia **(Fig. 5F)**. We found that the ‘marker genes’ defining endothelial and microglial cell-clusters were not significantly overexpressed in our bulk RNASeq data from the *Actb*^+/−^ brains **(Fig. 5F)**.

Overall, these results show that *Actb* haploinsufficiency manifests *in vitro* as altered morphology and migration in mouse neuronal cells and likely results in aberrant or ectopic expression of a set of genes in non-endothelial and non-microglial brain cells. These observations provide an insight into possible mechanisms underlying the neurological features of ‘*ACTB*-pLOF disorder’.

### BWCFFS causing variants show abnormal neuronal migration in neurospheres

To understand the impact of CYA MVs at the organism level, we generated two heterozygous knock-in mouse models carrying the variant T120I in *Actb* or *Actg1*. The efforts did not result in a F0 mouse suitable for further breeding **(Suppl. note)**. Therefore, we used patient-derived iPSCs to study neuronal proliferation and migration. In this human BWCFF model, we performed neuronal differentiation with the variants *ACTB* T120I (BWCFF1) and *ACTG1* T203M (BWCFF2) and a control iPSC line. Patients with these variants showed prominent pachygyria, suggesting a neuronal migration defect (patients 16-B and 33-G in Supp Table S2). The migratory and proliferative capacity of neuronal cells was assessed in a neurosphere migration assay representing a heterogeneous population of neuronal progenitor cells and immature Tuji-positive neurons replicated in multiple batches **(Suppl. Fig. S20)**. We detected significant differences in neutrosphere expansion in both mutants. These defects were particularly pronounced with *ACTB* T120I **(Fig. 6, Suppl. Movies 1-3)**. Interestingly, both mutant cell lines showed nearly normal neurite expansion capacity **(Fig. 6E)**. However, we observed only minimal cell migration along the neurites in *ACTB* T120I. Moreover, the relation between the external cell expansion area and proliferating neurosphere core mass suggests that cellular expansion in *ACTB* T120I reflects neuronal proliferation rather than actual migration **(Fig. 6F)**.

**Fig. 6.**
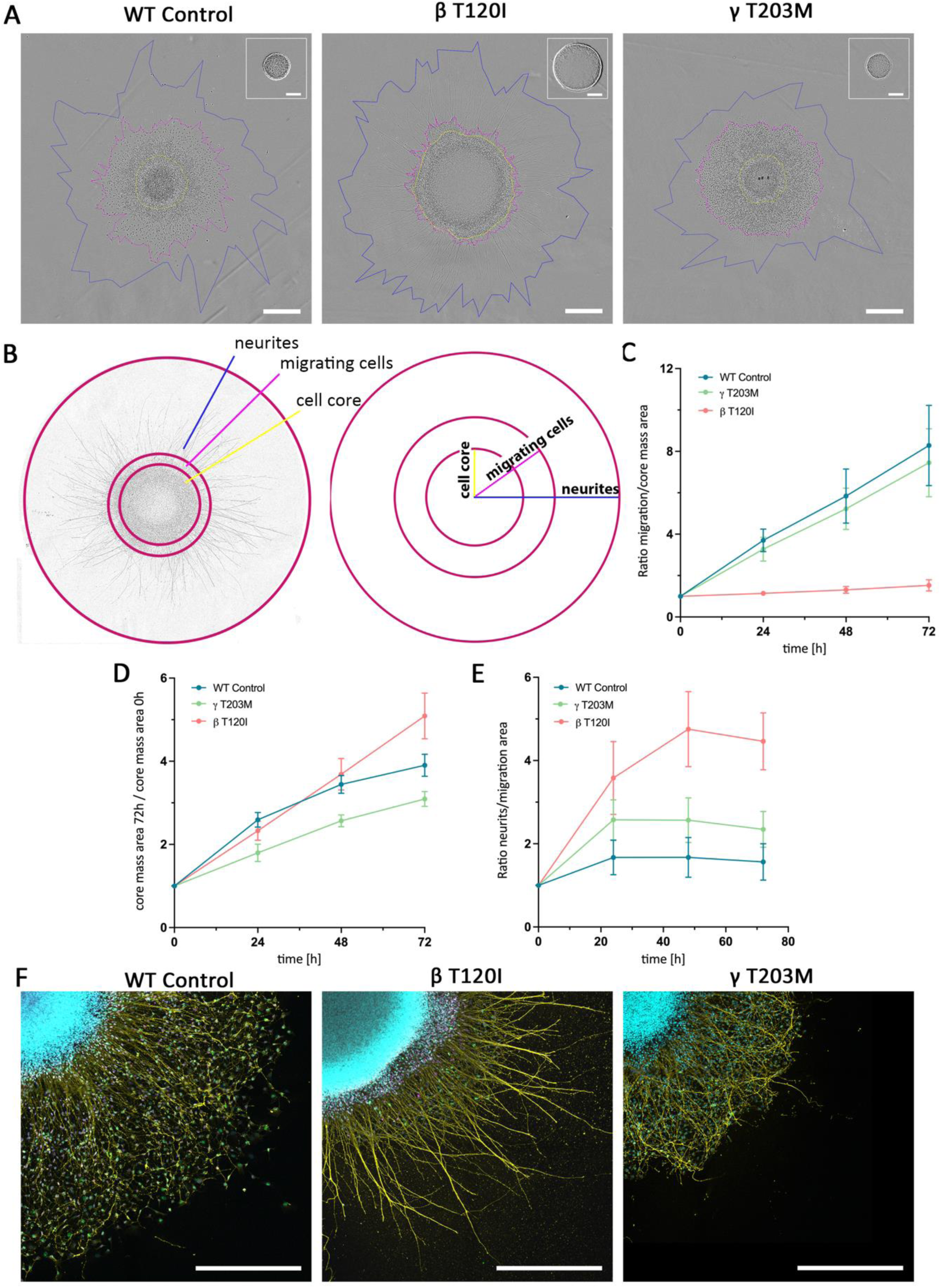
BWCFF variants in βCYA result in abnormal neuronal migration. **(A)** Phase contrast images (IncuCyte^TM^) of the neuronal spheroid cultures at the end point of the migration assay from the control iPSCs and two BWCFF iPSC lines, scale bar 500 µm; an additional image in the right upper corner shows the spheroid at the start point of the experiment, scale bar 200 µm; yellow mark delineates cellular core of the spheroid defined as an area of the tightly packed nuclei, magenta indicates the area of the migrating cells, and blue line marks the area occupied by the neurites extending from the spheroid. **(B)** Schematic overview of the measured distances used in the analysis as well as the evaluation strategy applied to analyse neuronal migration in the spheroid assay. **(C)** Quantification of the cell displacement from the spheroid along the processes after 72 hours as a ratio of migration area to the core area **(D)** Quantification of the enlargement of the cell core area proportional to the spheroid area at the start point. **(E)** Quantification of the length of the neurites in relation to the area occupied by the migrating cells after 72h; the graphs show mean values ± SD **(F)** Immunohistochemical staining for SOX2 (magenta), TUJ1 (yellow) und Ki-67 (green) combined with DAPI (blue); scale bar 500 µm.

## Discussion

The combination of large datasets, clinical studies and complimentary disease-relevant laboratory models led to a coherent functional classification of the NMA spectrum. It finally resulted in the delineation of eight distinct NMAs, demonstrating extraordinary pleiotropy and a spectrum of disease severity associated with *ACTB* and *ACTG1* variants **(Fig. 7)**. Based on our data, NMAs can be categorised into three major groups. First, genomic variants resulting in decreased or absent expression or production of unstable b-actin are associated with relatively mildly affected postnatal function. Second, MVs resulting in stable expression of actin with severely impaired poly-/depolymerization dynamics are associated with significantly abnormal embryonic development (abnormal migrations of the neurons and most probably also neural crest cells) and progressively abnormal postnatal function in multiple tissues. Third, MVs resulting in stable expression of actin with normal or slightly abnormal polymerization dynamics are likely associated with altered function and survival of one or a few specific cell types rather than embryonic migration. The divergence in the spectrum of *ACTB* and *ACTG1* variants and the associated clinical features clearly demonstrate the functional differences of the two human CYAs. This delineation of disorders has direct implications for the diagnosis and treatment of patients, showing the power of large systematically collected cohorts in rare diseases. We hypothesise that this remarkable number of conditions associated with the two genes reflects the large number of cellular interactions of CYAs, with downstream effects being context-dependent and clinical consequences are therefore highly variant-specific.

**Fig. 7.**
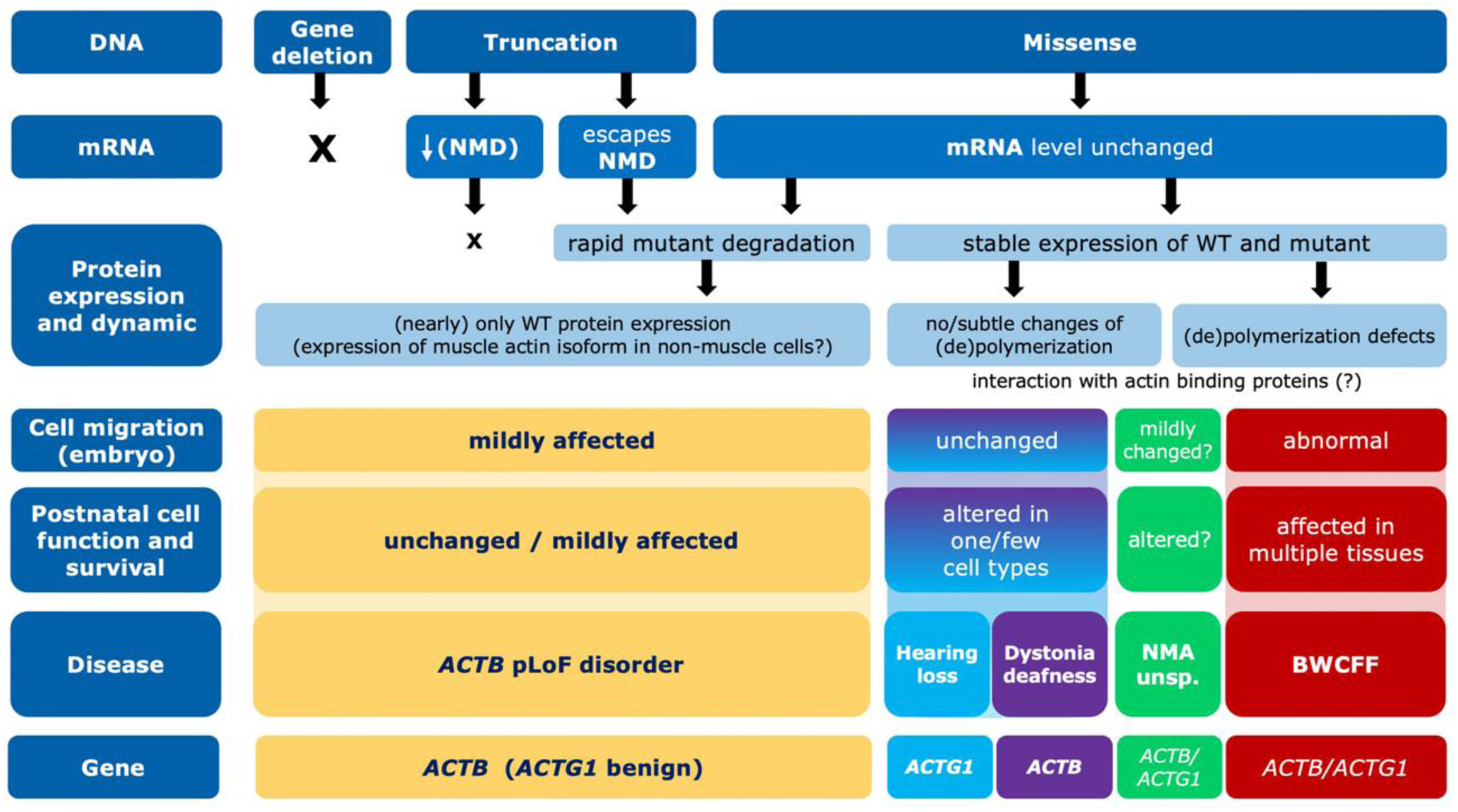
Functional classification of non-muscle actinopathies.

Neurodevelopmental and neurological features are the predominant characteristics of many NMAs. This is in line with the known importance of CYAs for neurodevelopment, neural function, and memory(26, 27). Variants in several other genes that control cytoskeletal dynamics, such as *DIAPH1*, *RAC1*, *CYFIP*, have been shown to result in neurodevelopmental phenotypes similar to those of NMAs. Our findings highlight the increasing recognition of the importance of fine-tuned cytoskeletal control and dynamics for normal development. Furthermore, understanding of actin dynamics in the neurodevelopmental context is of greater importance as several other monogenic neurodevelopmental disorders are caused by variants in genes encoding actin regulators and interactors (28–33). This pathway appears to be particularly susceptible to variant-specific effects as several of these genes result in allelic disorders(34, 35).

Deafness is another common feature of several NMAs, reflecting the central role of actin filaments in maintaining the functional and structural integrity of inner ear hair cells. Actin filaments in inner ear hair cells contribute critically to the structural integrity of stereocilia, mechanotransduction, regulation of stereocilia length and maintenance and repair of these sensory structures. Polymerization studies revealed a number of allosteric effects, typically associated with moderate changes in actin dynamics but more pronounced in K118N, the most common *ACTG1* variant leading to hearing loss(36). We have shown that *ACTB* R183W results in a stable protein with approximately 2-fold increase in polymerization half-time and ATP-turnover as well as impaired interaction with NM-2A(37). As multiple myosin isoforms are produced in cochlear hair cells(38), impaired interaction with these myosins and changes in filament dynamics with consequences for stereocilia length and maintenance are likely to contribute to the specific vulnerability of cochlear hair cells. Craniofacial dysmorphism and organ malformations are also common and are explained by the importance of CYAs for the development and migration of cells, such as the cells of the neural crest(39).

In conclusion, the present work will improve the diagnosis and management of NMA patients. The functional insights will pave the way for new and efficient treatment strategies. These results also reflect the considerable amount of variant-level collaborative clinical trials and multi-modal mechanistic studies that may be required to realise the promise of precision medicine for rare allelic disorders.

## Materials and Methods

### Study approval

The study was approved by the Institutional Review Board of the TU Dresden (EK-127032017 and BO-EK-341062021) and the Central Manchester (02/CM/238) and local IRBs from refereeing physicians. All patients were informed of their participation prior to participating. Written informed consent was also received for all photographs of patients included. The record of informed consent has been retained.

### Data availability

Data sets supporting the results of this article are available in the supplementary material or could be requested from the corresponding authors.

### Simulation of all possible nucleotide substitutions within genomic regions of ACTB and ACTG1

For each of the 3454 reference alleles (REF) in *ACTB* transcript NM_001101.5 (hg38 7:5527148-5530601) and 2829 REFs in *ACTG1* transcript NM_001614.5 (hg38 17:81509971-81512799), all three possible single-nucleotide alterations were simulated *in silico*, resulting in a list of 6283*3 = 18849 alternate alleles (ALT) for both transcripts. The functional consequences of all possible single nucleotide alterations were annotated according to Ensembl Variant Effect predictor(40).

### Compilation of *ACTB* and *ACTG1* population variants and public databases

We retrieved **gnomAD v3.1.1** [hg38] genomic variants from genomic regions 7:5527148-5530601 [ACTB] and 17:81509971-81512799 [*ACTG1*].(14) To prevent redundancy with v3.1.1., the **gnomAD v2.1.1** [hg19] variants were downloaded for exomes only; exons were defined by gnomAD standard transcripts ENSG00000075624.9 [*ACTB*] and ENSG00000184009 [*ACTG1*], including gnomAD’s default padding of 75 nt. The Ensembl tool “Assembly Converter” converted the genomic positions of gnomAD v2 hg19 variants to hg38. We exploited each of the different gnomAD subsets separately: controls (2148 variants), non-topMed (3107 variants), non-neuro (3476 variants), non-cancer (3605 variants). We retrieved 2184 genomic variants passing **TOPMed** Filter (i.e. flagged as PASS in VCF “FILTER” column) (Freeze8 on GRCh38, access on 01.11.2021) within the genomic regions 7:5527148-5530601 [ACTB] and 17:81509971-81512799 [ACTG1].(16) We retrieved 2280 variants from https://grch38.pggsnv.org/index.html (access on 31.07.2021) within the genomic regions 7:5527148-5530601 [*ACTB*] and 17:81509971-81512799 [*ACTG1*].(**15**) 754 variants were retrieved from **COSMIC** catalogue using gene names *ACTB* and *ACTG1*, 440 and 309 variants respectively. The query for **cBioPortal** mutations (not including structural variants and copy number alterations) was performed using an user-defined list containing HUGO gene symbols *ACTB* and *ACTG1* (access on 31.07.2021 with the query of 46,305 patients, representing 48,834 samples in 188 studies).(**41, 42**) This query identified 389 unique and a total number of 508 variants. **ClinVar** variants were retrieved with the NCBI search term using hg38 coordinates of both genes (access on 03.11.2021)(**43**): (7[CHR] AND 5527148[CHRPOS]:5530601[CHRPOS]) and (17[CHR] AND 81509971[CHRPOS]:81512799[CHRPOS]). From **LOVD** the search for all variants in *ACTB* (NM_001101.3 transcript reference sequence) and *ACTG1* (NM_001614.3 transcript reference sequence) resulted in 110 and 195 variants respectively (access on 03.11.2021).(**44**)

### Clinical and genomic analysis of patient data

#### Compilation of the clinical cohort

Patients were recruited primarily at the Institute for Clinical Genetics, University Hospital Dresden (N.DD) as part of the active patient registry within EJPRD funded PredACTINg project as well as at the Manchester Centre for Genomic Medicine within the NHS sequencing projects (S.B) after the rare variant in ACTB or ACTG1 was discovered in the clinical genetic testing. Additionally, patients were referred to N.DD. and S.B. by multiple clinicians for the second opinion regarding variant interpretation, management advice or specifically for this study. Clinical and molecular data was collected with the standardized proforma (Supplementary data). Clinical photographs were available for 160 patients and were independently assessed by four authors experienced in dysmorphology and specifically familiar with the BWCFF (N.DD, S.B., G.V.M., D.P.). Additionally, PubMed was searched for publications using terms “ACTB” OR “ACTG1” OR “Baraitser-Winter syndrome” OR “Baraitser-Winter-Cerebrofrontofacial syndrome” OR “Dystonia-deafness syndrome” OR “ACTG1 hearing loss” OR “ACTB neurodevelopmental disorder” OR “ACTG1 neurodevelopmental disorder”. We extracted reported data from identified publications and added follow up information where available. In addition, we screened ClinVar database for (likely) pathogenic variants and variants of uncertain significance and contacted responsible laboratories requesting clinical information.

#### Genomic and phenotypic-led approach for classification of the clinical cohort

For classification of the cohort into relevant NMA groups, first all individuals were separated according to the gene in which their variant was located. Next, pLOF and PAV (missense or in-frame) variants were separated. Clinical features of patients with pLOF variants were manually analysed. The identified clinical features identified from these patients were used to identify patients with PAVs whose phenotype overlapped with those with pLOF variants. Next, individuals with recurrent PAVs were identified, their clinical features were manually analysed and genotype-phenotype relationships were extracted. Clinical data from these individuals were used to generate diagnostic criteria for each NMA. These diagnostic criteria were applied to individuals with non-recurrent variants and additional patients were identified for each sub-group. The clinical and mutation spectrum of each NMA was compiled using the combined data. The approach is summarised in Supplementary Figure S3.

### GestaltMatcher facial analysis

To investigate the similarities between the three groups, BWCFF with mutations in *ACTB* (BWCFF_ACTB), with mutations in *ACTG1* (BWCFF_ACTG1), and LoF we analysed 57 images (BWCFF_ACTB: 23; BWCFF_ACTG1: 15; LoF: 19), including the patients reported in this work and the previously published patients in the GestaltMatcher Database(45) (GMDB; https://db.gestaltmatcher.org). We first encoded each image to a 512-dimensional feature vector by GestaltMatcher, and further utilized tSNE(46) to visualize the distribution of the four groups of patients in the two-dimensional space.

Moreover, we wanted to compare the similarities among the three groups with the control distribution sampled from GMDB. Because the facial phenotypic similarity between two patients was quantified by the cosine distance, when the distance was smaller, the two patients were more similar as they are closer in the space. We then examined the mean pairwise cosine distance between patients among the three groups. We sampled control distributions from 1,499 images with 321 different disorders in the GMDB that were not included in the training of GestaltMatcher and calculated the mean pairwise distances between two cohorts stemming a) from the same syndrome and b) from two different syndromes (shown in blue (a) and red (b) in Supplementary Figure 4 (b)-(d). We further derived a threshold to decide whether two cohorts stem from the same or different syndromes by ROC analysis, resulting in a final threshold of *c* = 0.896. To assess the similarity between the two cohorts *C*_1_ and *C*_2_, we computed their mean pairwise cosine distance *d*(*C*_1_, *C*_2_), and compared it to the threshold c. Additionally, we conducted 100 subsampling iterations from each cohort to generate subcohorts, calculating the mean pairwise cosine distance for each iteration. If at least 50% of these 100 subsampled comparisons yielded values above the threshold c, it would provide evidence suggesting that the two cohorts stem from different syndromes.

### Fibroblasts experiments

#### Establishment of the patient-derived and control fibroblast cultures

Primary dermal fibroblasts were obtained following 3 mm cutaneous punch biopsies and cultured in BIO-AMF™-2 Medium (Biological Industries USA, Cromwell, CT, USA). For sub-culturing, primary fibroblasts were washed twice with 1× dPBS and detached at 37 °C for at least 3 minutes with 0.05% Trypsin/EDTA (Gibco®; Thermo Scientific, Waltham, MA, US). Cells were resuspended in BIO-AMF-2 medium, seeded onto Corning plasticware (Corning, NY, US) and maintained in BIO-AMF-2 medium at 37 °C in the presence of 5% CO2. Cultures were continued for a maximum of 3 passages afterwards the cultures were cryopreserved in multiple cryovials for long term storage at −150°C. 90% FBS + 10% DMSO was used as a freezing medium. Thawing of the frozen cells was performed rapidly at a 37°C water bath. Thawed cells were centrifuged at 240x*g*/5min and resuspended in fresh BIO-AMF™−2 medium into a new flask. Only cultures in early passages (maximum 7) have been used in the experiments. Cultures were labelled with the actin amino acid change and a patient ID corresponding to the ID in the Supplementary Table 2 (request from corresponding author, e.g. G343S 61-B).

Primary dermal fibroblasts from nine healthy adult individuals were obtained and stored following the same procedure described above. These cell lines are labelled as Control_1, Control_2, as well as Control_6 through Control_12. Only cultures in early passages (maximum 7) were used in the experiments. Additionally, three control cell lines were acquired from the Coriell biobank: GM00013 (passage 13), GM04390 (passage 11), and GM05294 (passage 8) and labelled Control_3, Control_4 and Control_5 respectively. Coriell cultures were only used for the transcriptome analysis.

#### Phase contrast microscopy of the fibroblasts

For the cell cultures G74S 11-B, G302A 110-B, R196H 39-B, T120I 16-B, T203M 33-G, and control cell line Control_10 phase contrast images were obtained with a Nikon Eclipse TS100 microscope using 10×/0.25 NA objective and a Nikon DS-Fi2 camera (Nikon, Minato, Tokyo, Japan). Fibroblast cultures R196H 181-B, R183W 25-B, V159fs*28 165-B, S338_I341del 59-B, G302A 110-B, G343S 61-B, V152M 175-B, R183W 176-B, R183W 178-B, T203M 33-G, as well as Control_11 and Control_12 were imaged with a Keyence BZ-X800 microscope using a 10X phase contrast objective.

#### Cell lysis and western blot analysis

##### Cell lines N12D 8-G, G74S 11-B, T120I 16-B, R196H 39-B, G302A 110-B, S338_I341del 59-B Control_8 Control_10

Sample preparation and western blot analysis were done as described previously(4). Briefly, total cell lysate of final concentration of 1×10^4^ cells per µl of lysis buffer were loaded per lane on Tris Tricine SDS PAGE. Total protein concentration was normalized based on total intensity of Coomassie staining per lane. Gels used for immunoblotting had their protein load per lane adjusted to ±15% of the control cells. Based on the Ponceau S staining and pre-stained marker, nitrocellulose membranes were cut into 2 segments and probed with antibodies for target proteins. Primary antibodies were incubated overnight in blocking solution at 4 °C on a rocking table (Suppl. Table 4). Following three TBS-T washes, membranes were incubated in secondary antibody in blocking solution for 1 h and washed thrice in TBS-T prior to developing. Signals were developed with the SuperSignalTM West Dura Extended Duration Substrate (Thermo Scientific) and images were obtained with the ChemiDoc MP Imaging system using ImageLab software (Bio-Rad). At least three biological replicas of different passages of every culture have been analysed. Each replica was blotted at least 4 times. Beta-, gamma-, pan actin signals in patients immunoblots were normalized to the signal in the immunoblots from the control cultures. Statistical analysis was performed with OriginPro 2021 software (OriginLab Corporation, Northampton, Massachusetts, USA) where the normal distribution of each data set was checked and, depending on the result, either ANOVA and Tukey test or Kruskal-Wallis ANOVA and Dunn’s test were performed. Outlier analysis performed using Grubb’s Test (p=0.05) with a maximum of one outlier per dataset.

##### Cell lines G343 61-B, R183W 25-B, R183W 176-B, V159fs*28 165-B, V152M 175-B, T203M 33-G, R196H 181-B, ACTB del, Control_11

Cells grown to approximately 70% confluency were washed with room temperature dPBS twice. 0.6 ml of RIPA buffer (Santa Cruz, sc-24948) was added to the monolayer cells in a T25 flask and gently rocked for 15 minutes at 4° C. Adherent cells were removed with a cell scraper and the lysate was transferred into a microcentrifuge tube. Lysate was incubated for 5 minutes on ice before centrifugation at 14,000 x g for 10 minutes at 4° C and supernatant was collected. Total protein concentration was measured with the Qubit fluorometer and the Qubit Protein Assay Kit (Thermo Fischer Scientific). Approximately 7 µg protein was separated on 4-12% NuPAGE Bis Tris gels (Thermo Fischer Scientific) by electrophoresis (90 V, approx. 2.5 hours) under reducing conditions with 6 µl of ProSieve QuadColor prestained protein marker loaded. Proteins were blotted to a 0.2 µm pore nitrocellulose membrane with the iBlot2 system (Thermo Fischer Scientific). Revert™ 700 Total Protein Stain (LI-COR, 926-11011) was performed for normalization of target protein signal intensities and images of wet membranes were acquired with the Odyssey SA Li-Cor Infrared Imaging system in the 700 nm channel (LI-COR). Subsequently, membranes were blocked overnight at 4° C in 5% (w/v) skim milk in TBS-Tween20 (TBS-T). The next day membranes were incubated with primary antibodies diluted in 0.5% (w/v) skim milk in TBS-T for 2 hours at room temperature (see Suppl. Table S4 for antibodies and dilutions used). After 3 washing steps with 0.5% (w/v) skim milk in TBS-T membranes were incubated with secondary antibodies for 1 hours at room temperature with 3 subsequent washes with TBS-T followed by 3 washes with TBS. Next, images of dried membranes were acquired again with the Odyssey Imaging system in the 800 nm channel. Analysis of western blot signals was performed with the CLIQS Gel Image Analysis Software (TotalLab). Target protein signals were normalized to corresponding total lane signals of total protein stain and finally patient samples were normalized to the respective control cell line. Results were obtained from three different lysates of independent cell passages with up to three replicates for each sample.

### Immunofluorescence microscopy of primary fibroblasts

Immunofluorescence experiments were performed as described previously(4) with slight modification. Specifically, cells were seeded at a density of 2 × 10^4^ on 10 mm glass coverslips in 24-well plates and cultured for 48 hours prior to collection. Cells were washed twice with pre-warmed DMEM/F-12 medium and fixed for 30 min with 1% PFA diluted in DMEM/F-12. Following two washes with 1X dPBS, cells were permeabilized with ice cold methanol for 5 min. Samples were washed twice with 1X dPBS (Gibco®; Thermo Scientific) and blocked for 1 hour at 23 °C in sterile-filtered 2% BSA/dPBS blocking solution prior to antibody labelling. Primary and secondary antibodies were diluted in blocking buffer, as shown in Supplementary Table 2 (request from corresponding author), and were applied for 1 hour and 30 min, respectively, with three intermittent blocking buffer washes. Samples were washed thrice with 1× dPBS, rinsed with ddH_2_O and mounted in ProLongTM Gold Antifade Mountant (Thermo Scientific). Z-stack images (0.3 µm slices) were obtained with the Leica TCS SP8 Confocal Microscope (Leica Microsystems, Solms, Germany) using a 63x Oil/1.4NA objective or Zeiss LSM980 Confocal Microscope (Carl Zeiss Microscopy GmbH, Jena, Germany) using a 63x Oil/1.46NA objective. Image analysis was performed using Image J software (NIH, Bethesda, ML, US). For displaying purposes Z-stack slices were displayed as Sum Slice image projection with each channel adjusted to have 0.3% saturated pixels in the final image. Cell line Control_10 was used as a healthy control.

### Transcriptome analysis of primary fibroblast cultures

#### Library preparation and sequencing (biological and technical replicates)

Transcriptome analysis was done using primary fibroblast cultures from 15 patients with missense variants in *ACTB*, 3 patients with variants in *ACTG1* and 7 control cultures from healthy adult individuals (Control_1, Control_2, Control_6 through 10) as well as three fibroblast cultures from Coriell biobank (Control_3 trough 5). All cultures were harvested at early passages of maximum 7, except cultures from Coriell available in the later passages (up to passage 15).

Fibroblasts were seeded in T75 flasks and harvested at ~70% confluence. The culture was performed twice to produce biological replicates. RNA was extracted using the miRNeasy Mini Kit (Qiagen) according to the manufacturer’s instructions. On column DNA digestion was included to remove residual contaminating genomic DNA. All experiments were performed in triplicate, meaning that RNA was independently extracted three times from each culture. In total we performed six library preparation per individuum (patients and controls). For library preparations, the TruSeq Stranded mRNA Library Prep Kit (Illumina) was used according to the manufacturer’s protocol, starting with 1 μg total RNA. All barcoded libraries were pooled and sequenced 2 × 75 bp paired-end on an Illumina NextSeq500 platform to obtain a minimum of 10 million reads per sample. Raw reads from Illumina sequencers were converted from bcl to fastq format using bcl2fastq (version v2.20) allowing for 1 barcode mismatch.

### Bioinformatic pipeline

The quality of the obtained fastq files was initially checked by FastQC v0.11.4 (https://www.bioinformatics.babraham.ac.uk/projects/fastqc/) followed by adapter removal and quality trimming using Trim Galore v0.4.2 (http://www.bioinformatics.babraham.ac.uk/projects/trim_galore/). Mapping of reads to the human reference genome (GRCh38 Ensembl release 95) was done using STAR v2.5.3a with standard settings(47) and duplicates were marked and removed using Picard tools v1.141 (http://broadinstitute.github.io/picard/). Quality analysis of mapped reads was done using RSeQC v3.0.0(48) to analyze read distributions across gene bodies. Raw read counts per gene were determined by counting gene-specific reads in exons of protein-coding genes using FeatureCounts v1.5.3(49). Finally, a gene expression data matrix was created by removing genes without any reads and lowly expressed genes (less than 1 read per million in more than 50% of samples) followed by cyclic loess normalization(50) resulting in normalized log2-counts per million for 12,772 protein-coding genes that were measured in each sample. The average gene expression levels per patient or control are provided in Supplementary Table 3.

### Similarity of gene expression profiles and differential gene expression analysis

Principal component analysis of average genome-wide gene expression profiles of patient and control samples was done to analyze if the different disease and control groups form separate clusters (R function prcomp). Further, similarity of genome-wide gene expression profiles of patient and control samples was determined by computing the Pearson correlation coefficient for each pair of samples utilizing the average expression profile of each patient and control sample (R function cor). The corresponding correlation matrix was visualized as heatmap (Suppl. Figure 12).

### Protein production and characterization

#### Generation of plasmid and baculovirus

The pFastBac vectors carrying the sequence of interest were constructed and generated as described previously(51). Mutations in the sequence were introduced via site-directed mutagenesis with oligonucleotides encoding the desired mutation. Baculovirus was generated as described in the Bac-to-Bac™ Baculovirus Expression System manual (Thermo, Waltham, USA). In short, pFastBac vectors carrying the sequence of interest were transformed into DH10EMBacY™ Escherichia coli to generate the recombinant bacmid. Sf-9 insect cells were then transfected with the recombinant bacmid to generate the recombinant baculovirus. Production of the recombinant actin WT or mutant was started by infecting 1.8 · 10^6^ cells/mL with 1:50 of the corresponding virus stock. Cells containing the protein of interest were harvested 3 days post infection and stored at −80 °C until used for purification.

#### Purification of recombinant cytoskeletal actin WT and mutants

Recombinant human cytoskeletal actin WT and mutants were purified from Sf-9 insect cells using an actin-thymosin β4 His8 fusion construct as described by Noguchi et al.(52). In short, Sf-9 cells were resuspended in lysis buffer (10 mM TRIS pH 7.8, 5 mM CaCl_2_, 1.25% TRITON X-100, 1 mM ATP, 100 mM KCl, 7 mM β-mercaptoethanol, 1 mM PMSF, 100 µg/mL TAME, 80 µg/mL TPCK, 2 µg/mL Pepstatin, 5 µg/mL Leupeptin) and sonicated to disrupt the cells. The lysate was cleared by centrifugation and the supernatant incubated with 2 mL of lysis buffer-equilibrated Pure Cube^TM^ NiNTA column material (Cube Biotech, Monheim am Rhein, Germany) per litre of expression culture for 2 hours rotating at 4 °C. The material was washed with 25 column volumes of wash buffer 1 (10 mM TRIS pH 7.8, 5 mM CaCl_2_, 10 mM imidazole, 200 mM KCl, 1 mM ATP) followed by 25 column volumes of wash buffer 2 (10 mM TRIS pH 7.8, 5 mM CaCl_2_, 10 mM imidazole, 50 mM KCl, 1 mM ATP). The protein was eluted with 250 mM imidazole and the purity of the eluate was verified via SDS-PAGE. Fractions that contained the majority of the protein were pooled and dialysed against G-buffer (10 mM TRIS pH 7.8, 0.2 mM CaCl_2_, 0.1 mM DTT, 0.1 mM ATP) over night to remove imidazole. The dialysed sample was then digested with 1:300 weigth/weigth of ɑ-chymotrypsin from bovine pancreas (Merck KGaA, Darmstadt, Germany) to remove the thymosin β4 His8-moiety including the linker to yield the pure actin with native N- and C-termini. The reaction was quenched after a minimum of 45 minutes by the addition of 1 mM PMSF. The exact time of digest strongly depends on the age of the used batch of ɑ-chymotrypsin. The sample was concentrated to 10 – 15 mg/mL and polymerization of actin was induced by the addition of 100 mM KCl and 5 mM MgCl^2^. The polymerization reaction was incubated for at least 3 hours at room temperature and then moved to 4 °C overnight. On the following day, the F-actin was sedimented by centrifugation at 130,000 × g for 1 hour at 4 °C. The pellet was washed with G-buffer and finally resuspended in 0.5 – 1 mL G-buffer using a Dounce homogenizer. The sample was dialysed against a total of 5 litres of G-buffer supplemented with 0.1 mM PMSF over 4 days. The buffer was changed at least 3 times over that period. After dialysis the protein sample was centrifuged at 15,000 × g for 15 minutes to remove precipitate. The pure protein was flash-frozen in liquid nitrogen and stored at −80 °C.

#### Assays probing polymerization and depolymerization of actin

Pyrene-actin based assays to monitor polymerization and depolymerization of actin filaments were performed as previously described(53) with some slight modifications. To determine the rate of actin polymerization Mg^2+^-ATP-G-actin was supplemented with 5% pyrene-labelled Mg^2+^-ATP-G-actin (WT) as a tracer to a final concentration of 10 µM. 20 µL of this solution were placed in a black flat bottom 96-well plate (BrandTech Scientific, USA). The polymerization reaction was monitored as a function of increasing pyrenyl-fluorescence in a Synergy 4TM microplate reader (BioTek Instruments, Winooski, USA) using the built-in filter set (Excitation: 340/30 nm, Emission: 400/30 nm). Polymerization was induced by applying 80 µL of 1.25x polymerization buffer to a final concentration of 10 mM TRIS, pH 7.8, 100 mM KCl, 5 mM MgCl_2_, 0.5 mM EGTA, 0.1 mM DTT and 0.1 mM ATP using the built-in pipetting function. To determine the rate of depolymerization Mg^2+^-ATP-G-actin was polymerized at 20 µM in the presence of 5% pyrene-labelled Mg^2+^-ATP-G-actin (WT) over night at 4°C. 3 µL of the F-Actin solution were placed in a black flat bottom 96-well plate and rapidly diluted by applying 297 µL of G-Buffer (10 mM TRIS, pH 7.8, 0.2 mM CaCl_2_, 0.1 mM DTT and 0.1 mM ATP). The dilution-induced depolymerization of the actin filaments was monitored using the settings mentioned above. The apparent half-time of the polymerization and depolymerization reaction was calculated by applying a single-exponential fit to the kinetic traces.

#### Thermofluor assay

Thermal stability of WT and mutant proteins was assessed using the Thermofluor-assay. The assay utilizes the fluorescent dye Sypro Orange, which shows an increase in fluorescent intensity upon binding to hydrophobic core regions of proteins that become exposed during thermal denaturation. Ca^2+^-ATP-G-actin was converted to Mg^2+^-ATP-G-actin by incubation with magnesium-exchange-buffer (10x, 10 mM EGTA, 1 mM MgCl_2_) for 2 minutes on ice prior of the experiment. 0.2 mg/mL of Mg^2+^-ATP-G-actin and 5x Sypro Orange (Life Technologies, Carlsbad, USA, stock: 5000x) were mixed in assay buffer (10 mM TRIS, pH 7.8, 0.1 mM MgCl^2^, 1 mM EGTA, 0.1 mM DTT, 0.1 mM ATP) to a final volume of 25 µL. The samples were placed in a MicroAmpTM 48-well plate (Applied Biosystems, Waltham, USA). The change in fluorescence intensity over a linear temperature gradient (1 °C/min) was measured in a StepOne Real-Time PCR-System (Applied Biosystems, Waltham, USA). The melting temperature was derived from the peak value of the first derivative of the melting function.

#### Nucleotide exchange assay

The rate of nucleotide exchange of Mg^2+^-ATP-G-actin was determined as previously described(37) using the intrinsically fluorescent ATP analogue ε-ATP (Jena Bioscience, Jena, Germany).

### Mouse neuroblastoma N2a line experiments

#### *Actb* silencing

Mouse neuroblastoma N2a cells were passaged and cultured using standard techniques and transfected with *Actb* siRNA in 25 nM FlexiTube siRNA Premix (Qiagen) at desired concentrations. A one way ANOVA with multiple comparisons was used to define the significance of the knockdown of *Actb* expression in relation to *Gapdh* expression determined using rt-qPCR.

#### Scratch wound assay

Wounds were produced using a P200 pipette tip in a confluent monolayer of N2a cells 24 hours after transfection. Wounds were imaged at 0, 24, 48 and 72 hours, using an automated computer-controlled microscope deck images can be taken of the same area of the culture vessel at higher magnification. The area of the wound was measured with the wound healing plugin in FIJI (Variance window radius=1, Threshold value=20, Percentage of saturated pixels=0.4, Global scale=Yes and diagonal scratch= No).

#### Bromodeoxyuridine exposure assay

Mouse neuroblastoma N2a cells were exposed to Bromodeoxyuridine (BrdU) for one hour and then fixed with paraformaldehyde and stained using rabbit anti-BrdU primary antibody (Bio-Rad) followed by biotinylated goat anti-rabbit secondary before an ABC kit (Vector laboratories) and DAB (Vector laboratories) staining and a Hematoxylin counterstain.

#### Lactate dehydrogenase assay

Cells were cultured without phenol red in 12 well plates at a density of 40,000 cells, allowed to adhere for 24 hours, and then transfected with siRNA. The maximum LDH was provided by introducing triton X-100 at a 1% v/v concentration to the growth media 40 minutes prior to sample collection. Culture media samples were taken at 24, 48 and 72 hours post transfection with siRNA. 100 µl of culture media was pipetted into a 96-well plate and followed by 100 µl of LDH assay working reagent. Wells were well mixed through pipetting up and down several times. The 96-well plate was spun at 1000 RPM for 1 minute to remove bubbles from the wells. An initial measurement was taken at 490 nm. The plate was stored in a dark drawer at room temperature for 30 minutes. A second measurement was taken at 490 nm. The difference in absorption was used to calculate the LDH levels and therefore the cell stress and cytotoxicity of the siRNA mediated *Actb* knockdown.

### Mouse experiments

#### *Actb*^+/−^ mice husbandry, phenotyping- and sample collection

Mice were bred under Home Office project licence PAFFC144F and culled using Schedule 1 (S1) killing. Actb^lox^ C57BL/6 male mice (gift from Dr James Ervasti, University of Minnesota) (B6.129S6(FVB)-*Actb*^tm1.1Erv^/J; Strain #:029888; RRID:IMSR_JAX:029888, The Jackson Laboratory), were crossed with K14-Cre C57BL/6 female mice (STOCK Tg(KRT14-cre)1Amc/J Tg(KRT14-cre)1Amc/J, Strain #:004782; RRID:IMSR_JAX:004782, The Jackson Laboratory) that express Cre recombinase in the oocytes resulting in whole body deletion of *Actb* exons 2 and 3 in progenies. Mouse breeding and husbandry was performed at the University of Manchester on a 12 hour light and dark cycle with temperature regulated to 21°C (±2 °C) and humidity 40-50%, mice were kept in cages containing nesting material and wood chips. Access to food and water in sterile pouches was provided ad libitum. Enrichment opportunities of cardboard rolls, wooden blocks and cardboard housing units are provided in the cages and replaced when worn. The cages are checked daily for neonate pups at 10 am ±1 hour and are cleaned out and bedding replaced as per standard operating procedures for the animal unit. Extraction of gDNA from tail and ear snips for genotyping was performed using standard techniques and genotyping was performed using PCR (primers and PCR conditions can be provided on request). Progenies generated from crossings of male *Actb*^lox^ and female K14-Cre mice. Neonatal D0 mice were collected and sacrificed using S1 methods in the animal unit and transported on wet ice to the dissection room in the laboratory at D0, and measured, weighed and examined for anomalies in the craniofacial region, palate, and limbs. Their brains, hearts and kidney were collected, and snap frozen on dry ice and stored at −80 °C.

#### RNASeq of *Actb*^+/−^ mice brain samples

##### Sample extraction and library preparation

RNA extraction from the neonatal brain was performed by washing the weighed brain samples in ice cold PBS to remove blood contamination2 times prior to homogenisation in 600 μl of cell lysis buffer from the RNAeasy kit (Qiagen). Homogenisation was performed by 20 seconds at 5000 rpm in a Prelysis machine in homogenisation tubes containing lysing pellets (MP Biomedicals). From this step the suspension of tissue sample was then kept on wet ice and a sample volume corresponding to 30mg tissue was removed to ensure the correct amount of tissue sample was used in the RNAeasy kit columns as per the manufacturer’s instructions. The optional step of drying the membrane using centrifugation prior to RNA elution was performed and the total RNA was eluted using 2x 30 μl of RNAse free water provided in the kit.

Total RNA was submitted to the Genomic Technologies Core Facility (GTCF) at the University of Manchester. Quality and integrity of the RNA samples were assessed using a 2200 TapeStation (Agilent Technologies) and then libraries generated using the TruSeq® Stranded mRNA assay (Illumina, Inc.) according to the manufacturer’s protocol. Briefly, total RNA (0.1-4ug) was used as input material from which polyadenylated mRNA was purified using poly-T, oligo-attached, magnetic beads. The mRNA was then fragmented using divalent cations under elevated temperature and then reverse transcribed into first strand cDNA using random primers. Second strand cDNA was then synthesised using DNA Polymerase I and RNase H. Following a sin‘l’’A’ base addition, adapters were ligated to the cDNA fragments, and the products then purified and enriched by PCR to create the final cDNA library. Adapter indices were used to multiplex libraries, which were pooled prior to cluster generation using a cBot instrument. The loaded flow-cell was then paired-end sequenced (76 + 76 cycles, plus indices) on an Illumina HiSeq4000 instrument. Finally, the output data was demultiplexed (allowing one mismatch) and BCL-to-Fastq conversion performed using Illumina’s bcl2fastq software, version 2.20.0.422. Unmapped paired-end sequences from an Illumina HiSeq4000 sequencer were tested by FastQC (http://www.bioinformatics.babraham.ac.uk/projects/fastqc/). Sequence adapters were removed, and reads were quality trimmed using Trimmomatic (Bolger, Lohse, and Usadel 2014). The reads were mapped against the reference mouse genome (mm10/GRCm38) and counts per gene were calculated using annotation from GENCODE (http://www.gencodegenes.org/) using STAR (Dobin et al. 2013). Normalisation, Principal Components Analysis, and differential expression are calculated with DESeq2 (Love, Huber, and Anders 2014).

##### Bioinformatic analysis

Unmapped paired-end sequences from an Illumina HiSeq4000 sequencer were tested by FastQC (http://www.bioinformatics.babraham.ac.uk/projects/fastqc/). Sequence adapters were removed, and reads were quality trimmed using Trimmomatic_0.39(54). The reads were mapped against the reference mouse genome (mm10/GRCm38) and counts per gene were calculated using annotation from GENCODE M25 (http://www.gencodegenes.org/) using STAR_2.7.7a(47). Normalisation, Principal Components Analysis, and differential expression was calculated with DESeq2_1.36.0(55). Adjusted p-values were corrected for multiple testing (Benjamini and Hochberg method). Heatmaps were drawn with complexHeatmap v2.12.1(56). Pseudo-bulk gene expression profiles derived from mouse cerebral cortex single-cell RNA-seq data at D0 were obtained from a previous published study (Suppl. Data 1 in Loo et al(57)). From this pseudo-bulk data, an expression heatmap for our ‘genes of interest’ (identified from our bulk RNASeq experiments) was generated, and ‘cell-clusters of interest’ were identified. Next, using k-means clustering the published pseudobulk data were sub-clustered in two stages (firstly, 8 clusters of gene expression profiles were generated from the entire D0 pseudobulk data, followed by sub-clustering into 4 groups to further resolve the gene expression patterns) and the ‘marker genes’ that define our cell-clusters of interest were identified. These marker genes were highlighted in the bulk RNASeq volcano plots het versus WT.

#### qPCR validation *Actb*^+/−^ mice brain samples

Rt-qPCR was performed in triplicate using the DDCT method using the StepOne facilities provided by the genomic technologies core facility at the University of Manchester. A master mix was made containing the equivalent of 5 μl SYBR Green (Thermo Fischer scientific) 0.5 μl forward primer for the gene of interest and 0.5 μl reverse primer for the gene of interest for each reaction and a total of 6 μl of this mix was pipetted into each required well of a 96 well plate (Corning), enough cDNA from each sample at a concentration of 2ng/μl was combined with the equivalent volume of nuclease free water to provide a master mix of cDNA for each triplicate reaction and each gene tested, 4 μl of this mix was added to the relevant wells. The reaction was set to include a 95 °C hold for 10 minutes before 40 cycles of 95 °C for 30 seconds, 60 °C for 30 seconds and 72 °C for 35 seconds. A final hold at 72 °C for 10 minutes was performed prior to a melt curve analysis from 65 °C to 95 °C with 0.5 °C increments for 0.05 seconds. The resulting data was exported to excel from the software for further analysis. Data analysis was performed by the previously mentioned DDCT in which the threshold cycle (CT) is the cycle where the fluorescence of the sample reaches a set threshold, and is compared between the target gene and a reference gene. The difference in CT is compared between the target sample and a reference sample giving the DDCT value as fold change in gene expression from the reference sample (Rao et al. 2013). The housekeeping gene eukaryotic translation elongation factor 2 (eEF2) was used in rt-qPCR due to difficulties found with maintaining a steady expression level in our samples. eEF2 was chosen as it had very little change and most definitely no significant change in expression in the RNAseq data of male D0 mouse brain we produced. Furthermore studies have shown that it is a stable and reliable housekeeping gene(58).

#### BWCFF mouse models

CRISPR/Cas9 mutagenesis and genotyping was attempted in different experiments using both chemical guides (i.e. RNA) and guide- and Cas9-expressing plasmids. Detailed description is provided in Suppl. Note 7.

#### iPSC-derived neurosphere experiments

##### Reprogramming of the patient-derived primary fibroblasts to generate induced pluripotent stem cells (iPSCs)

Reprogramming of the patients’ fibroblasts and further application of the iPSC cultures for neural differentiation was reported previously(59). Briefly, two cell lines referred as mut*ACTB* or BWCFF *ACTB* Thr120Ile (heterozygous variant NM_001101.5(ACTB):c.359C>T p.Thr120Ile) and mut*ACTG1* BWCFF *ACTG1* Thr203Met (NM_001614.5(ACTG1):c.608C>T p.Thr203Met) were reprogrammed using the CytoTune-iPS 2.0 Sendai Reprogramming Kit (Thermo Fisher Scientific). iPSC line SC102A-1 (System Biosciences) was utilised as a control (c1 in Niehaus et al(59)). In this study we used the clone mu*tACTB*-1(59) and mut*ACTG1*-3. The presence of the NM_001614.5(ACTG1):c.608C>T p.Thr203Met in mut*ACTG1*-3 was confirmed by locus specific Sanger sequencing as described in Niehaus et al. All iPSC clones were tested for the removal of Sendai virus genomes and transgenes by reverse transcription-PCR analysis, and further checked for pluripotency marker expression, differentiation to three germ layers and an intact karyotype by G-banding(59). iPSC lines were cultured under feeder-free conditions on Matrigel-coated plates (Corning) in mTeSR™1 (StemCell Technologies) at 37 °C, in a humidified atmosphere of 5% CO^2^ and 95% air. Medium was changed daily and cells were passaged every third day, using ReLeSR™ (StemCell Technologies)(59).

##### Neural differentiation of the iPSCs

BWCFF *ACTB* Thr120Ile, BWCFF *ACTG1* Thr203Met as well as control iPSC line SC102A-1 were differentiated using a STEMdiff™ SMADi Neural Induction Medium according to the manufacturer protocol. Briefly, iPSC colonies were detached with TrypLE™ Express (Gibco) to obtain a single cell suspension. 10.000 cells were seeded into each well of a 96-well Clear Round Bottom Ultra-Low Attachment Microplate to form embryoid bodies (EBs). After 5 days EBs were re-plated onto a Matrigel™ (Corning) coated 24-well plate with STEMdiff™ SMADi Neural Induction Medium. During the next days of incubation, neural rosettes formed within EBs showing characteristic morphology. On day 11 or 12 neural rosettes were isolated using STEMdiff™ Neural Rosette Selection Reagent and transferred to one well of a 6-well plate with STEMdiff™ SMADi Neural Induction Medium. During the following days, cells migrated out of the neural rosettes forming a monolayer of neural progenitor cells (NPCs). After the monolayer culture reached 95-100% confluency, NPC were passaged into the 6-well plate with the STEMdiff™ Neural Progenitor Medium where NPCs were maintained in culture till the next experiments. NPC identity was verified by expression of neuronal progenitor markers SOX2 and Nestin (data not shown).

##### Neurosphere migration assay

Neuronal migration was assessed using a modified neurosphere outgrowth assay previously described by Marchetto et al.(60). 5000 NPCs cells were seeded into each well of a 96-well Clear Round Bottom Ultra-Low Attachment plate and incubated for 5 days to form neurospheres in STEMdiff™ Neural Progenitor Medium. On day 5 these neurospheres were replated into the center of a Matrigel™ coated 24-well plate using a pipette with wide bore tips. The plate was carefully placed into the Incucyte® S3 Life-Cell Analysis System and phase contrast images were taken every 4 hours for 72 hours. In total 4 neurospheres were analysed per cell line and migration assay was performed three times. Phase contrast images taken by Incucyte® S3 Life-Cell Analysis System were analysed using Image J software (NIH) where averaged NPC migration distance from each neurosphere was measured and normalised to the cell core area at the begin of the assay. Evaluation strategy is visualised in Figure 6B.

##### Immunocytochemistry of the neurospheres

After 72 hours, neurospheres were fixed with 1% PFA for 30 min and washed with PBS for three times (3 neurospheres per cell line). Cells were permeabilized with ice cold methanol for 5 min and blocked for 1 hour in the blocking buffer (2% FBS in PBS). Primary antibody incubation of Tuj1, Ki-67, SOX2 (Suppl. Table 4) was then performed for 1 hour at room temperature and secondary antibody incubation as well as DAPI staining for 30 min also at room temperature. Neurospheres were washed with PBS for three times and then mounted in Moviol® 4-88. Samples were stored at 4 °C in darkness until further imaging. Stained neurospheres were imaged using confocal microscopy. Images were acquired with the 10x objective in a Zeiss LSM 880 Airy upright. Stitching and maximum intensity z-projection was performed using ZEN black software (Zeiss). Image processing was performed using Image J software (NIH, Bethesda, ML, US).

## Author contributions

Conceptualization: NDD, DM, ASW, SB

Methodology: NDD, DM, ASW, SB, AR, KG, GM, DM, ES, MHT, PK, LZ, MH Investigation: AT, JNG, MK, JC, SC, SC, BC, HC, MC, SC, PhD, KE, AEF, LC, SH, WGJ, MK, IK, KK, PMR, AM, IN, IR, FSS, AM, KT, ST, JV, AV, BW, TCH, MS, Supervision: NDD, AR, KG, ES, MHT, PK, LZ, MH, CBL, NR, DM, ASW, SB

Writing - original draft: NDD, DM, ASW, SB Writing - review & editing: all coauthors

## Supporting information

Supplemental moviews

## Acknowledgments

N.DD, Ph.D, M.K., and I.N. gratefully acknowledge support of the Core Facility at the NCT/UCC-CMTD Dresden and Stem Cell Engineering Core Facility of the CMCB Technology Platform at TU Dresden.

D.J.M. and his lab gratefully acknowledge support provided by the Research Core Unit for Structural Biochemistry. Computing time was provided on supercomputers Lise and Emmy at NHR@ZIB and NHR@Göttingen, as part of the Alliance for National High-Performance Computing (NHR) infrastructure. The calculations for this research were conducted with computing resources under the project ID nib00018.

