## Supplemental moviews for "Genomic and biological panoramas of non-muscle actinopathies"

### Slide 1
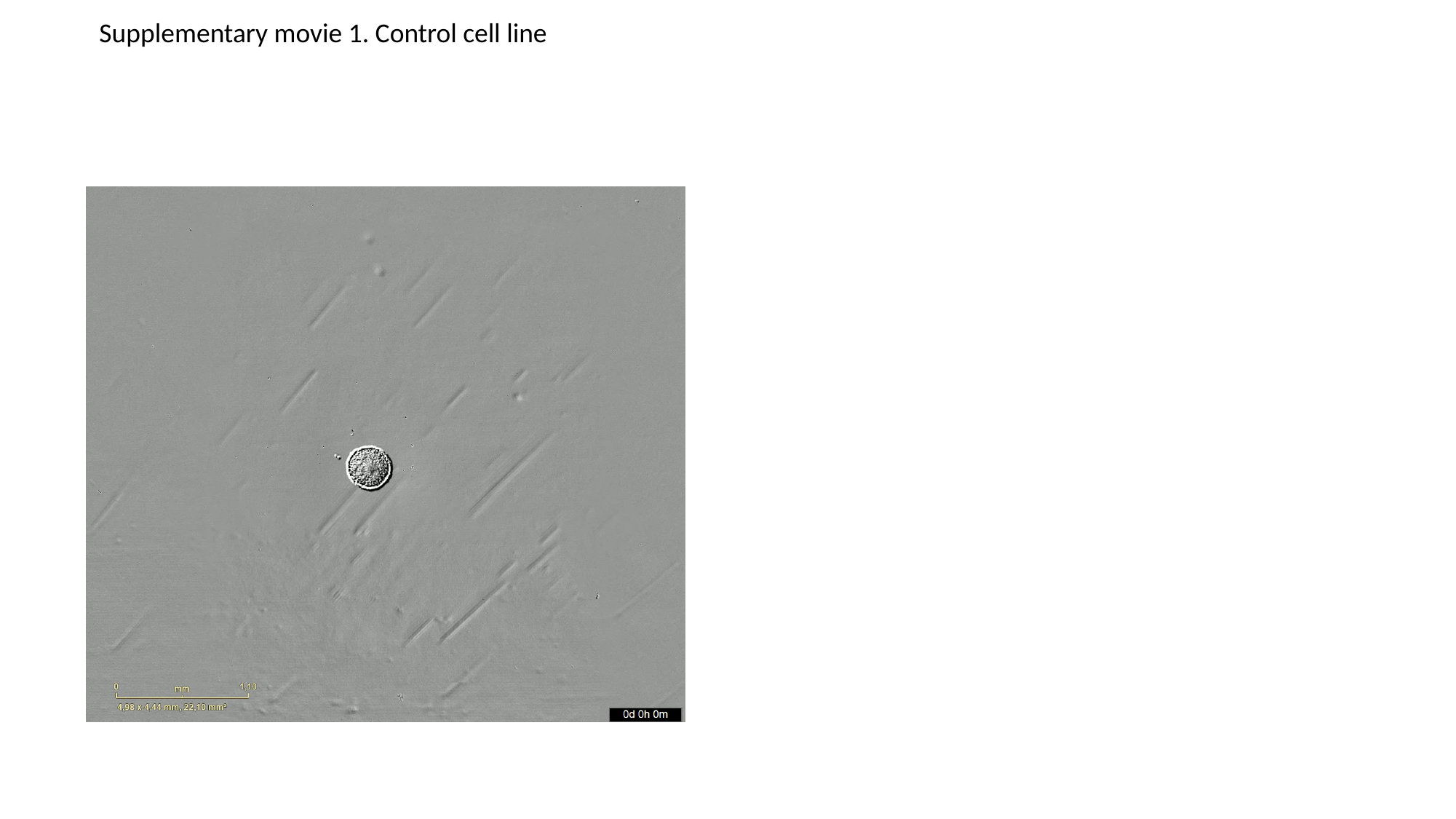

Supplementary movie 1. Control cell line

### Slide 2
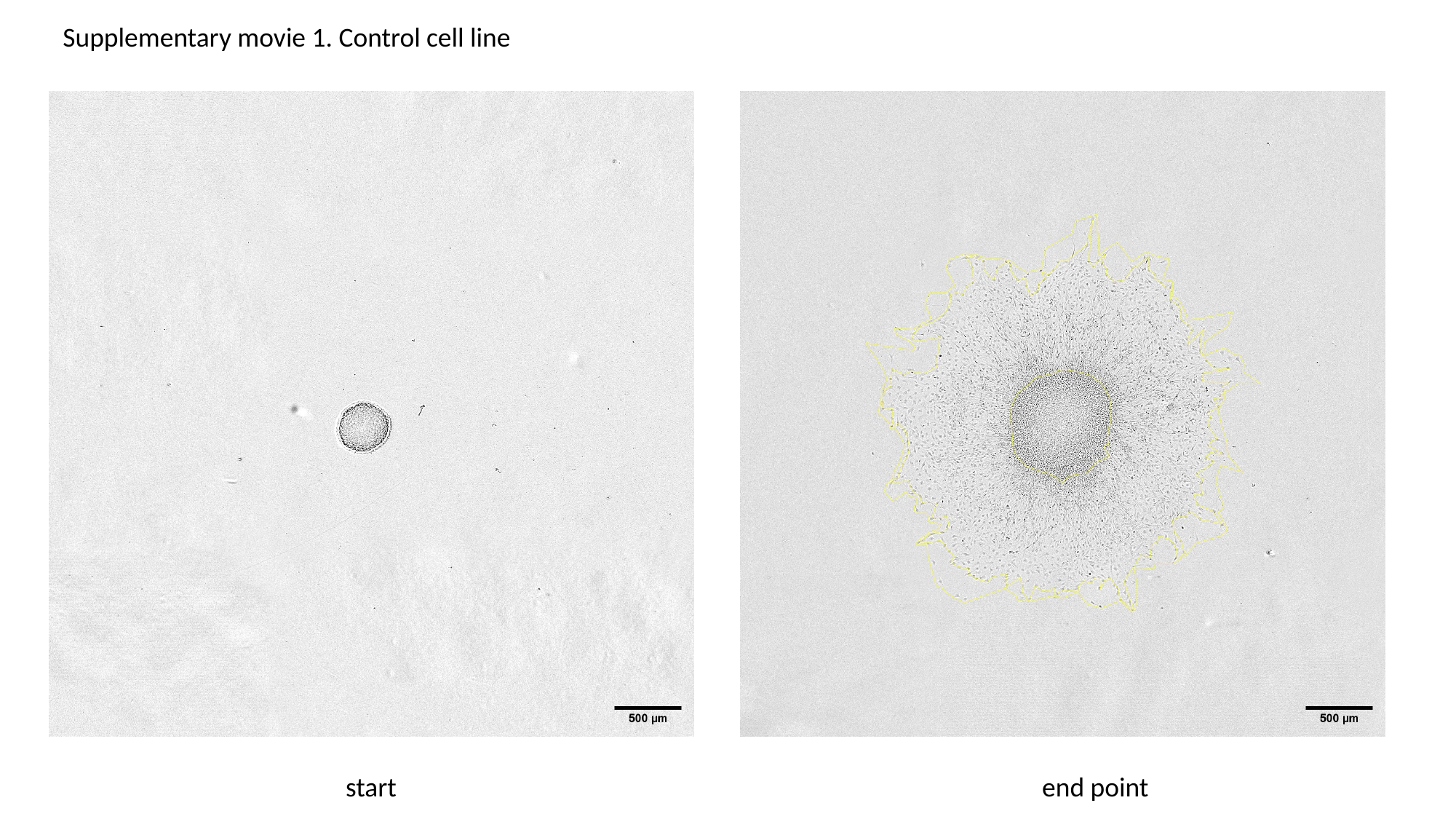

Supplementary movie 1. Control cell line
start
end point

### Slide 3
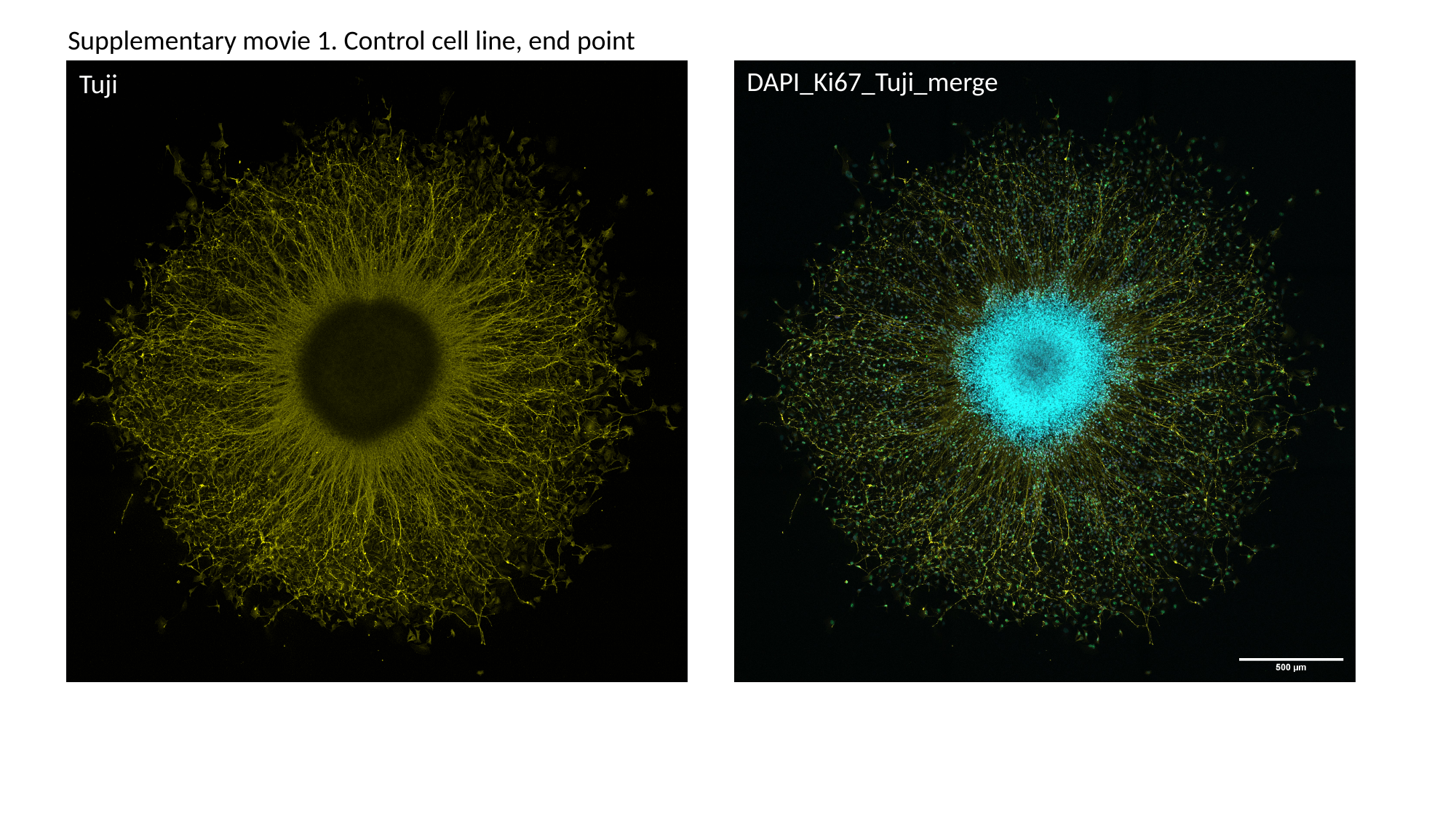

Supplementary movie 1. Control cell line, end point
DAPI_Ki67_Tuji_merge
Tuji

### Slide 4
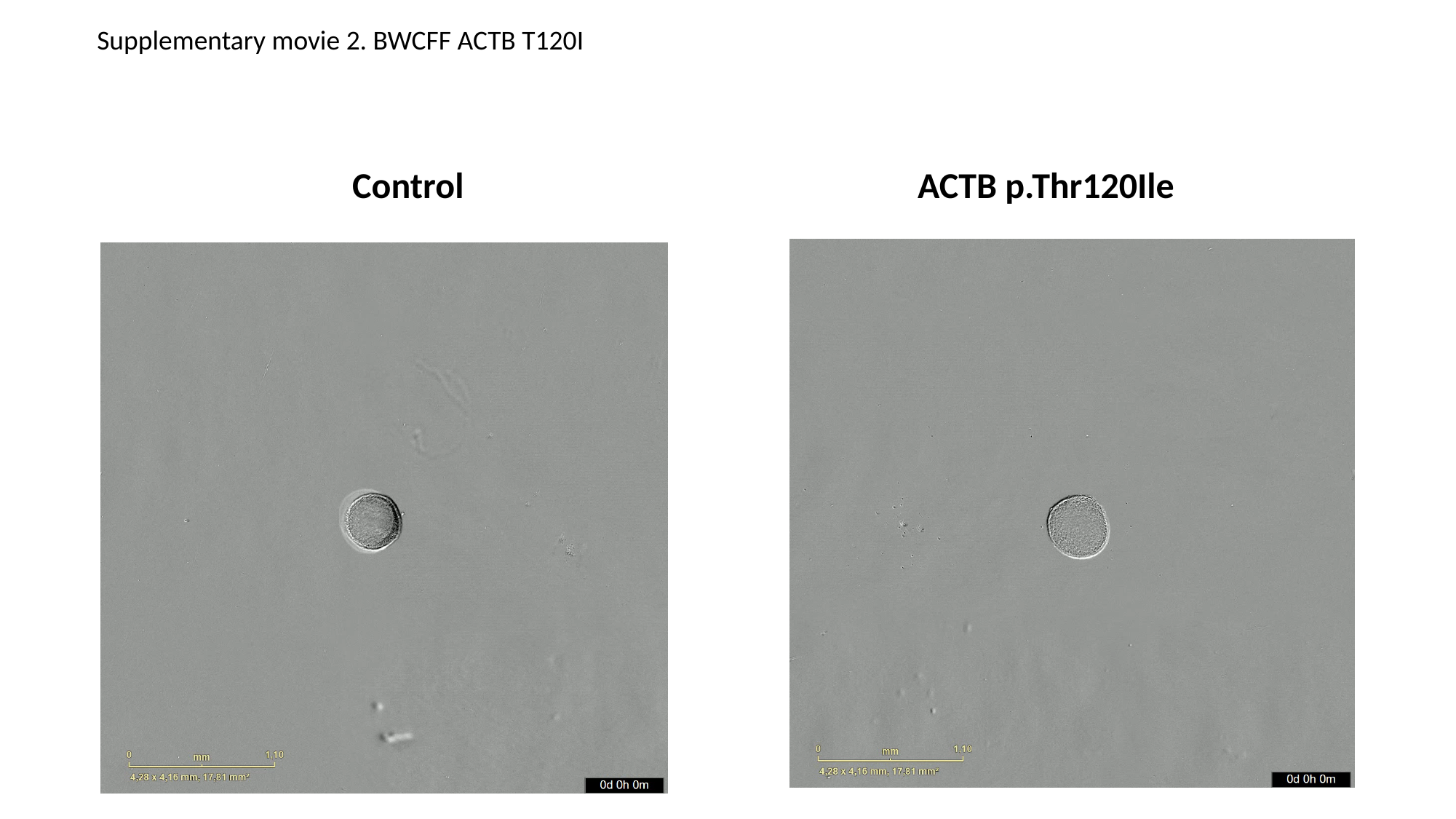

Supplementary movie 2. BWCFF ACTB T120I
Control
ACTB p.Thr120Ile

### Slide 5
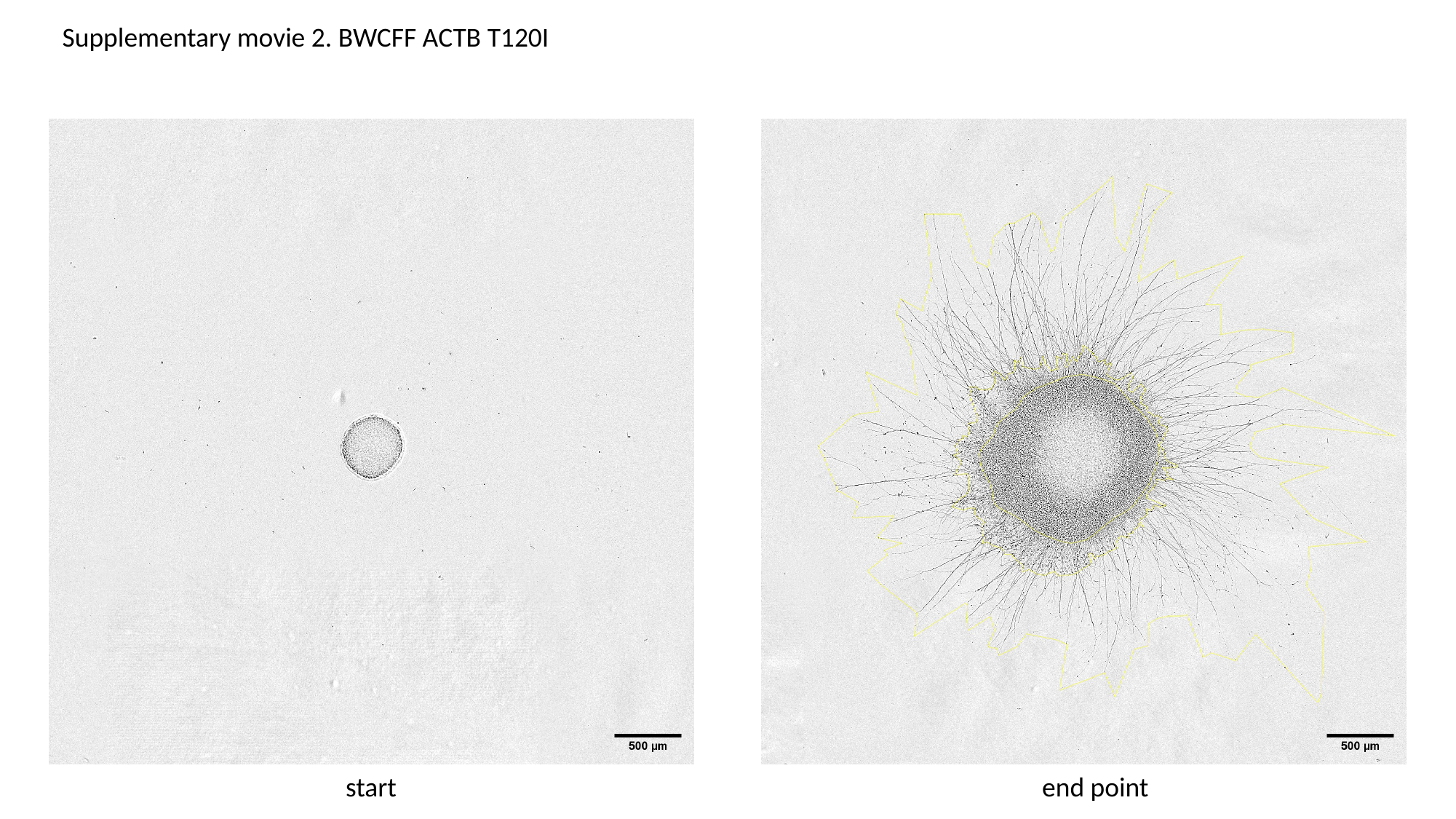

Supplementary movie 2. BWCFF ACTB T120I
start
end point

### Slide 6
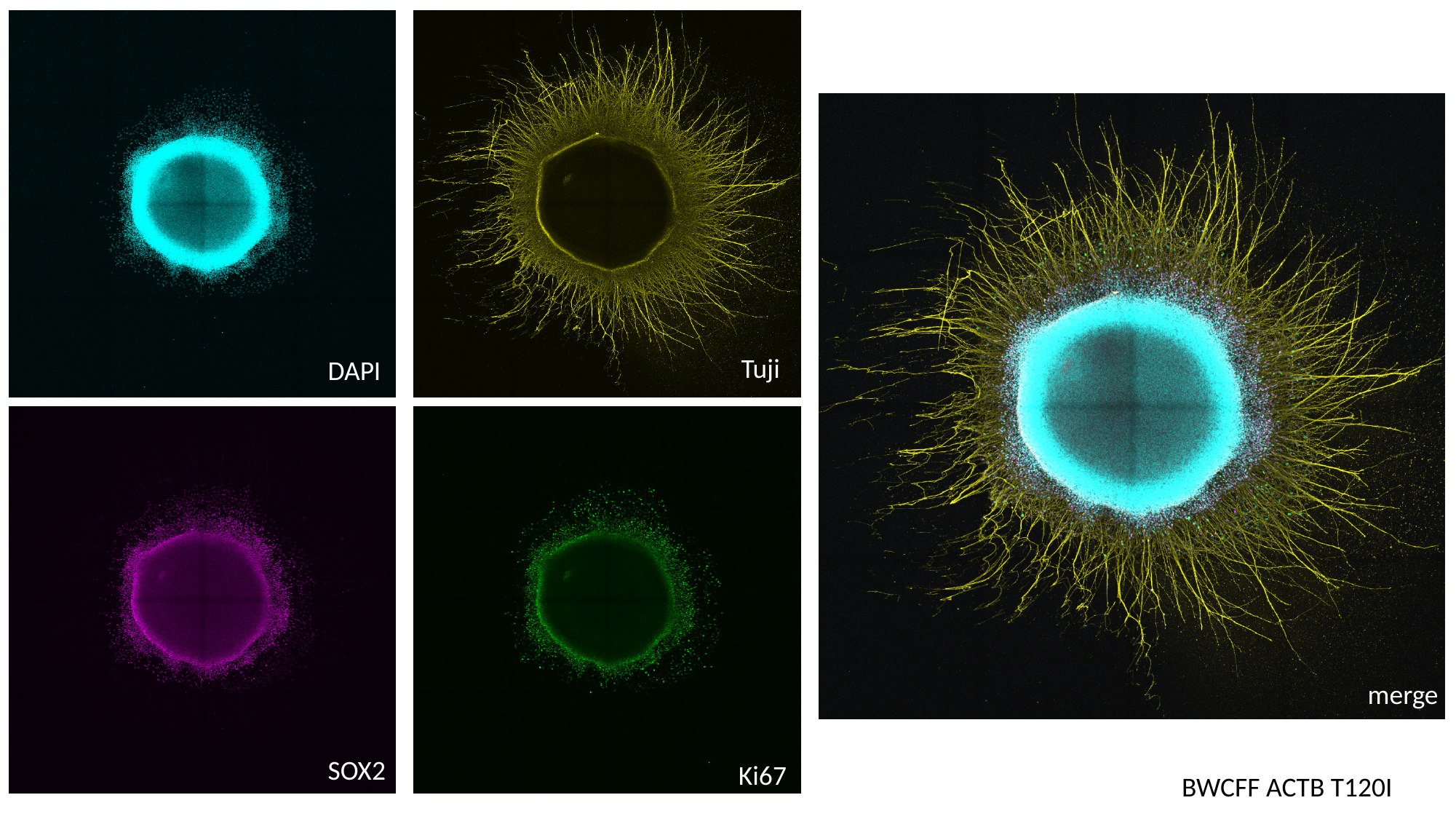

Tuji
DAPI
merge
SOX2
Ki67
BWCFF ACTB T120I

### Slide 7
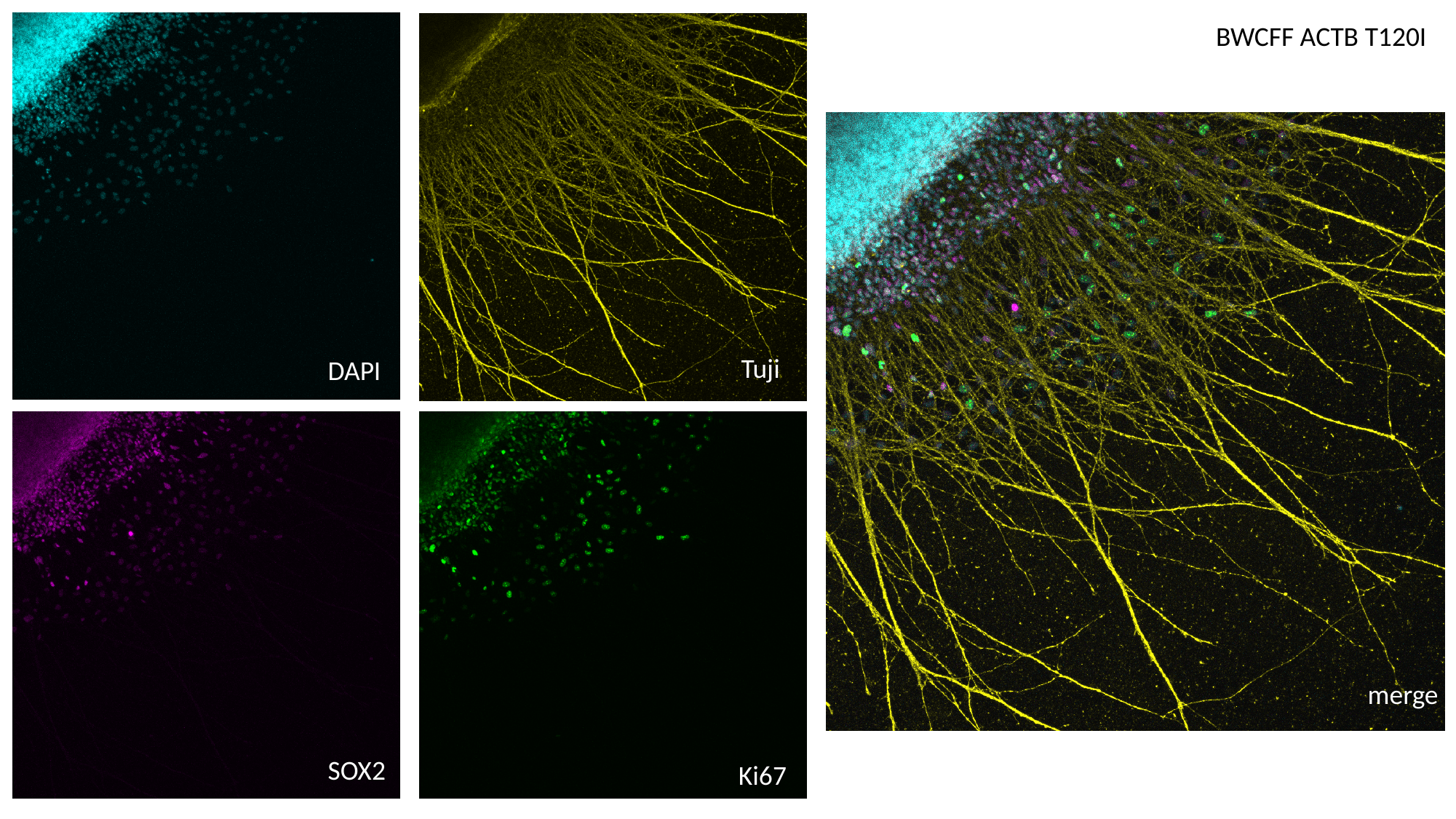

BWCFF ACTB T120I
Tuji
DAPI
merge
SOX2
Ki67

### Slide 8
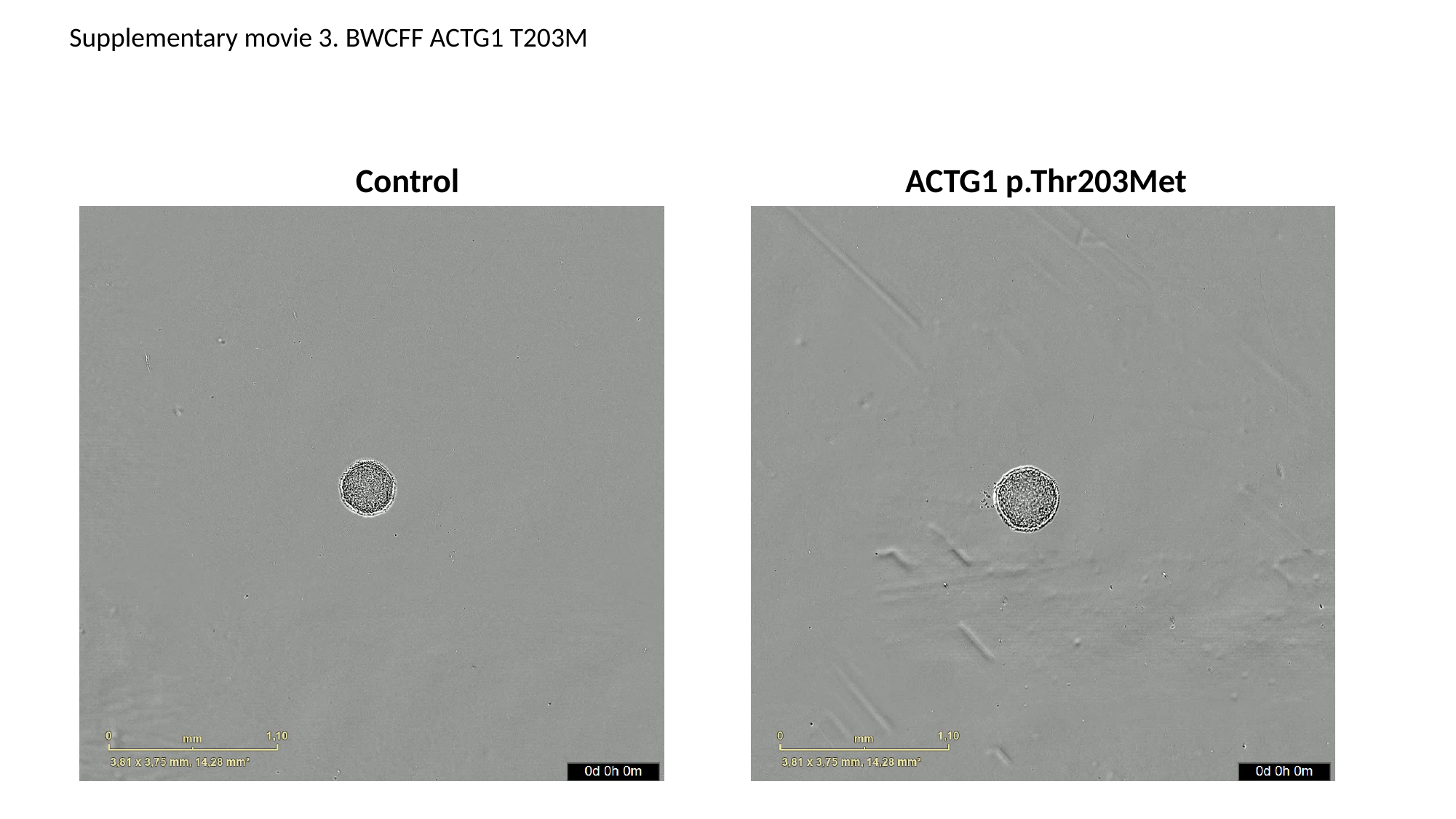

Supplementary movie 3. BWCFF ACTG1 T203M
Control
ACTG1 p.Thr203Met

### Slide 9
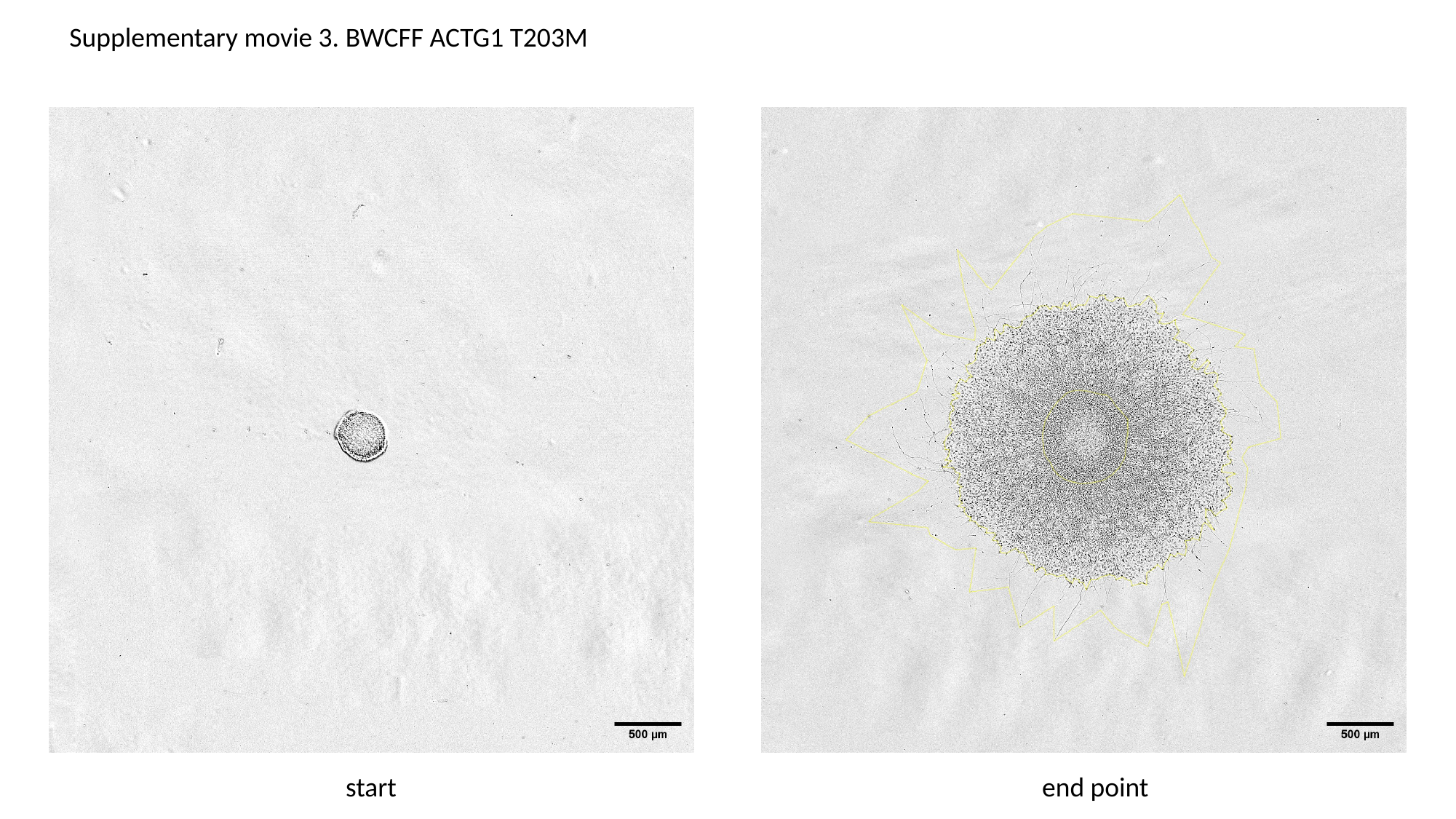

Supplementary movie 3. BWCFF ACTG1 T203M
start
end point

### Slide 10
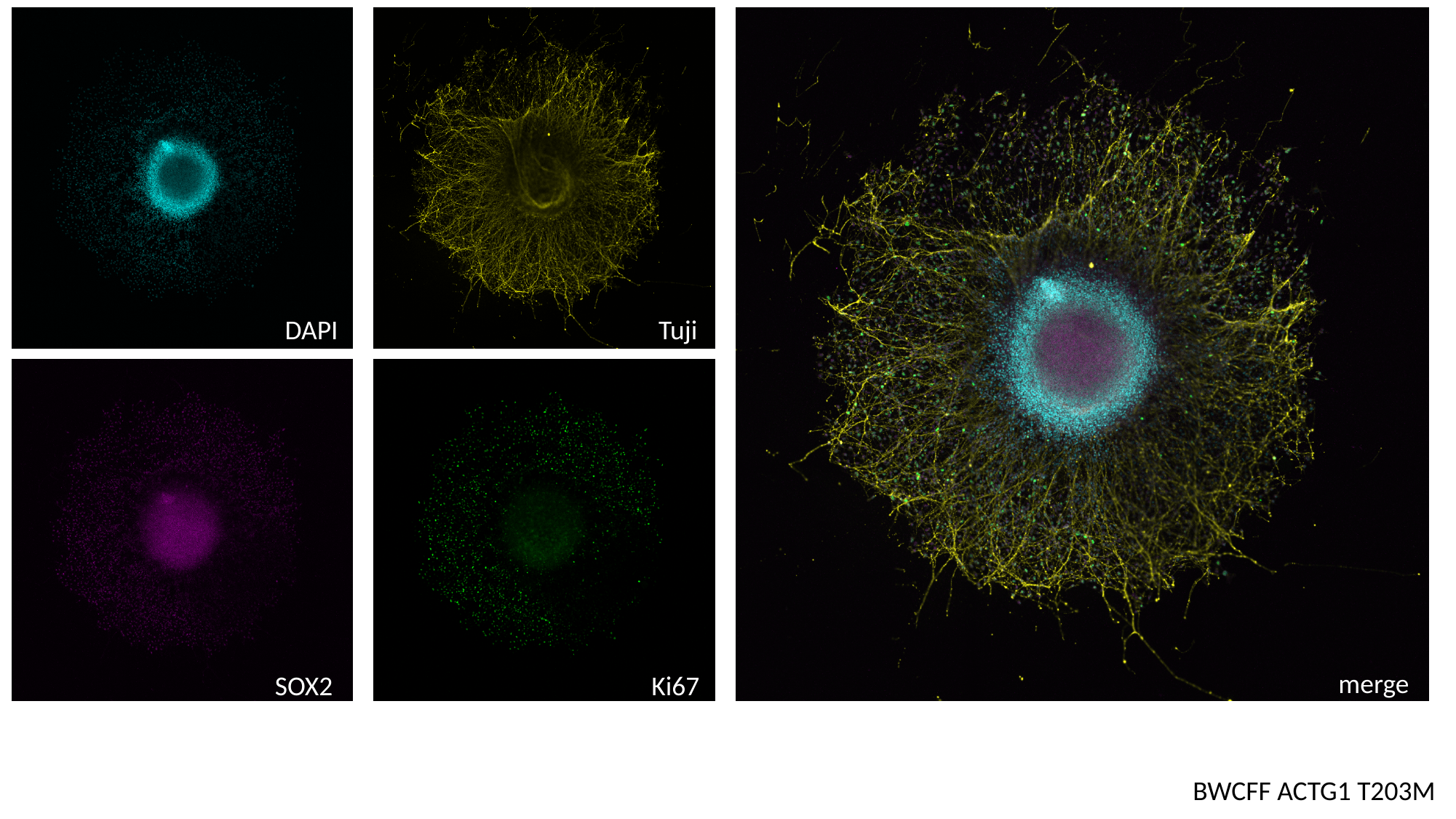

Tuji
DAPI
merge
SOX2
Ki67
BWCFF ACTG1 T203M

### Slide 11
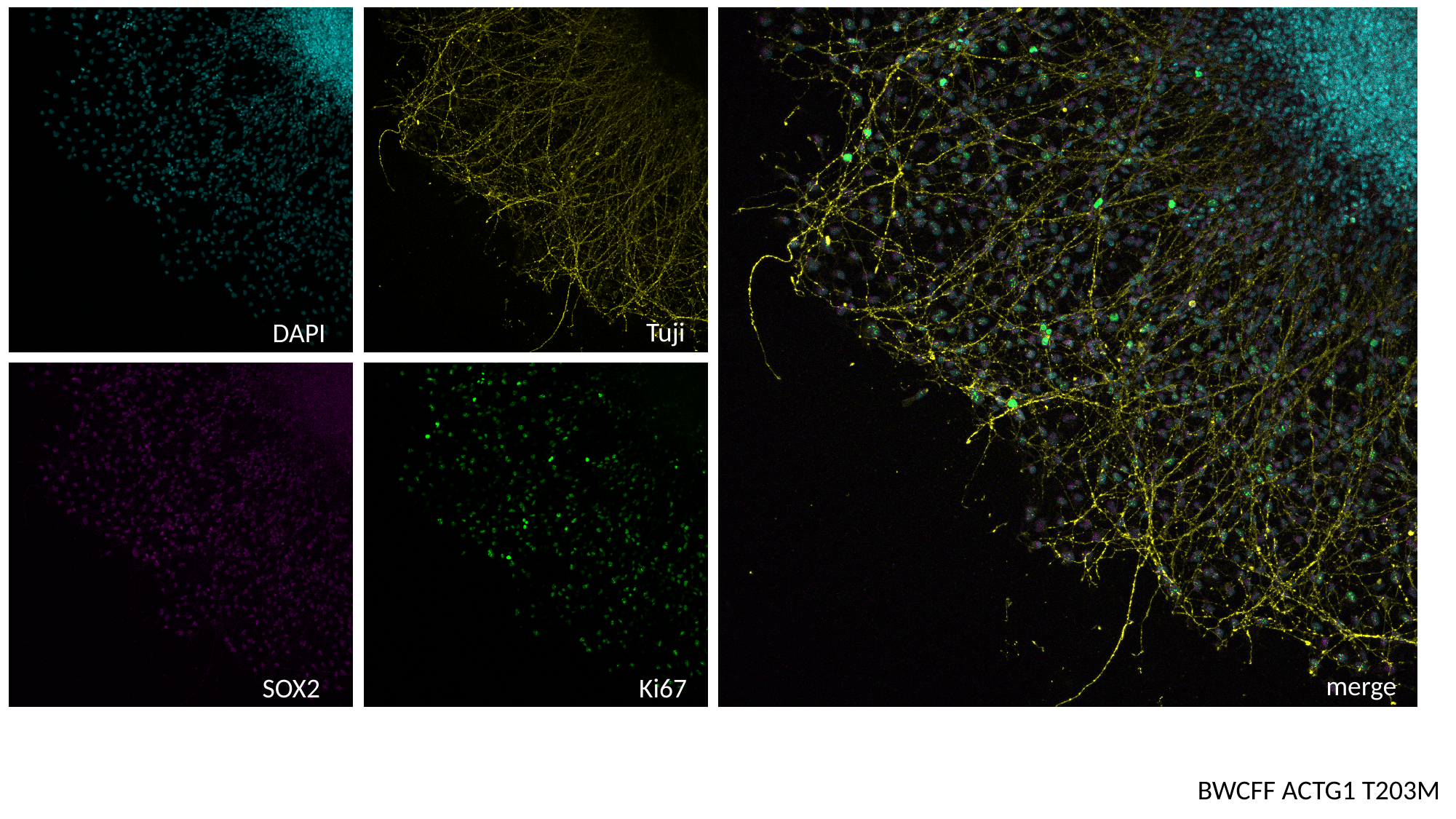

Tuji
DAPI
merge
SOX2
Ki67
BWCFF ACTG1 T203M
