## Supplementary material for "Genomic and biological panoramas of non-muscle actinopathies"

### Supplementary Material for
### Genomic and biological panoramas of non-muscle actinopathies

**SUPPLEMENTAL NOTE S1: SYNDROME CAUSED BY ACTB pLoF VARIANTS**

This note includes details about the syndrome cause by heterozygous germline predicted loss of function (pLOF) variants (nonsense and frameshift), and missense variants (MVs) resulting in instability of cytoplasmic β-actin (β-CYA).

0% Absent, <10% Rare, 10-25% Sometimes, 25-75% Frequent, >75% Very frequent
100% Always

Tabular summary of clinical features of individuals with *ACTB* pLOF variants (N=31*)

| | |
|---|---|
| **Intellectual development and behaviour problems** (Frequent) | 21 had intellectual difficulties (ID), mostly borderline/mild and moderate (in two). Where information was available, individuals without ID had either normal or low normal IQ (75-80). Generally, individuals had open and pleasant personalities. Behaviour anomalies reported in 14/31 (45%) individuals (with or without ID), and included attention deficit hyperactivity disorder (ADHD), temper tantrums, autism and difficulty in socialising. |
| **Craniofacial Anomalies** (Very frequent) | Recognisable facial features with long face, straight eyebrows, deep set eyes, epicanthus, narrow or flat nasal bridge with broad nasal tip, and large mouth. Microcephaly was present in 13. |
| **Eye coloboma** (Rare) | Iris coloboma and cataract in one individual (116-B). |
| **MRI anomalies** (Frequent) | Reported in 5/14 (36%) and included periventricular nodular heterotopia (PNVH) in three, hypoplastic corpus callosum (CC) and hypoplastic cerebellar vermis in one, and unspecific areas of high T2 signal in one. Pachygyria was not reported in anyone. MRI was not performed in 17/31. |
| **Growth problems** (Frequent) | Short stature was documented in 11 (up to -4 SD). |
| **Epilepsy** (Absent) **or Seizures** (Rare) | No individual was reported to have epilepsy. Seizures were reported in two and included as single episodes of seizures during the early childhood with spontaneous remission later (in individual 168-B with additional variant in *MID2* and individual 107-B). |
| **Dystonia** (Rare or absent) | Reported in one previously published individual (63-B, patient XXIV(1)) who was lost for follow-up. No neurological features noted in other individuals (included five in the $4^{th}$ decade an older). |
| **Hearing loss** (Rare) | Reported in three and included bilateral sensorineural (173-B, XXVI(1)), bilateral mixed (153-B, XXII(1)), or conductive that resolved after the first few years of life (111-B). |
| **Skeletal anomalies** (Frequent) | Reported in nine and included pectus deformities, scapula winging, leg deformities, craniosynostosis, congenital parietal foramina, and scoliosis. |
| **Heart Defects** (Frequent) | Congenital heart defects reported in eight individuals and included atrial septal defect (ASD), ventricular septal defect (VSD), pulmonary stenosis, and patent ductus arteriosus. One individual reported with cardiomyopathy (remission at the $2^{nd}$ decade). One individual reported with left ventricular dilatation without functional consequences. |
| **Respiratory problems** (Sometimes) | Reported in five and included severe and prolonged respiratory infections and pneumonias in three and asthma in the remaining two |
| **Gastro-intestinal problems** (Frequent) | Reported in 12 and included feeding difficulties, failure to thrive in early childhood, constipation, gastro-oesophageal reflux, esophageal atresia with trachea-oesophageal fistula (in one) and gallstones (in one). |

| | |
|---|---|
| **Genito-urinary anomalies** (Sometimes) | Reported in five and included horseshoe kidney in three, hypospadias and renal cortical cysts. |
| **Skin and integument** (Rare) | Atopic reactions (in three), sparse scalp hair, generalised hirsutism, facial haemangioma, extra skin folds on abdomen and back. |
| **Repeated infections** (Frequent) | Reported in 8 and included recurrent respiratory infections including pneumonias, and multiple acute otitis media, chronic ear infections. Tests of immune function were performed in one and no abnormality was detected. |
| **Haematological anomalies** (Sometimes) | Thrombocytopenia was documented in 6 individuals from the original report of the ACTB-associated syndromic thrombocytopenia(2), one individual with *ACTB* gene deletion (111-B) and one patient with an *ACTB* MV (61-B). Blood counts were not available for 4/31. |
| **Healthy carriers** (Rare) | All carries demonstrated typical features that however might have been limited to mild craniofacial dysmorphism and learning difficulties mentioned only on enquiry after the molecular diagnosis |

*Some features could not be assessed in all individuals.

Diagnostic and follow-up recommendations

- Individuals with larger deletions can have more severe presentation, perhaps due to loss of other genes.
- 6 out of 31 patients inherited the ACTB pLOF variant from a parent who was similarly affected, or mildly affected, or apparently unaffected. pLOF *ACTB* variants should, therefore, be considered as pathogenic variants even if inherited from apparently unaffected parent.
- Clinical follow-up should consider appropriate nutritional status in infancy and early childhood; screening for heart and renal defects; regular hearing test in early childhood in patients with recurrent otitis; singular blood count with occasional follow-up if necessary (transitory thrombocytopenia with spontaneous remission during first decade was reported); GI function monitoring; developmental and behaviour assessment with appropriate intervention (patients with severe behaviour anomalies benefit from symptomatic medication).

#### SUPPLEMENTAL NOTE S2: BARAITSER-WINTER-CEREBROFRONTOFACIAL SYNDROME

The first report(3) of the syndromic condition later named as Baraitser-Winter syndrome described three children with ID and a unique combination of clinical features including iris coloboma, bilateral ptosis, telecanthus, hypertelorism and short stature, the gestalt resembling Noonan syndrome(4). The major clinical features were further extended with trigonocephaly and/or prominent metopica suture and lissencephaly mostly in form of frontal predominant pachygyria with or without posterior subcortical band heterotopia(5, 6). Delineation of the genetic cause(7) demonstrated that two other syndromes originally described as separate conditions (Fryns-Aftimos and cerebrofrontofacial syndromes) were part of the same spectrum with a unifying name of the Baraitser-Winter-Cerebrofrontofacial syndrome (BWCFFS) (8, 9).

Tabular summary of clinical features in patients with BWCFFS (N=113)

| | |
|---|---|
| **Intellectual development and behaviour** (Nearly always) | 101/104 individuals whose developmental level could be assessed, presented with developmental delay (DD) (N=25, age 9mo-3y) or ID, that was mild (in 24), moderate (in 33), severe (in 21) and profound in 6 individuals. For 17 individuals ID grade was not specified. Remaining 5 individuals were younger than 9 months; 4 fetal cases were also excluded from this evaluation. One of three individuals without ID had IQ>130. 14 individuals with ID showed behaviour anomalies that included ADHD, hyperactivity, temper tantrums with aggression and autism; however, majority of the patients had open and pleasant personalities. |
| **Craniofacial Anomalies** (Nearly always) | Craniofacial anomalies were very consistent resulting in distinct and recognizable facial gestalt. Typical features included prominent metopic ridge, hypertelorism, high-arched eyebrows, ptosis, long palpebral fissures with everted lower lid, broad nasal tip, long smooth philtrum, large mouth with thin upper lip and everted lower lip, grooved chin, large (narrow) ears [need to describe properly], low posterior hair line. Face becomes coarser in the 2$^{nd}$ decade, so gestalt is easier to recognize. Microcephaly was reported in 52% (59/113) patients with HC in range of -2 to -5.6 SD. Craniosynostosis requiring surgical correction was documented in 5 patients. |
| **Eye coloboma** (Frequent) | Iris and/or chorioretinal colobomas were reported in 28. However, vision problems and other eye anomalies were seen in 43 individuals represented by reduced vision, refractive errors and in some patients microphthalmia/microcornea, nystagmus, cataract and bilateral congenital fibrosis of the rectus medial and inferior extraocular muscles seen in one patient. |
| **MRI anomalies** (Very frequent) | MRI anomalies were documented in 81/97 (84%) individuals and were represented by lissencephaly/pachygyria in 52. Other cortical malformations included dysgyria/polymicrogyria in five, and periventricular nodular heterotopia (PNVH) in two individuals. Other anomalies included agenesis or hypoplastic CC, leukomalacia, Chiari I anomaly, hypoplastic cerebellum, and ventriculomegaly. MRI was not performed in 16 individuals. |
| **Growth** (Frequent) | Short stature was documented in 39 (height -6,4 to 3,4 SD) One individual had significantly delayed bone age and received growth hormone therapy with some catch up growth (23-B). Significant failure to thrive in early childhood was reported in at least 6 patients |
| **Epilepsy** (Frequent) | Epilepsy was present in 41, with age of onset ranging from neonatal period to 24y. Cortical malformations were reported in 28 individuals with epilepsy, five were reported not to have structural brain anomaly, and in four individuals brain imaging was not performed. |
| **Dystonia** (Absent) | Not reported. |

| | |
|---|---|
| **Hearing loss** (Frequent) | Reported in 35 patients and included bilateral sensorineural (in 19), conductive (in two) or unspecified in the remaining individuals. |
| **Skeletal anomalies** (Frequent) | Reported in 55 and included spine anomalies in 22, pectus anomalies, scapula winging, hip dysplasia, Polydactyly (in seven) and feet deformities. |
| **Heart Defects** (Frequent) | Reported in 39 and included ASD, VSD, pulmonary stenosis, patent ductus arteriosus, aortic coarctation, and valve anomalies. |
| **Respiratory problems** (Sometimes) | Reported in 14 and included severe and prolong respiratory infections and pneumonias in six patients, sleep apnoea and asthma in two patients as well as laryngomalacia and narrow nasal passage in one patient, respectively. |
| **GI problems** (Frequent) | Reported documented in 27 and included constipation, feeding difficulties (four required gastrostomy feeding), structural anomalies (in four individuals including duodenal atresia, jejunal atresia, intestinal malrotation and partial bowel obstruction). One individual had progressive liver cirrhosis, and another presented with chronic diarrhoea. Interestingly, GI complains were not documented in the first BWCFF cohort(8) and were often reported only on enquiry and no detailed information was available for 44 of 109 patients (4 fetal cases excluded). |
| **Genito-urinary anomalies** (Sometimes) | Structural renal anomalies were reported in 23 individuals and included 10 with duplicated kidneys and/or collecting system, 7 with severe hydronephrosis, and two individuals each with ectopic kidneys, renal fusion or hypoplastic kidneys. Abnormal external genitalia were described predominantly in males including cryptorchidism, inguinal hernia and small penis. Hypoplastic external genitalia were also reported in one female individual. |
| **Skin and integument** (Sometimes) | Reported in 19 patients, included dysplastic skin derivatives in 7 patients (sparse hair, hypoplastic nails, hypodontia, small teeth and delayed tooth eruption), cutis hyperelastica, vascular anomalies (cutis marmorata, teleangiaectasia, hemangiomas) and skin hyperpigmentation (café au lait marks and Mongolian sacral spot), dermatitis, pterygia and interdigital webbing reported in one or two patients each. |
| **Repeated infections** (Sometimes) | Repeated and/or excessive infections were documented in 13 patients mostly as recurrent respiratory infections including pneumonias, multiple acute otitis media as well as chronic ear infections, urinary tract infections and necrotising enterocolitis. |
| **Haematological anomalies** (Absent) | Not reported (including thrombocytopenia). |
| **Other** | We could confirm previous observation about BWCFFs typical body posture than becomes apparent in the second or third life decade(8). This includes anteverted shoulders with generally narrow shoulder gridle, scoliosis and semiflexed knees. Children are often present with excessive nuchal skinfolds or pterygium colli, low posterior hairline, pectus excavatum and mild diastasis recti resulting in prominent navel or umbilical hernia. |
| **Healthy carrier** (Almost absent) | All carries demonstrated typical features and vast majority of the patients had *de novo* variants. Only one patient inherited the pathogenic variant from affected mother(9), who presented mild but typical gestalt and had low normal intelligence. |

*Some features could not be assessed in all individuals.

#### BWCFF specific brain anomalies

BWCFF is associated with a specific MCD pattern: frontal-predominant pachygyria, frontal pachygyria accompanied with a thin occipital band heterotopia and PVNH (Suppl. Figure S21). As bilateral PVNH were also observed in patients with *ACTB* pLOF disorder, we did not consider the later MCD to be BWCFF specific. Enlarged (prominent) perivascular spaces in the centrum semiovale (but not in basal ganglia) were noted in several BWCFF patients with and without cortical malformations (N=18). As this is a quite common non-specific finding, we suspect that enlargement of perivascular spaces might have been overlooked or not mentioned in the final radiological report in patients where no MRI images were available for the evaluation.

#### Prenatal manifestation of BWCFFs

43 of 76 BWCFFs patients with available pregnancy data had abnormal prenatal history. For the remaining 37 patients, early clinical information was not available. 3 pregnancies were terminated between 26$^{th}$ and 35$^{th}$ gestational weeks. The most common manifestation was increased nuchal translucency (N=22) either transient or persisting and reaching the form of cystic hygroma (N=7). 14 patients presented with hydrops fetalis. Other recurrent features were microcephaly, agenesis of the CC, ventriculomegaly or hydrocephalus, polyhydramnios, cleft lip/palate, cortical anomalies, structural renal and heart anomalies. Reduced foetal movements, oligohydramnios and duodenal atresia were reported in a single patient respectively. Although common, prenatal manifestation is not specific and does not allow for clinical suspicion of BWCFF in absence of family history. Prenatal diagnosis is only possible through exome/genome-wide genetic testing. We recommend a very careful consideration of the clinical diagnosis in every patient with a novel MV in *ACTB* and *ACTG1* as well as MVs that were previously observed in less than three patients and/or MVs with insufficient clinical information.

#### Adult complications and reduced life expectancy in BWCFFs

Our BWCFFs cohort includes 19 individuals at the age 18-45 years. The oldest known patient was older than 60y at the time of the last follow-up (personal experience of Allan Bayat, limited clinical data were available). 16/19 patients had epilepsy with AO from 2-24y and all 19 patients showed ID ranging from mild (2 patients), moderate (N=7) to severe (N=10). 10/19 patients demonstrated progressive spinal deformity, limited extension of large joints and slow decline in overall motor activity. Two previously reported patients died during the 3$^{rd}$ life decade (10)(8) from complications of the acute ileus and progressive feeding difficulties resulting in recurrent respiratory pneumonias. Both patients carried the same MV p.Thr120Ile in *ACTB*. The whole BWCFFs cohort encompasses two other deceased individuals reported in the literature with presumable cause of death being adverse reaction to codeine administration as a young adult (10) and progressive sepsis in a neonate with untreated decompensated heart defect(11). The current adult cohort might not be representative for milder affected individuals that were currently diagnosed via genotype-first approach.

#### Diagnostic and follow-up recommendations

- BWCFF diagnostic criteria delineated in this work - (1) specific facial dysmorphism, and/or (2) frontal predominant pachygyria with or without other brain malformations or minor anomalies in a patient with (3) (likely) pathogenic MV in *ACTB* or *ACTG1*.
- Clinical follow-up includes initial organ screening (brain MRI, EEG, heart ultrasound, abdominal and renal ultrasound, assess of the nutritional status, ophthalmologic evaluation including fundoscopy, audiologic evaluation, developmental assessment and genetic counseling) and annual surveillance (surveillance might be more frequent depending on the individual situation)(12)

**SUPPLEMENTARY NOTE S3: *ACTB*:p.Arg183Trp-RELATED DYSTONIA-DEAFNESS SYNDROME**

Our cohort included 9 individuals with well documented progressive generalized dystonia; all of them carried an identical pathogenic variant in *ACTB*:p.Arg183Trp. All patients had a history of the profound prelingual sensorineural hearing loss with significant improvement with the cochlear implants in patients who received them. Four remaining individuals were ascertained following detection of *ACTB*:p.Arg183Trp and presence of the congenital deafness without BWCFF specific features. Dystonia was a fully penetrant feature in all adults. However, this might represent an ascertainment bias as all adult patients underwent genetic testing because of dystonia, whereas younger individuals received exome sequencing because of congenital hearing loss. As the cohort remains small, the exact penetrance of dystonia cannot be estimated.

First patients reported with *ACTB*-DDs were monozygotic twins(13, 14) that were subsequently discussed as a part of BWCFF spectrum(8).

It remains currently unknown whether other MV within the same *ACTB* codon would also result in *ACTB*-DDs. In ClinVar we identified one individual with a de novo MV NM_001101.5(*ACTB*):c.547C>G (p.Arg183Gly). This variant was evaluated by a single submitter as a likely pathogenic for *ACTB*-related disorder. No clinical data could be provided after our active enquiry.

Tabular summary of clinical features in patients with deafness-dystonia syndrome (N=13)

| | |
|---|---|
| **Intellectual development and behaviour** | Borderline/mild ID was present in 5 out of 13 patients, however two patients with normal intelligence had delayed motor and/or speech development. Three patients demonstrated abnormal behaviour with anxiety and insecurity in two patients and psychotic episodes with paranoid delusions in another one. |
| **Craniofacial anomalies** | No specific gestalt has been documented, however 6 patients had hypertelorism with arched eyebrows and/or mild ptosis. None of the patients had BWCFF facial gestalt. |
| **Eye coloboma** | No coloboma was reported, one patient had cataract diagnosed in childhood (13) |
| **MRI anomalies** | MRI anomalies were present in two patients and included arterial ischemic stroke during childhood in one subject (26-B) and bilateral symmetrical FLAIR T2 hyperintensity in the basal ganglia(15) in the other patient with manifest dystonia. |
| **Epilepsy** | Focal seizures with AO during first five years of life were reported in one patient(16). |
| **Dystonia** | Dystonia manifested in 9 of 13 patients, AO varied from 11 till 24 years and the disease progression from focal into severe generalised dystonia, 5 patients received deep brain stimulation with positive effect, 3 patients died. Dystonia did not respond to conventional drug treatment such as LDopa, biperiden and clonazepam(16). Three patients without dystonia were between 1y and 12y at the last follow-up. |
| **Hearing loss** | Profound hearing loss was documented in all patients, 7 patients received cochlear implant. |
| **Skeletal anomalies** | Skeletal anomalies (scoliosis) were documented in 3 patients(13, 16) and most probably represented secondary complications of progressive uncontrolled dystonia. |
| **Heart Defects** | Structural heart defects were not documented. |
| **Respiratory anomalies** | Respiratory features were present in 3 patients and included aspiration pneumonia, asthma, and impeded breathing during cold most probably |

| | |
|---|---|
| | representing secondary complication of progressive uncontrolled dystonia. |
| **GI anomalies** | Gastro-intestinal concerns were documented in 5 patients and included constipations reported in three individuals as well as achalasia reported in monozygotic twins(13). |
| **Genito-urinary anomalies** | No GU anomalies were documented. |
| **Skin and integument** | One patient presented with dermatitis. |
| **Repeated infections** | Repeated and/or excessive infections were not documented |
| **Thrombocytopenia and other haematological anomalies** | Thrombocytopenia was not observed. |
| **Healthy carrier** | All carries presented with congenital deafness; as dystonia is an age-related manifestation, its penetrance in the youngest patients remains unknown. |

Diagnostic and follow-up recommendations

- *ACTB*-DDs is diagnosed in patients with early onset severe hearing loss carrying MV *ACTB*:p.Arg183Trp.
- Hearing loss is severe and rapidly progressive suggesting that early cochlear implants should be considered to maintain adequate language development.
- The incidence of dystonia remains unknown but current data suggests that it may be as high as 100%.
- Regular monitoring of motor and language development with appropriate early intervention program if necessary.
- Early connection to the neurologist specialized in movement disorders is highly recommended to facilitate future treatment.
- Bilateral globus pallidus interna deep brain stimulation currently represents the only treatment option resulting in substantial clinical improvement.
- Supportive therapy such as early initiation of physiotherapy and application of the adaptive aids after onset of dystonia.

#### SUPPLEMENTAL NOTE S4: ACTG1-ASSOCIATED ISOLATED HEARING LOSS (ACTG1-ADHL)

Non-syndromic hearing loss was the first disorder associated with the cytoplasmic actin genes(17-19). Heterozygous variants in *ACTG1* were reported segregating in six families with multiple affected individuals including a large Norwegian family with more than 40 affected family members presenting with post lingual progressive sensorineural hearing loss, clinically defined as DFNA20/26(19, 20). The typical characteristics include a bilateral slowly progressive sensorineural hearing loss with the age of onset between the first and the third decades of life. The hearing loss begins at the highest frequencies and steadily progresses into profound deafness across all frequencies. The majority of the affected individuals would demonstrate the sloping configuration audiogram at the early age while hearing threshold remains intact at the lower frequencies. The hearing loss is progressing into deafness by the 6$^{th}$ decade(21). Tinnitus, vertigo, and other vestibular symptoms were occasionally reported in individuals with the *ACTG1*-associated hearing loss. The recent review summarized 36 *ACTG1* variants reported in patients with hearing loss. However, several patients presented with additional symptoms including other malformations and neurodevelopmental disorder(21) indicating that these patients should be classified as unspecified non-muscle actinopathies. Considering the highly variable symptomatic even within the same family(22), we recommend careful consideration of the clinical assignment in patients with the novel *ACTG1* variants especially when the molecular diagnosis was done early in life. Several missense variants such as T89I(17), K118M/N(17, 23, 24), K213R(25), E241K(23, 24), T278I(18), and V370A(19) were recurrently observed in large well characterized families with non-syndromic hearing loss and some of these variants were also studied *in-vitro* and *in vivo*(23, 26, 27). These variants can be reliably associated with the non-syndromic hearing loss.

The penetrance was reported as complete; however, the age of onset, progression and severity differ greatly even within the same family(19, 21).

Tabular summary of clinical features in patients with non-syndromic hearing loss (N=60)

| | |
|---|---|
| **Intellectual development and behaviour** | Normal development. |
| **Craniofacial anomalies** | No specific gestalt has been documented. |
| **Hearing loss** | Bilateral progressive sensorineural hearing loss with the typical begin at the higher frequencies and characteristic audiogram with the sloping configuration that may maintain even at the advanced stage. The progression rate varies from 1 dB/year to 6 dB/year(24). |
| **Vestibular symptoms** | Vestibular dysfunction, manifested as some equilibristic instability, was claimed occasionally by some of the elderly, profoundly hearing-impaired individuals but was formally assessed(19). Tinnitus is occasionally reported. |
| **Healthy carrier** | Not reported; hearing loss is an age-related phenotype with the variable onset within the same family. |

#### SUPPLEMENTAL NOTE S5: UNSPECIFIED NON-MUSCLE ACTINOPATHIES INCLUDING *ACTG1*-ASSOCIATED ISOLATED COLOBOMA

This cohort encompassed patients with missense variants in either *ACTB* or *ACTG1* whose clinical features did not fit any of the disorders described above. It is possible that future work might define novel distinct entities within this group, one of which could be an *ACTG1*-associated isolated coloboma(28). unNMA is diagnosed in a patient without BWCCF typical facial gestalt and/or brain malformation with a (likely) pathogenic missense variant in *ACTB* or *ACTG1* (except *ACTB* R183W) presenting with any phenotype other than post-lingual non-syndromic hearing loss.

In line with the previous section, we want to point out the high phenotypic heterogeneity in this group observed even within the same family. Although most of the patients presented with the neurodevelopmental disorder, the severity of the intellectual impairment is usually mild with good developmental progress under intensive speech and occupational therapy. Speech was usually more severely impaired in comparison with motor skills.  Speech delay was more prominent in children with the congenital or early onset hearing loss and remained a significant health issue even after the administration of the adequate hearing aids or cochlear implants.

The available data on adult individuals in unNMA (10 individuals older than 20y) indicates the stable course with no additional neurological or other health issues being developed. However, this statement would need to be confirmed in a larger patient cohort.

Tabular summary of clinical features in patients with unspecified NMA (N=66)

| | ***ACTB*** **N=36** | ***ACTG1*** **N=30** |
|---|---|---|
| **Intellectual development and behaviour** | 21 individuals, borderline/mild in 13 and moderate in 6, 7 patients had normal mental development; 8 patients presented prenatally or during neonatal period; 10 individuals with and without ID had behaviour anomalies, presented with ADHS, hyperactive and aggressive behaviour and temper tantrums; single patients were reported to have sleep disorder, Tourette syndrome and auto aggression. | 20 out of 30, mild in 7, moderate in 6 but also severe and profound in 3 individuals; 8 individuals had behaviour anomalies with ADHS and temper tantrums, as well as autism with obsessions described in 1 patient. |
| **Craniofacial anomalies** | Craniofacial anomalies were present in 27 individuals and were mild in the majority of the patients. Microcephaly was documented in 11 patients. Interestingly, microcephaly was a consistent feature in three patients with MV within the codon 152. | Mild craniofacial anomalies were described in 16 individuals presented with an unspecific pattern.<br>However, 4 individuals had ptosis accompanied by epicanthus in 2. Only two patients presented with microcephaly. |
| **Eye coloboma** | Iris coloboma was reported in two patients. | Iris coloboma was reported in three patients. |
| **MRI anomalies** | MRI anomalies were documented in 15 patients but nobody presented with cortical malformations except one patient with single PVNH. Structural abnormalities included abnormal corpus callosum in 3, enlarged ventricles in 3 and hydrocephalus in 1, posterior fossa anomalies in 2, as well as Chiari I anomaly, multiple | MRI anomalies were present in 7 patients, thereof 4 patients had cortical malformations including PMG in 2, dysgyria in 1 and PVNH in 1; the remaining 3 patients had either agenesis or hypoplastic corpus callosum. |

| | | |
|---|---|---|
| | calcifications and abnormal white matter signal in one patient, respectively. | |
| **Epilepsy** | Epilepsy was present in 5 patients, two of them had abnormal MRI such as Chiari I anomaly and multiple calcifications. Another patient was diagnosed with Doose syndrome. | Epilepsy manifested in 5 patients, two of them had cortical malformations (PVNH and PMG). |
| **Dystonia** | Dystonia was not documented. | Dystonia was not documented. |
| **Hearing loss** | Hearing loss was documented in 6 patients as bilateral sensorineural in three patients, mixed in one patient and conductive in another two patients. | Hearing loss was present in 21 patients, all patients had bilateral sensorineural hearing loss with AO from birth/first year till 3rd and 4th decades. However, adult onset was observed only in one multigenerational family with MV p.Ille85Leu. Other patients had the onset in early childhood. |
| **Skeletal anomalies** | Skeletal anomalies were documented in 11 and included vertebral anomalies (N=4) as well as pectus deformity, joint hypermobility, feet deformities, brachydactyly and long and slender fingers described in individual patients. Eight patients had short stature (till -3,4 z). | Skeletal anomalies were present in 9 patients, 5 had scoliosis, two had short stature (-4,7 z) and two bilateral feet deformities, respectively. |
| **Heart Defects** | Structural heart defects were present in 9 patients and included ASD, VSD, aortic coarctation, and PFO. Two patients had transposition of the great arteries and one had dextrocardia. Two patients had mitral valve prolapse. | Heart anomalies were seen in 5 patients as ASD/VSD, PDA, pulmonary stenosis and right descending aortic arch with aberrant left subclavian artery and diverticle of Kommerell, respectively. |
| **Respiratory anomalies** | Respiratory features were present in 4 patients and included severe and prolonged respiratory infections and pneumonias in three and respiratory distress in the remaining patient. | One patient had asthma; another patient presented with laryngomalacia and two patients had documented tracheomalacia in early months. |
| **GI anomalies** | Gastro-intestinal concerns were documented in 12 patients and required operative treatment in 4 patients.<br>Incomplete data in 6 patients and not assessed in 4 fetuses. | Gastro-intestinal concerns were documented in 4 patients and presented as duodenal atresia in 1, intestinal pseudo-obstruction and TNT dependency in 1, and constipations in the other two patients. In 8 patients GI data was incomplete. |
| **Genito-urinary anomalies** | GU anomalies included renal anomalies in 5 (pyelectasis/hydronephrosis, cystic dysplasia, and pyelonephritis) and abnormal genitalia in other 7 patients. | One patient presented with hydronephrosis, two with inguinal hernias and one with cryptorchidism. |

| | | |
|---|---|---|
| **Skin and integument** | Diverse dermatological concerns were recorded in 5 individuals: skin laxity, mild angiomas, photosensitivity and cutaneous infections with impetigo. | CALFs were documented in a single patient. |
| **Repeated infections** | Repeated and/or excessive infections were documented in 7 patients presented as recurrent respiratory infections including pneumonias in and multiple acute otitis media as well as chronic ear infections. However, 3 patients demonstrated systemic disorder with recurrent abscesses and cutaneous infections (158-B, 62-B and 119-B). One of these patients had thymus atrophy. One patient had periodic fever. | Repeated and/or excessive infections were documented in three patients. |
| **Thrombocytopenia and other haematological anomalies** | Three patients had thrombocytopenia presented as borderline or mildly diminished platelet count without manifesting bleeding disorder. All 3 patients had MV in the last exon. | Thrombocytopenia was not documented. |
| **Healthy carrier** | All carries demonstrated either ID or structural/morphological anomalies. | All carries demonstrated either ID or structural/morphological anomalies. |

Prenatal manifestation in unNMA

Abnormal prenatal history was documented in 15 patients (N=9 with variants in *ACTB* and N=6 in *ACTG1*). Whereas increased nuchal translucency was the most common prenatal feature in patients with BWCFF, it was reported in only two pregnancies in the unNMA cohort. Other features included ventriculomegaly, heart defects, cleft lip/palate, duodenal atresia, omphalocele, and fetal arrhythmia. Prenatal molecular diagnosis was made in four cases and led to the termination between 16 and 28 GWs. Three of four foetuses presented with ventriculomegaly or hydrocephalus, one had IUGR, transposition of the great arteries, renal cysts and omphalocele. Detailed neuropathological examination of the cerebral was available in two cases and reported normal cortical structure.

Diagnostic and follow-up recommendations

- Considering the high clinical heterogeneity within the unNMA patient cohort and still limited information about the natural history, developing general recommendations regarding the clinical management remains difficult.
- Referral to an early intervention program is strongly recommended for the detailed developmental and behaviour evaluation and intervention.
- Medical surveillance should be focused on individual presentation of the patients and may include the control of the growth parameters, cardiac evaluation, hearing test, ophthalmological surveillance and other evaluations depending on individual concerns. Young patients with uncertain clinical classification should have annual follow-up and their families should be informed that clinical diagnosis is ambiguous and so remains developmental and neurological long-term prognosis; families should be offered the maximal BWCFFs-oriented management that can become less intensive or lifted completely if BWCFFs can be prospectively excluded.

#### SUPPLEMENTAL NOTE S6: TRANSCRIPTOME SEQUENCING

##### Missense variants in CYA genes do not have major impact on overall gene expression.

In line with the overlapping expression profiles, we observed only few differentially expressed genes between patients derived and control cell cultures. Only one gene (*OLFM1*) was differentially expressed at an FDR-adjusted p-value cutoff of 0.01 in comparison with BWCFF patient cell cultures to control 2 cell cultures, whereas all other disease-specific cell cultures did not show any differentially expressed genes in comparison to control 2 cell cultures (Supplementary Table 3 Differential Gene Expression Analyses). Of note, seven fibroblast cultures of control 2 group were established under identical conditions like most of the patient derived cultures whereas control 1 consisted of three cultures acquired from Coriell. Some more genes were differentially expressed in comparison to control 1 cell cultures, but only for BWCFF vs. control 1 (90 genes) and *ACTB*-BWCFF vs. control 1 (43 genes).

Analysing the expression of the genes encoding for actin isoforms and actin-binding proteins(ABP) (as in Latham et al.(2)) we observed one cluster that mainly contained BWCFF samples (Suppl. Figure S8) together with another larger cluster with three subclusters including three control 2 samples in a subcluster, control 1 samples form a subcluster control samples, Dystonia Deafness, non-BWCFF and BWCFF samples that were more wide-spread across the subclusters. Therefore, analysed actin variants have only minor systemic impact even on the expression of ABP-encoding genes.

#### SUPPLEMENTAL NOTE S7: BWCFF VARIANT T120I IN EITHER CYTOSKELETAL ACTIN ISOFORM RESULTS IN EARLY LETHALITY IN THE MOUSE.

Gene editing strategy was overall successful but did not result in generation of the viable constitutive *Actb* T120I or *Actg1* T120I mouse neither via direct microinjection in oocytes nor by generating targeted ES cells.

Knock-In Mouse Model for T120I in Mouse *Actb*

Supplementary Figure S19 demonstrates step by step gene editing strategy. After the guide efficiency was tested (Suppl. Figure 19B), CRISPR/Cas9 complex was injected in the oocytes in combination with various guides and homology templates (Table S6-1).

In total 482 injected zygotes were transferred into recipient foster mice that yielded only 34 pups with only 22 pups survived through weaning. All survived pups were wild types in both *Actb* alleles.

| **Injection** | **Guides** | **Oligo** | **Oocytes injected** | **Oocytes transferred** | **Forsters** | **Offspring** |
|---|---|---|---|---|---|---|
| I014.1 | I014.88 | I014.5 | 133 | 102 | 3 | 0 |
| I014.2 | I014.88 | I014.5 | 189 | 144 | 5 | 7 |
| I014.3 | I014.88 | I014.5 | 172 | 124 | 4 | 16 |
| I014.4 | I014.88+81 | I014.8 | 154 | 68 | 2 | 9 |
| I014.5 | I014.2 | I014.8 | 137 | 48 | 2 | 2 |
| total | | | 614 | 482 | 16 | 34 |
| Table S6-1 | | | | | | |

In the next step, homologous recombination using the oligo template was verified via targeting in ES cells. Suppl. Figures 19C-D demonstrate the genotyping strategy of the overall highly efficient CRISPR targeting with 42% in I014.1 and up to 90% in I014.2. The sequencing results from selected and expanded clones with the expected heterozygous mutant are shown in Suppl. Figure 19E (clones 1G4, 4B1, and 4H9). Three selected clones were injected on eight different occasions (Table S6-2) resulting in visible pregnancies in multiple transferred fosters, however, only 10 pups were born, of those only one female showed a very low grade chimerism on the fur.

| **Injection** | **Clones** | **Blasts injected** | **Blasts transferred** | **Forsters** | **Pups** |
|---|---|---|---|---|---|
| I014.6 | 1G4, 4B1 | 43 | 38 | 2 | 0 |
| I014.7 | 1G4 | 34 | 31 | 2 | 0 |
| I014.8 | 1G4, 4B1, 4H9 | 75 | 44 | 3 | 0 |
| I014.9 | 1G4, 4B1 | 80 | 66 | 3 | 0 |
| I014.10 | 4B1, 4H9 | 78 | 67 | 4 | 0 |
| I014.11 | 1G4, 4B1 | 73 | 58 | 3 | 10 |
| I014.12 | 1G4, 4B1 | 46 | 40 | 2 | 0 |
| I014.13 | 1G4, 4B1, 4H9 | 54 | 52 | 3 | 0 |
| total | | 483 | 396 | 22 | 10 |
| Table S6-2 | | | | | |

The fact that the injection of the several independent clones of the heterozygous ES cells failed to generate even a low-grade chimera with only one unconvincing exception let us assume that the phenotype of T120I in *Actb* is too severe to lead to a viable organism in the mouse.

Knock-In Mouse Model for T120I in Mouse *Actg1*

Supplementary Figure S21 demonstrates step by step gene editing strategy. After the guide efficiency was tested (Suppl. Figure 20B), CRISPR/Cas9 complex was injected in the oocytes in combination with various guides and homology templates (Table S6-3).

In total 342 injected zygotes were transferred into recipient foster mice that yielded only 10 pups surviving through weaning. All but one surviving pups were wildtype in both *Actg1* alleles. One offspring had a double-allele alteration.

| **Injection** | **Guides** | **Oligo** | **Oocytes injected** | **Oocytes transferred** | **Forsters** | **Offspring** |
|---|---|---|---|---|---|---|
| I018.1 | I018.G1+G2 | I018.1 | 168 | 124 | 4 | 1 |
| I018.2 | I018.G1+G2 | I018.1 | 199 | 161 | 6 | 7 |
| I013.3 | I018.G1+G2 | I018.1 | 123 | 57 | 2 | 2 |
| total | | | 490 | 342 | 12 | 10 |
| Table S6-3 | | | | | | |

In the next step, homologous recombination using the oligo template was verified via targeting in ES cells. Suppl. Figures 20C-D demonstrate the genotyping strategy of the overall highly efficient CRISPR targeting with 35% for guide 1 and 8% for guide 2. The sequencing results from selected and expanded clones with the expected heterozygous mutant are shown in Suppl. Figure 19E (clones 1A4, 1D10, and 1E5). Three selected clones were injected on three different occasions (Table S6-4) resulting in visible pregnancies in multiple transferred fosters, and 11 pups showing a certain grade of chimerism from clone 1A4 (30% female and 30% male) as well clone 1D10 (3 30-100% chimeras).

| **Injection** | **Clones** | **Blasts injected** | **Blasts transferred** | **Forsters** | **Pups** |
|---|---|---|---|---|---|
| I018.4 | 1A4, 1C11 | 45 | 39 | 2 | 6 (1A4) |
| I018.5 | 1D10 | 25 | 22 | 1 | 5 |
| I018.6 | 1D10, 1E5 | 62 | 56 | 3 | 0 |
| total | | 133 | 117 | 6 | 11 |
| Table S6-4 | | | | | |

Founder animals from clone 1A4 all died between the age 8 and 15 weeks with visible signs of malocclusion. I018.12 (Suppl. Figure 20E) was attempted to breed but did not father offspring. Chimeric mice from clone 1D10 were mated to C57Bl/6Ng mice to assess transmission to the germ line. Fur-matched offspring could be obtained and three out of five offspring mice were positive by showing a HindII-digestable restriction pattern and did not demonstrate any obvious morphological anomalies. The sequencing, however demonstrated and additional frameshift mutation in cis with the edited missense variant, meaning that the mutated *Actg1* gene is truncated and non-functional (Suppl. Figure S21). This was in line with the normal phenotype of all F1 animals. Considering the lethality of the 30% chimera with the obvious morphological anomalies we concluded that phenotype of T120I not only in *Actb* but also in *Actg1* is too severe to lead to a viable organism in the mouse.

**Supplementary Figure S1. Differences in population genetic variability of actin loci.**
**(A)** Stacked column charts demonstrating functional consequences of all possible singe nucleotide substitutions simulated within the genomic regions of *ACTB* (NM_001101.5; hg38 7:5527148-5530601) and *ACTG1* (NM_001614.5; hg38 17:81509971-81512799) annotated with the Ensembl Variant Effect predictor (VEP) compared to the small nucleotide variants observed in public databases with (with recurrences) and without (w/o recurrence) population frequencies of the variants; variant types are listed in alphabetical order **(B)** integrated ucsc genome browser view demonstrating two paralogous regions within coding regions of *ACTG1* and *ACTB* (sequence differences are highlighted in red), I and I' relative usage of reference (dark blue) and alternative codon (light blue), II and II' PhyloP basewise conservation score (100 vertebrates), III and III' number of recurrent variants in public databases, pink top on bars indicates the data exceeding the display limit of 5,000.

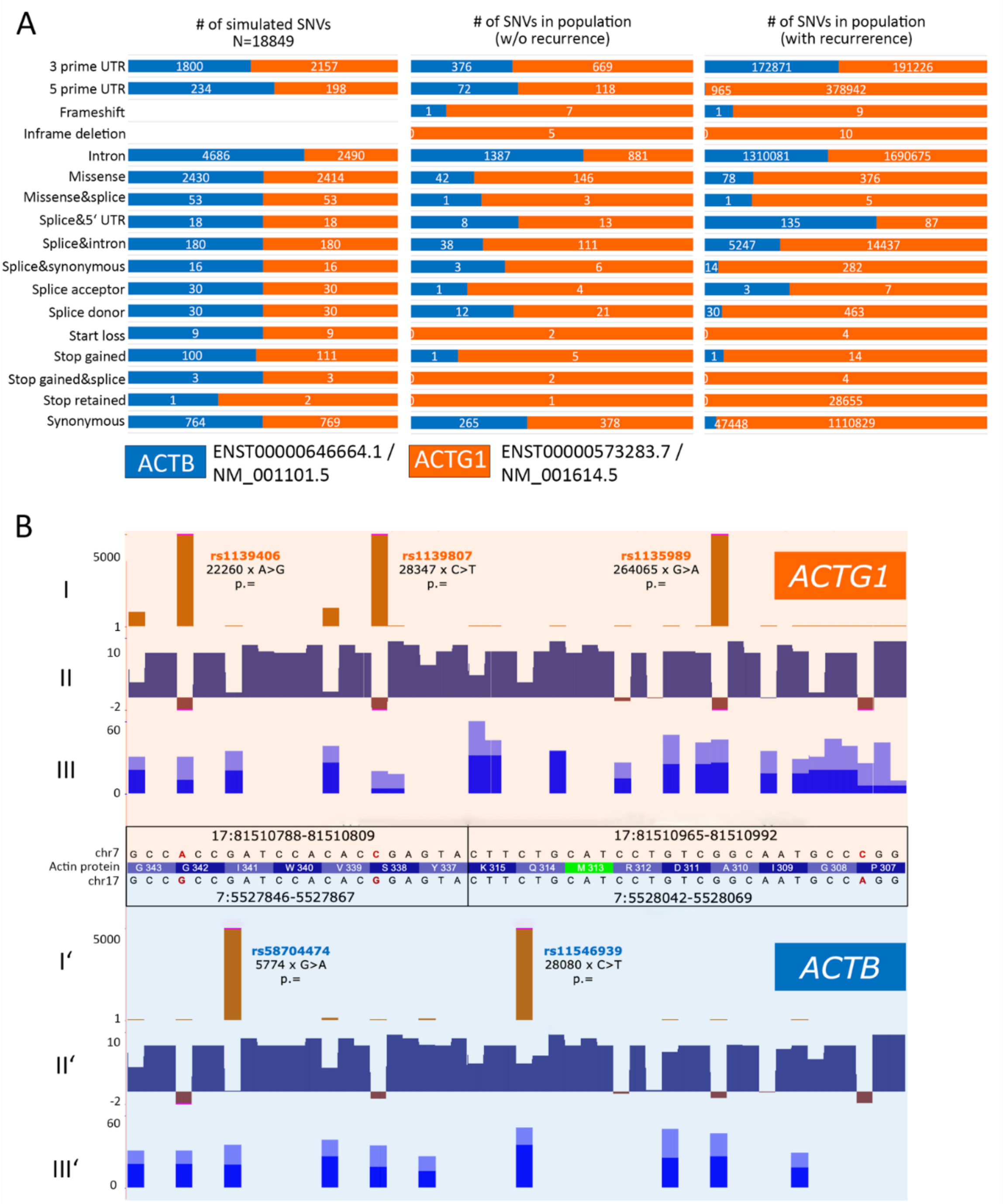

**Supplementary Figure S2. Compatible number of cancer-associated somatic variants in CYA genes**

**(A)** Stacked column charts demonstrating the spectrum and frequencies of somatic small nucleotide variations in *ACTB* and *ACTG1* observed in COSMIC and **(B)** cBIOPortal databases; note that cBIOPortal supports only non-synonymous and coding region small nucleotide variants; colour code and annotations correspond to the Suppl. Figure S1.

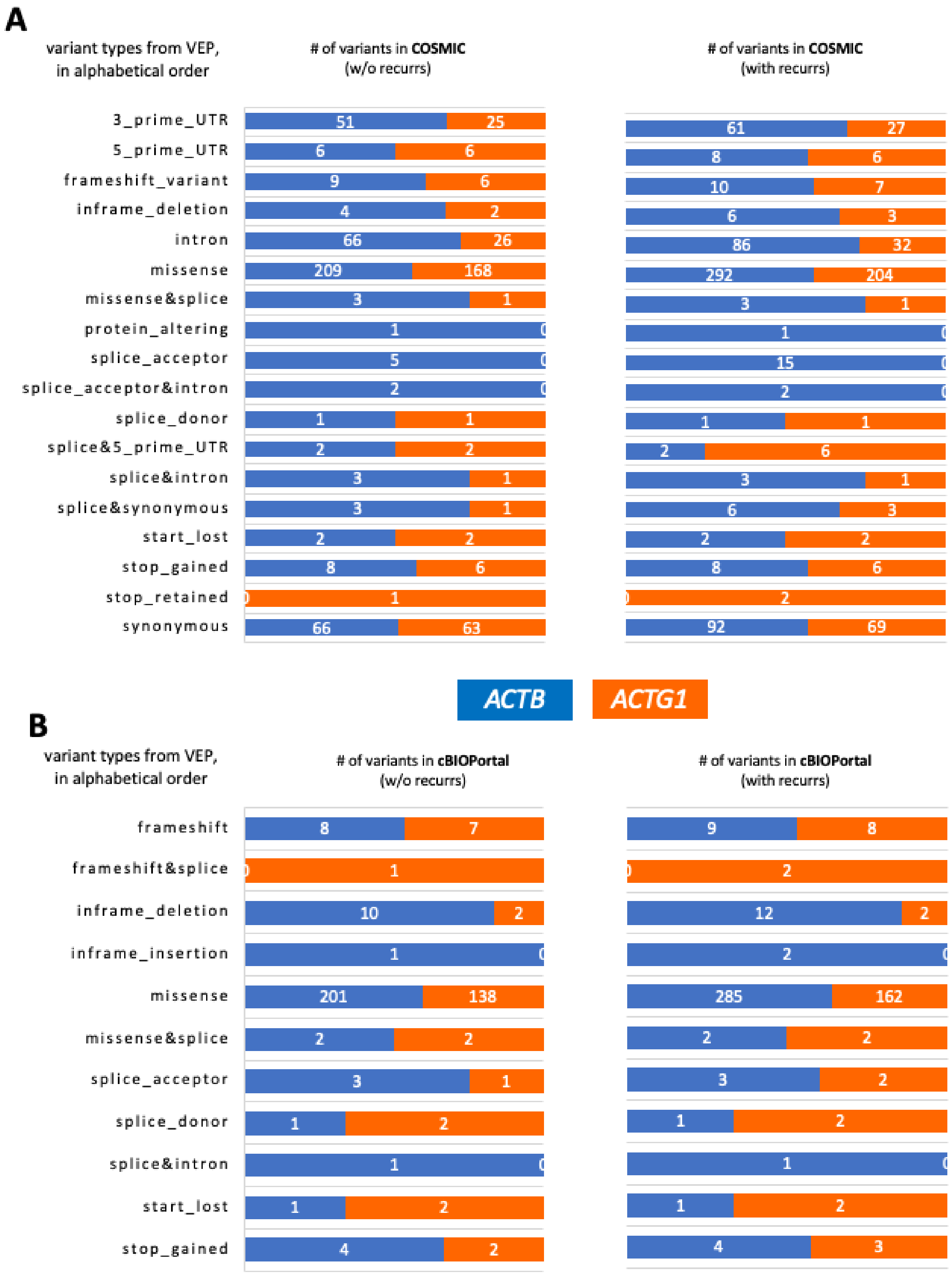

**Supplementary Figure S3. Classification of the NMA patient cohort applying genomic and phenotypic-led approach**.

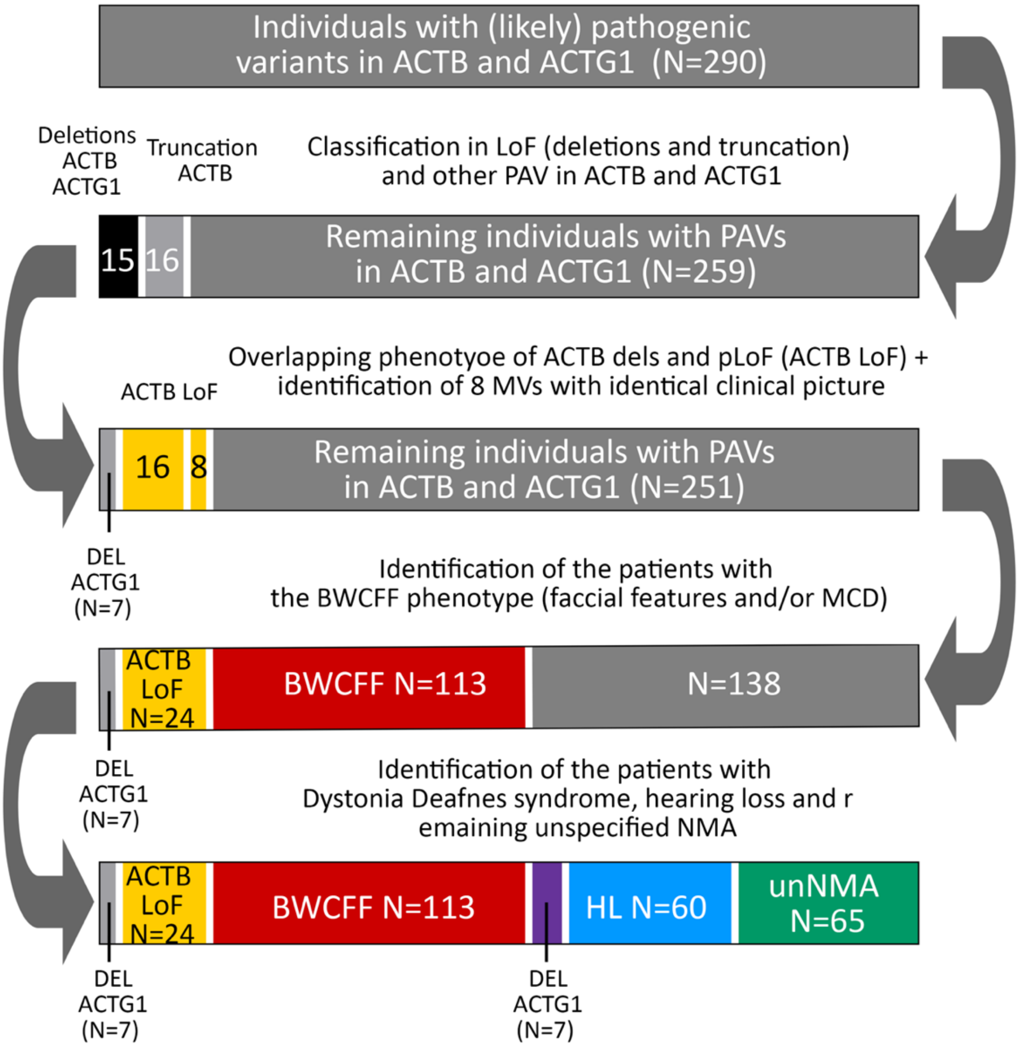

**Supplementary Figure S4. GestaltMatcher analysis of the NMA spectrum.**

tSNE visualization of different groups in GestaltMatcher analysis and the mean pairwise distance distribution of cohorts sampled from (blue) same syndrome, (red) different syndrome, and the orange distribution of the target comparisons. The threshold (c) is 0.896. When more than 50% of the orange region fall above the threshold, it indicates the two disorders are not similar. **(A)** The mean pairwise distance between BWCFF and *ACTB*_LoF patients is 0.977, and 100% of the sampling is above the threshold (the region falling on the right of the threshold). **(B)** The tSNE visualization between *ACTB*_BWCFF and *ACTG1*_BWCFF. **(C)** The mean pairwise distance between BWCFF_*ACTB* and BWCFF_*ACTG1* patients is 0.827, and 0% of the sampling is above the threshold. **(D)** The mean pairwise distance between unNMA and BWCFF patients is 0.916, and 92% of the sampling is above the threshold. **(E)** The mean pairwise distance between unNMA and *ACTB*_LoF patients is 0.962, and 98% of the sampling is above the threshold. F) The tSNE visualization between *ACTB*_unNMA and *ACTG1*_unNMA. **(G)** The mean pairwise distance between *ACTB*_unNMA and *ACTG1*_unNMA patients is 0.924, and 92% of the sampling is above the threshold. **(H)** The mean pairwise distance between BWCFF and BWCFF_unNMA patients is 0.853, and 1% of the sampling is above the threshold.

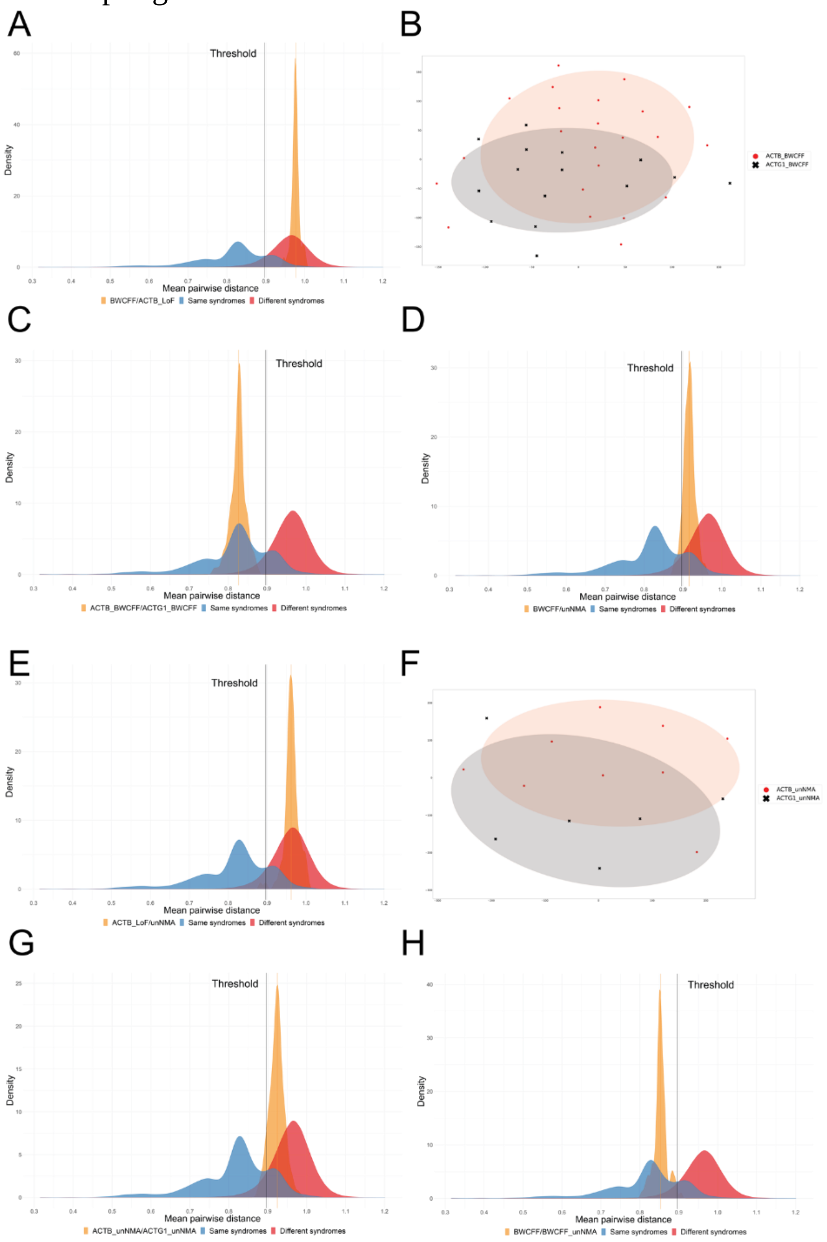

**Supplementary Figure S5. Western blots of βCYA in patient-derived and control fibroblasts**.

**(A)** Fluorescent total protein membrane staining (Revert$^{TM}$ 700 Total protein stain), gray scale image; samples are labeled corresponding to sample identifiers in Figure 4A, R183W corresponds to 25-B and R183W' to 176-B (**B)** Fluorescent bCYA protein detection using IRDye800CW. **(C)** Ponceau S total protein staining (dark background) **(D)** Chemiluminescent bCYA protein detection, samples are labeled corresponding to sample identifiers in Figure 4A. **E)** bCYA protein expression in patient-derived fibroblasts by western blot in the experimental set up using fluorescent detections. **(F)**□bCYA protein expression in patient-derived fibroblasts by western blot in the experimental set up using chemiluminescent detections.

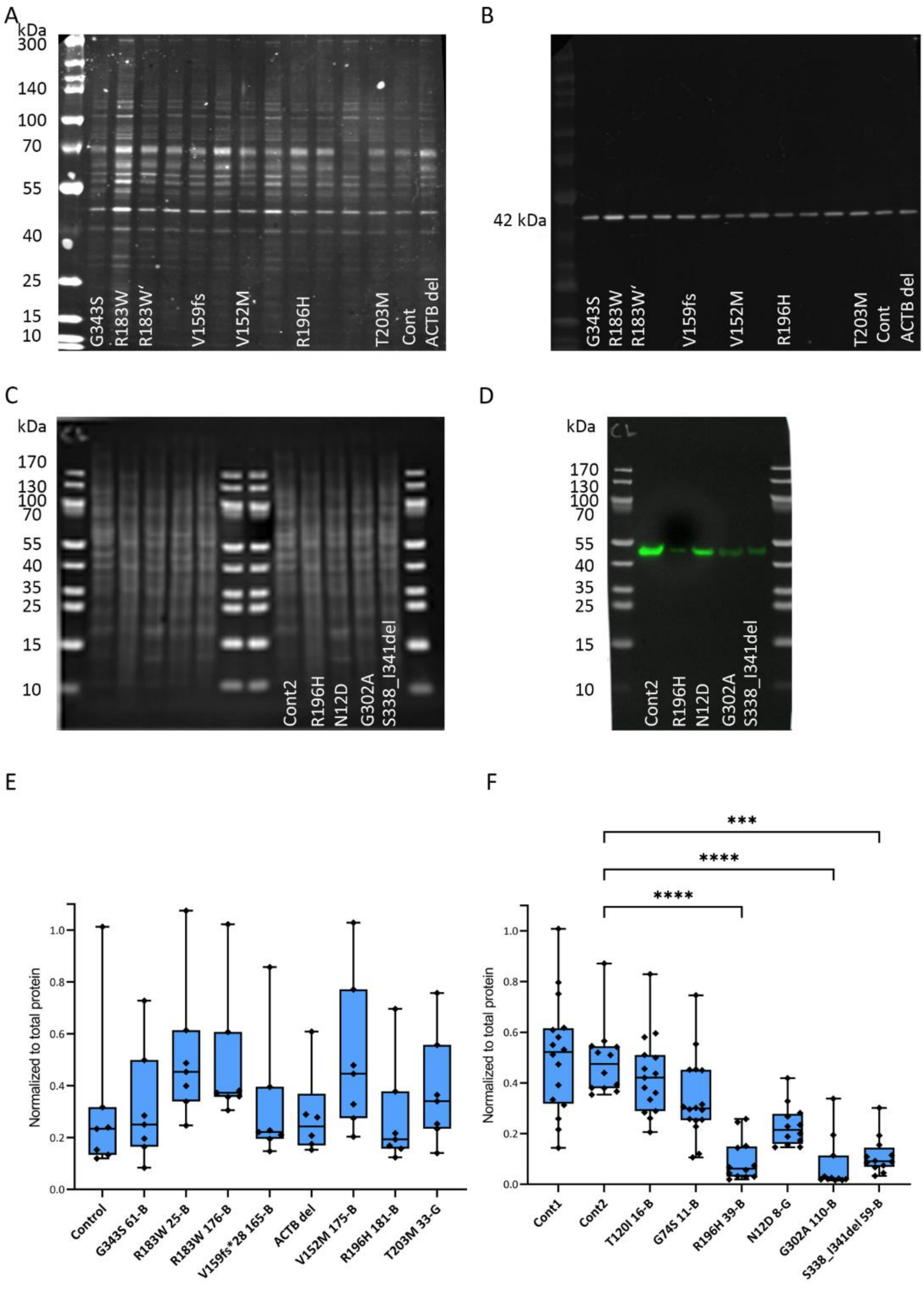

**Supplementary Figure S6. Western blots of gCYA in patient-derived and control fibroblasts**.

**(A)** Fluorescent total protein membrane staining (Revert$^{TM}$ 700 Total protein stain), gray scale image; samples are labeled corresponding to sample identifiers in Suppl. Figure 5 R183W corresponds to 25-B and R183W’ to 176-B (**B)** Fluorescent bCYA protein detection using IRDye800CW. **(C)** Ponceau S total protein staining (dark background) **(D)** Chemiluminescent bCYA protein detection, samples are labeled corresponding to sample identifiers in Suppl. Figure 6. **(E)** γCYA protein expression in patient-derived fibroblasts by western blot in the experimental set up using fluorescent detections. **(F)**□gCYA protein expression in patient-derived fibroblasts by western blot in the experimental set up using chemiluminescent detections.

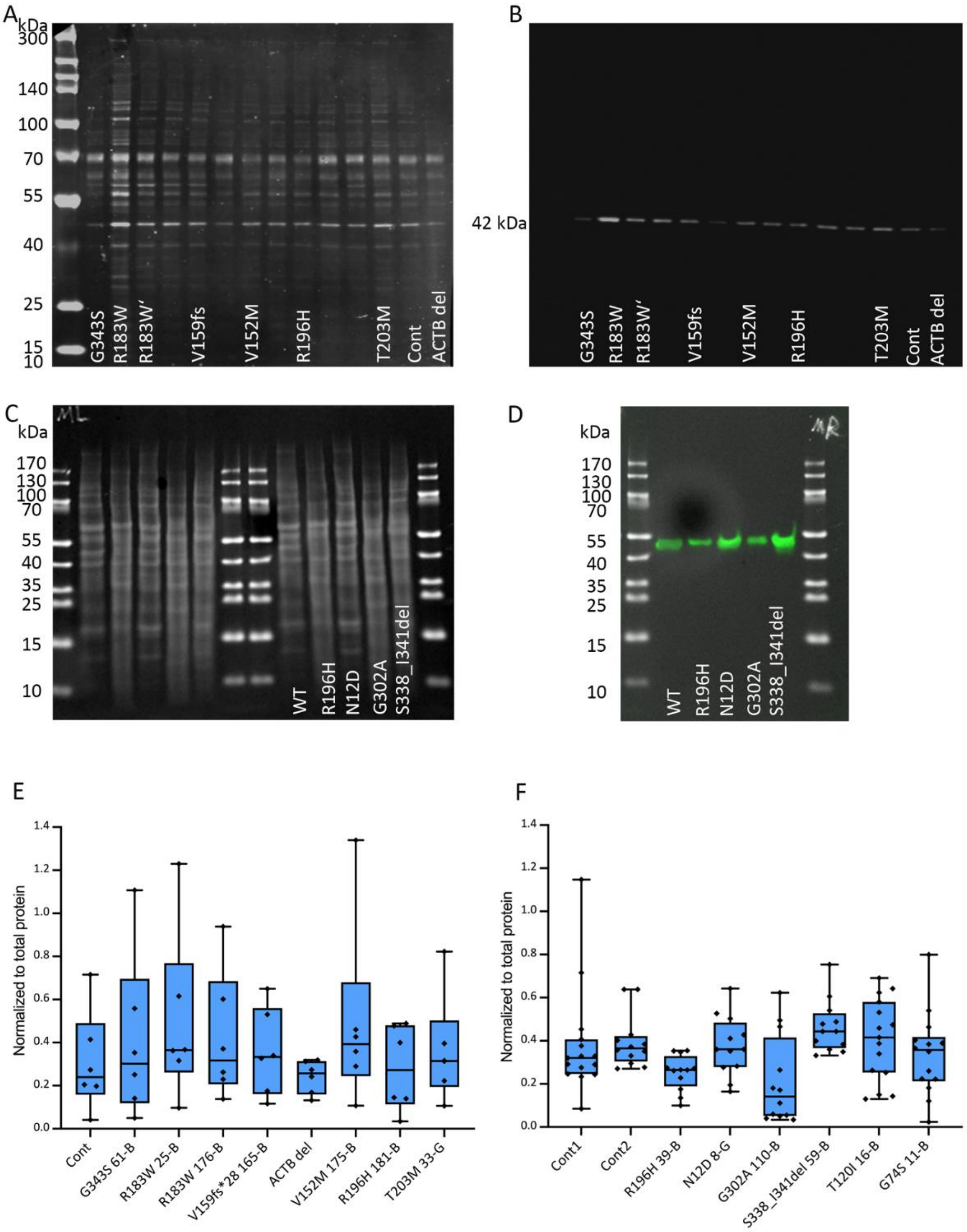

**Supplementary Figure S7. Western blots of panactin in patient-derived and control fibroblasts**.

**(A)** Fluorescent total protein membrane staining (Revert™ 700 Total protein stain), gray scale image; samples are labeled corresponding to sample identifiers in Figure 4B, R183W corresponds to 25-B and R183W' to 176-B (**B)** Fluorescent bCYA protein detection using IRDye800CW. **(C)** Ponceau S total protein staining (dark background). **(D)** Chemiluminescent panactin protein detection, samples are labeled corresponding to sample identifiers in Figure 4B **E)** Panactin expression in patient-derived fibroblasts by western blot in the experimental set up using fluorescent detections **(F)** Panactin protein expression in patient-derived fibroblasts by western blot in the experimental set up using chemiluminescent detections.

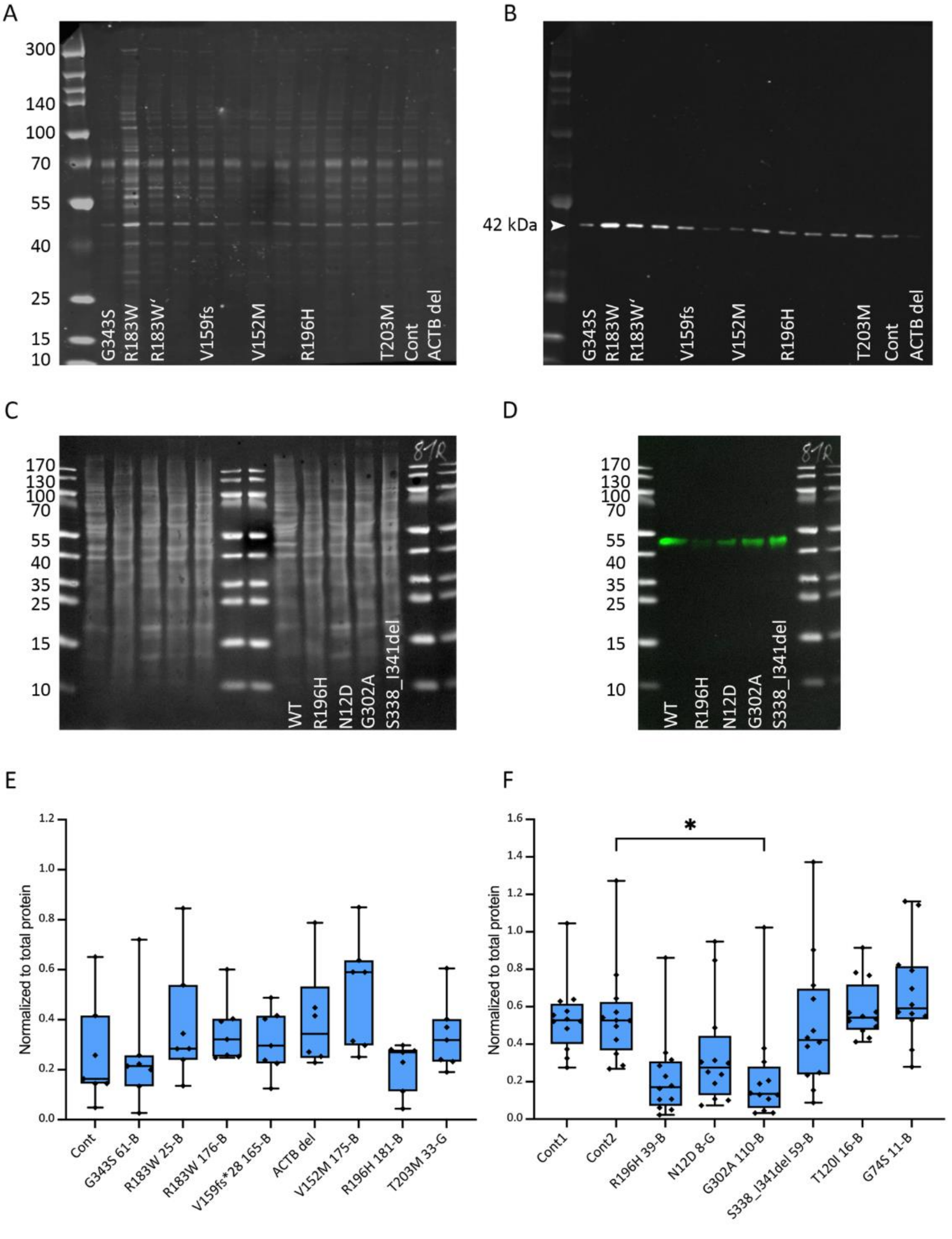

**Supplementary Figure S8. Expression profiles of the patient-derived and control fibroblasts.**

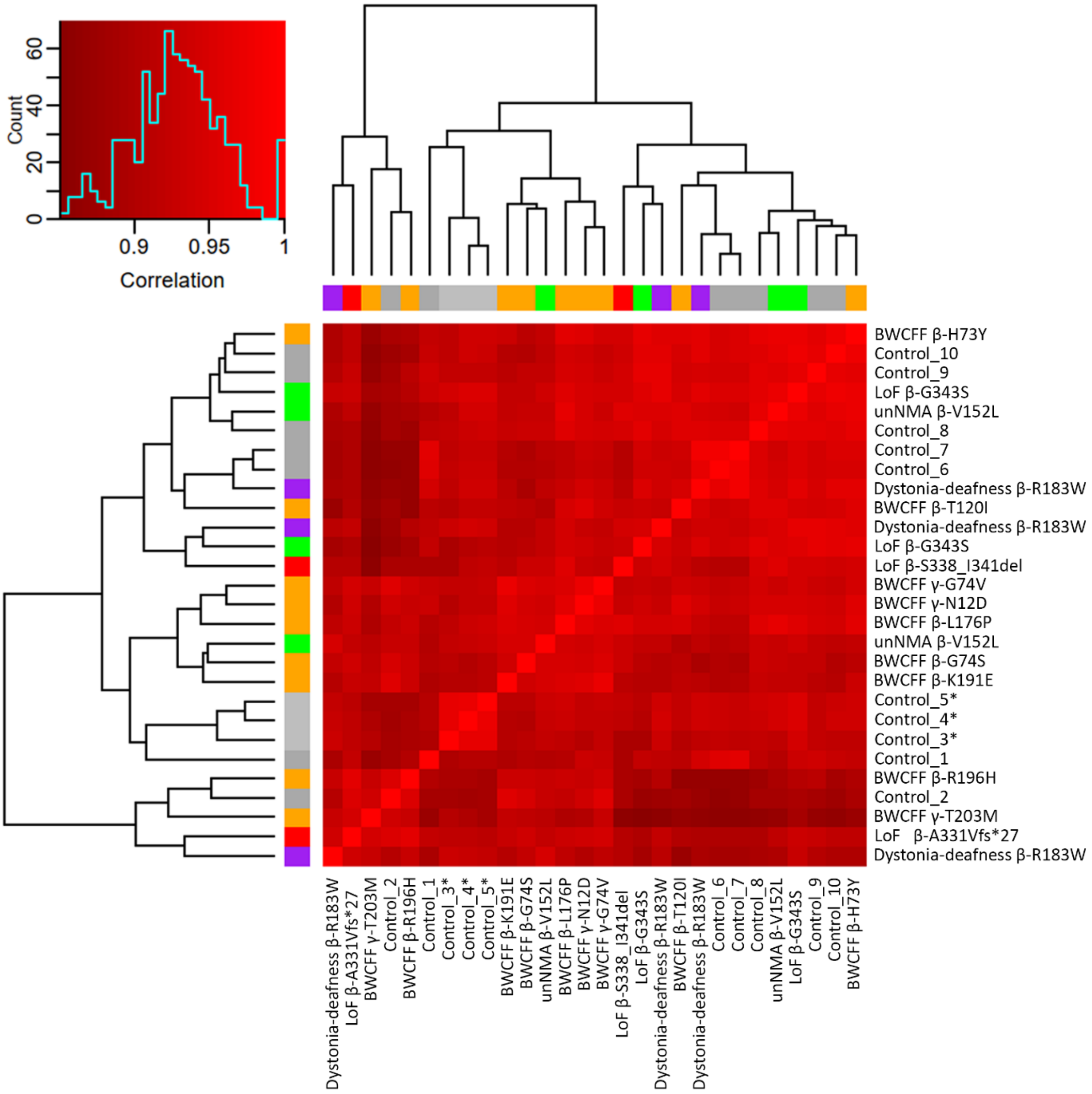

**Supplementary Figure S9. Principle component analysis of the average expression profile per patient.**

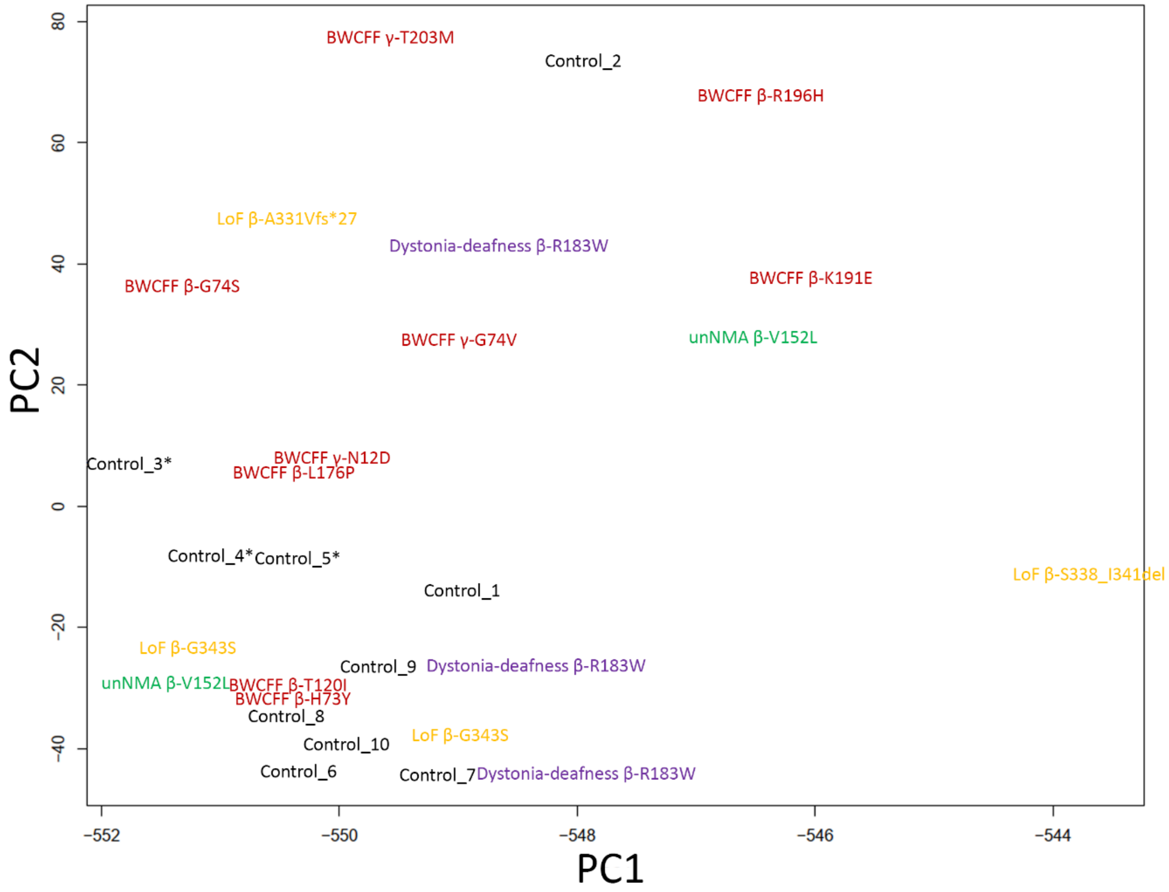

**Supplementary Figure S10. Dynamic cellular morphology depending on cell confluence in culture with *ACTB* variants associated with the *ACTB* functional haploinsufficiency**.

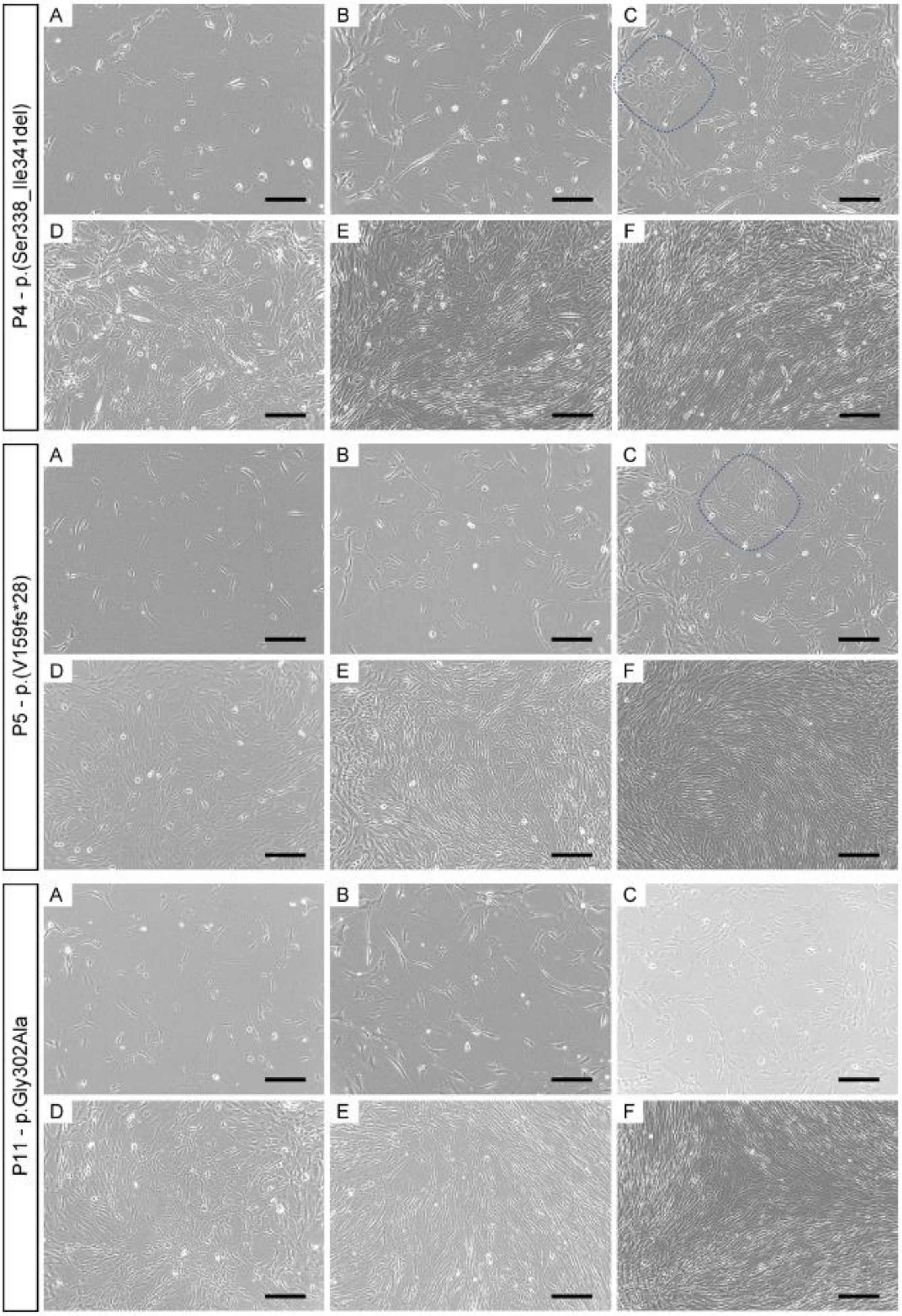

**Supplementary Figure S11. Morphology of control and patient-derived fibroblasts selected MV in *ACTB* and *ACTG1***

Phase contrast images were acquired at 10x magnification, scale bar = 200 µm; note relatively large cellular area of the fibroblast with bT120I (BWCFF) and gT203M (BWCFF) variants and long tubule-like processes visible in bG74S culture (arrows); two cultures from two unrelated patients with the variant bR196H appeared rather small; the morphology of bR183W and bG302A cultures were comparable to control.

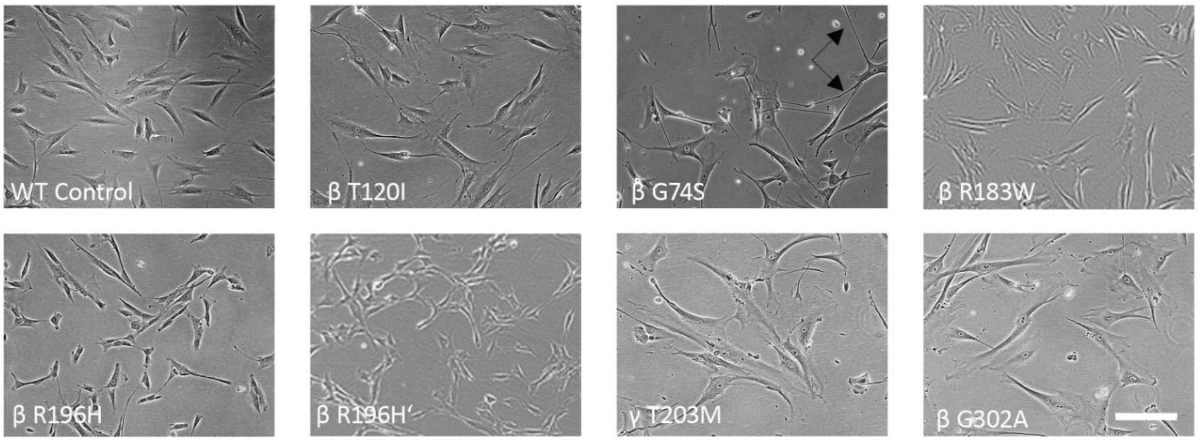

**Supplementary Figure S12. Pyrene–based bulk-polymerization and depolymerization experiments of CYA isoforms** (5% pyrene–labeled).

**(A, B)** Representative traces of pyrene-polymerization experiments with wild type b-actin and mutants. Experiments were performed with pure actin mutants (A) or a 1:1 mixture of wild type and mutant actin (B) **(C)** Representative traces of pyrene-based dilution-induced depolymerization experiments performed with pure wild type b-actin and mutant proteins. (**D, E**) Representative traces of seeded pyrene-polymerization experiments with g-actin wild type and mutants. Experiments were performed with pure actin mutants (D) or a 1:1 mixture of wild type and mutant actin (E). **(F)** Representative traces of pyrene-based dilution-induced depolymerization experiments performed with pure wild type g-actin and mutant proteins.

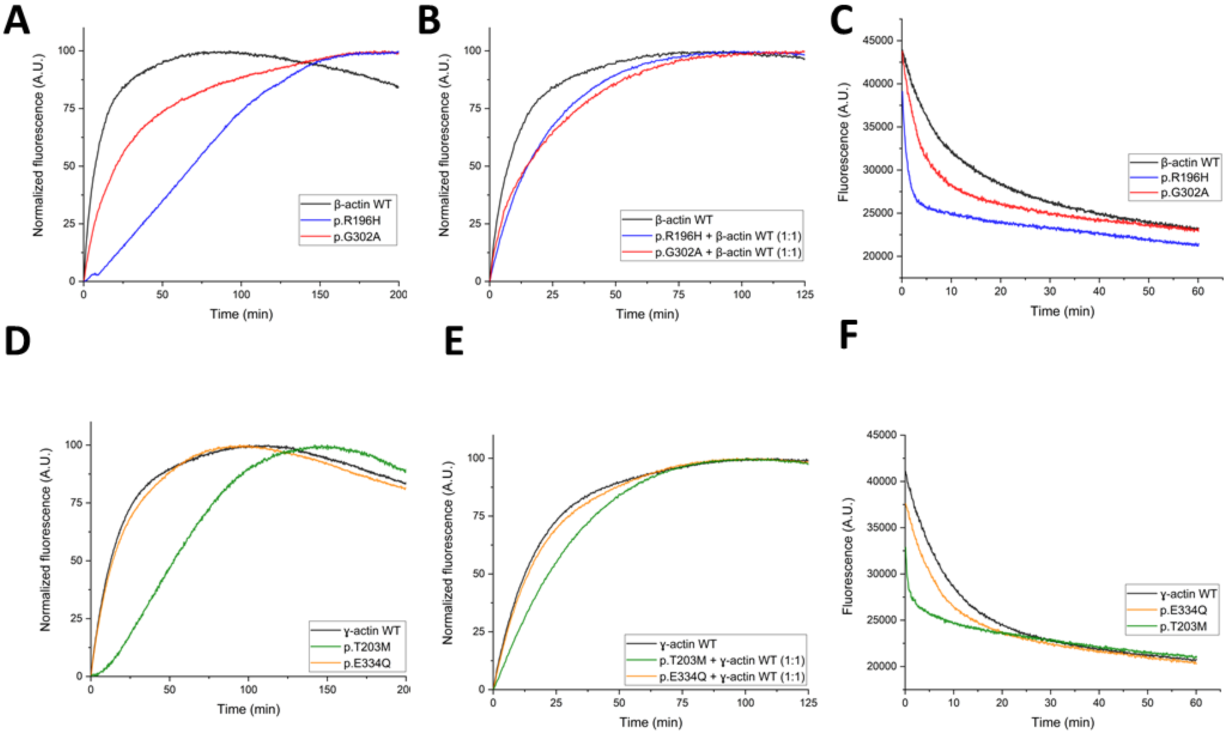

**Supplementary Figure S13.** ***Actb*** **siRNA knockout optimization in cultured N2a cells.** The concentration of the siRNA and the time of qPCR analysis after transfection is mentioned at the top of each panel. Each panel represents data from 3 technical repeats of scrambled and *Actb* siRNA transfections. The thick horizontal line for each experimental group represents the mean and the shorter thin horizontal lines represent the standard error of the mean. The statistical comparison was done via a t-test and the two tailed p-values are provided for each comparison.

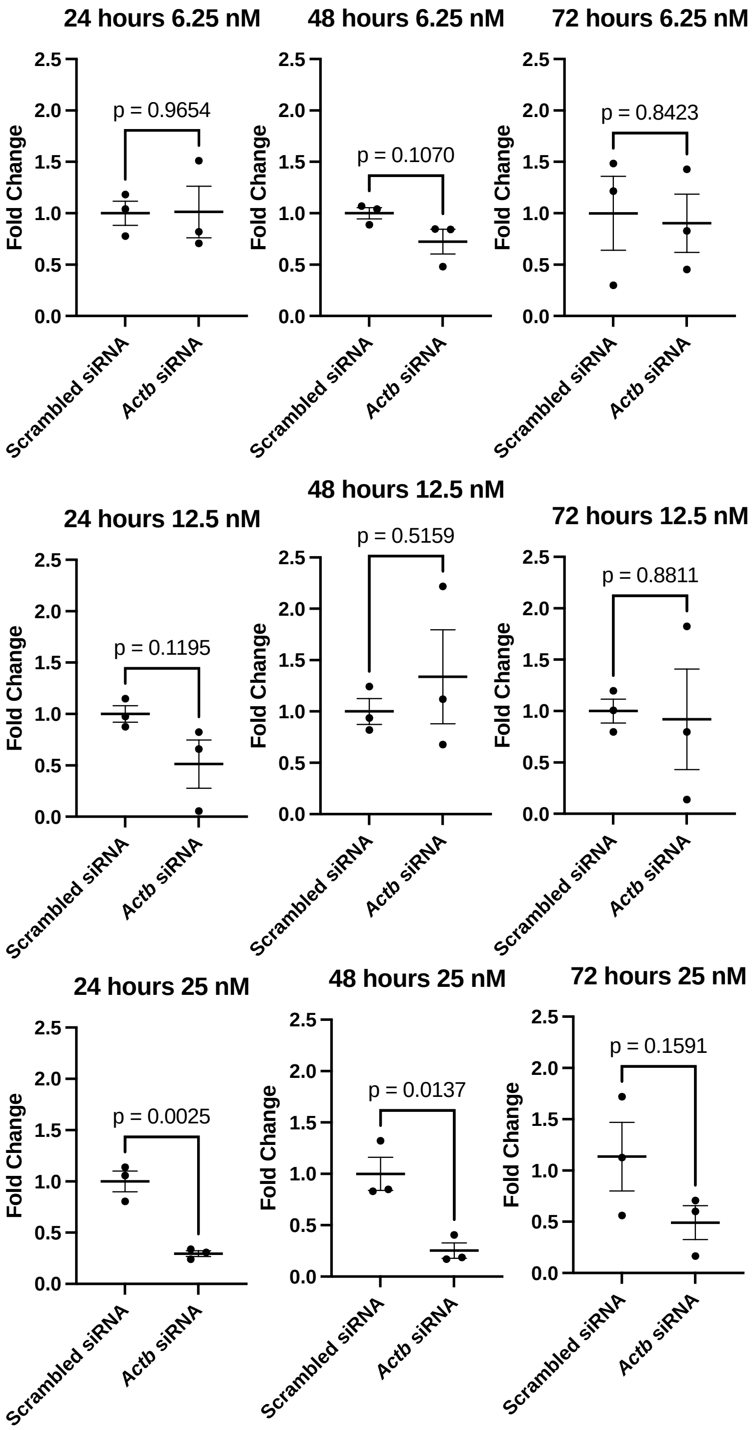

**Supplementary Figure S14. Quantitative western blots of total protein extracted from N2a cells post *Actb* siRNA transfection.**

20 µg of total protein extract from N2a cells was used to examine *Actb* at 24, 48 and 72 hours post transfection. The top panel is a representative image of western blots used to quantify the *Actb* expression in the N2a cells. The lanes are labelled in the figure and the right lane contains protein standard marker. Blot is targeted with *Actb*, Beta-tubulin and GapDH. The bottom panel shows the quantification of the images. The statistical comparison was done via an individual t-test and the two tailed p-values are provided for each comparison.

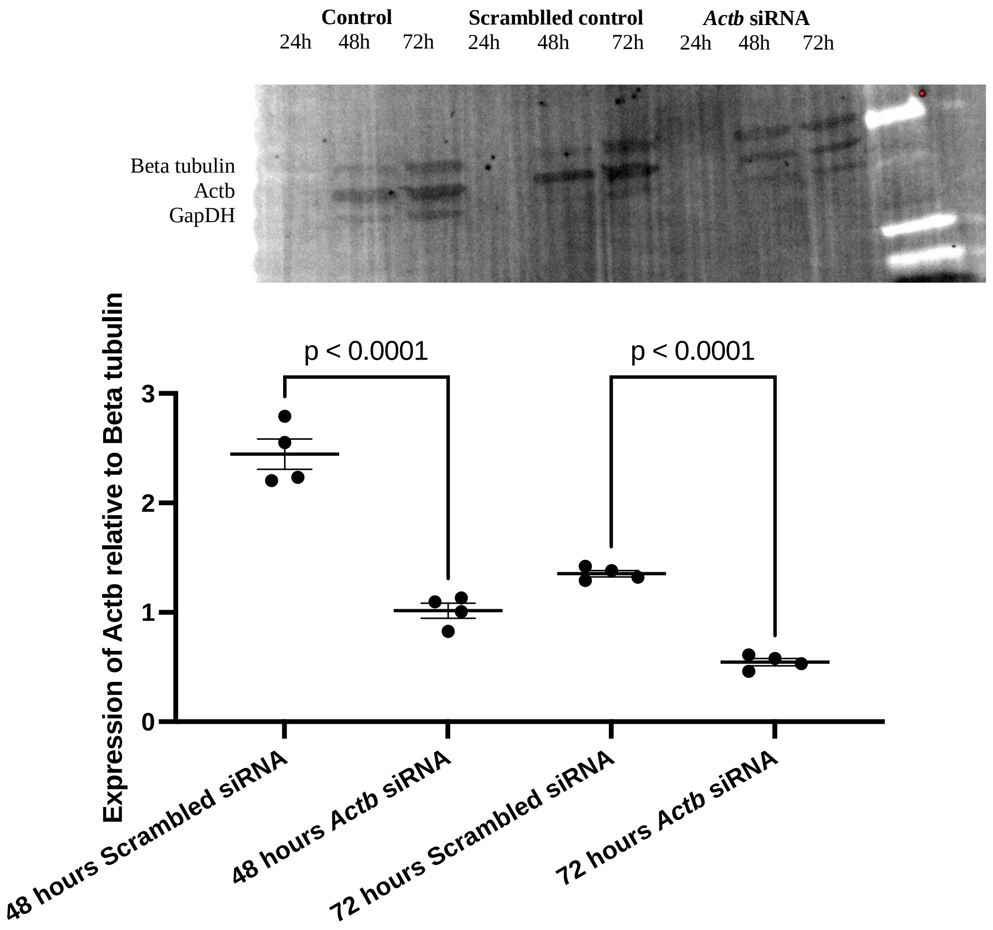

**Supplementary Figure S15. Characterisation of the siRNA-mediated *Actb* knockdown in mouse N2A neuronal cell line.**

**(A) BrdU proliferation assay** - N2a cells were grown on two glass inserts in each well of a 6 well plate and allowed to grow to approximately 70% confluency, exposure to BrdU was for one hour and the cells fixed for IHC after one hour exposure. Exposure to BrdU was performed in triplicate pairs. All groups have similar uptake of BrdU after IHC staining. The thick horizontal line for each measurement group represents mean and the shorter thin horizontal lines represent the SEM. The statistical comparison was done via a one-way ANOVA test with multiple comparisons and the p-values are provided for each comparison. BrdU integration to replicating cells is not impaired by *Actb* knockdown showing normal cellular proliferation. **(B) LDH cells stress assay** - Three technical replicates were used for each group. The thick horizontal line for each measurement group represents mean and the shorter thin horizontal lines represent the SEM. The statistical comparison was done via a one-way ANOVA test with multiple comparisons and the p-values are provided for each comparison. LDH levels measured showed no significant increase between all groups (untransfected, scrambled siRNA and *Actb* siRNA). Total LDH measured does increase over time in culture for all three experimental groups. The total LDH possible provided by lysis cells shows significant differences to the other groups showing significantly more LDH was present in these wells when tested.

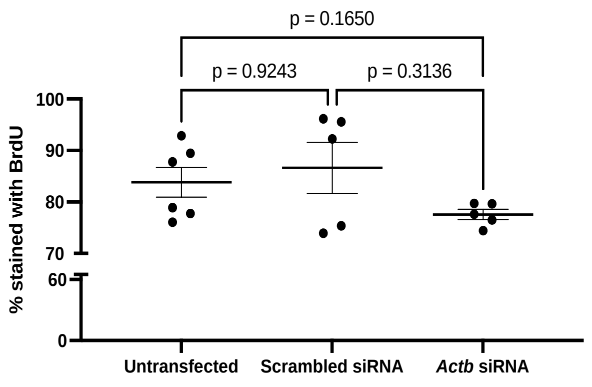

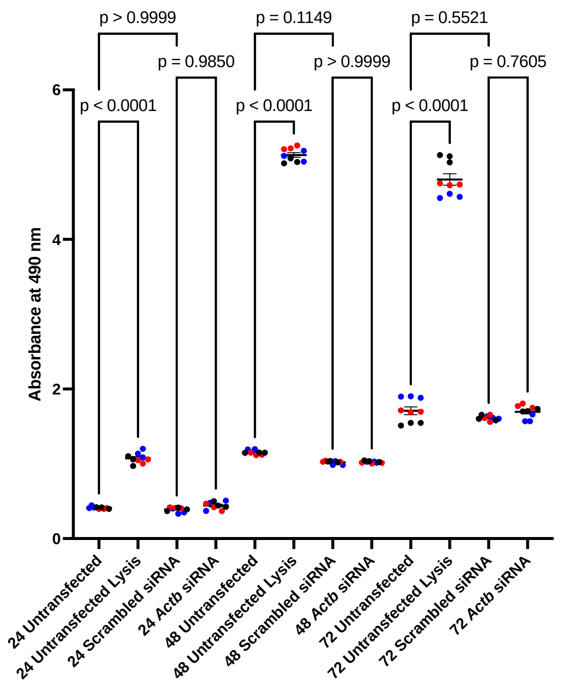

**Supplementary Figure S16. PCR validation of the genotypes of *Actb*$^{+/-}$ mice.**

The **top image** shows the schematic for primer locations for the validation of Floxed *Actb* alleles and a representative gel. The **bottom image** shows the schematic for primer locations for the validation of K14 Cre and the *Actb* deletion alleles and a representative gel. Internal control was *Myogenin*. (1) and (2) represent progenies of female *K14-Cre* crossed with male *Actb*$^{flox}$ resulting in *Actb*$^{del}$. (3) represents a progeny of female wild crossed with male *Actb*$^{flox}$ resulting in an *Actb*$^{flox}$ pup.

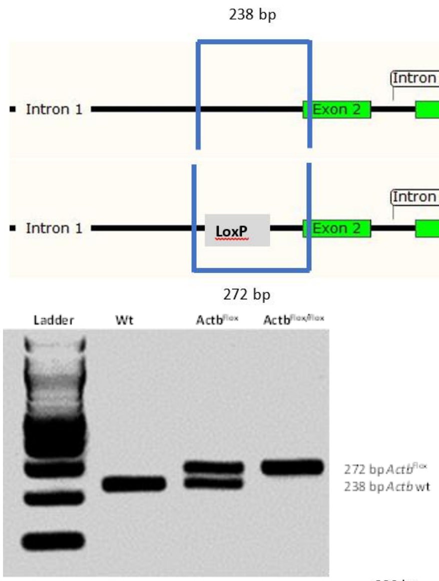

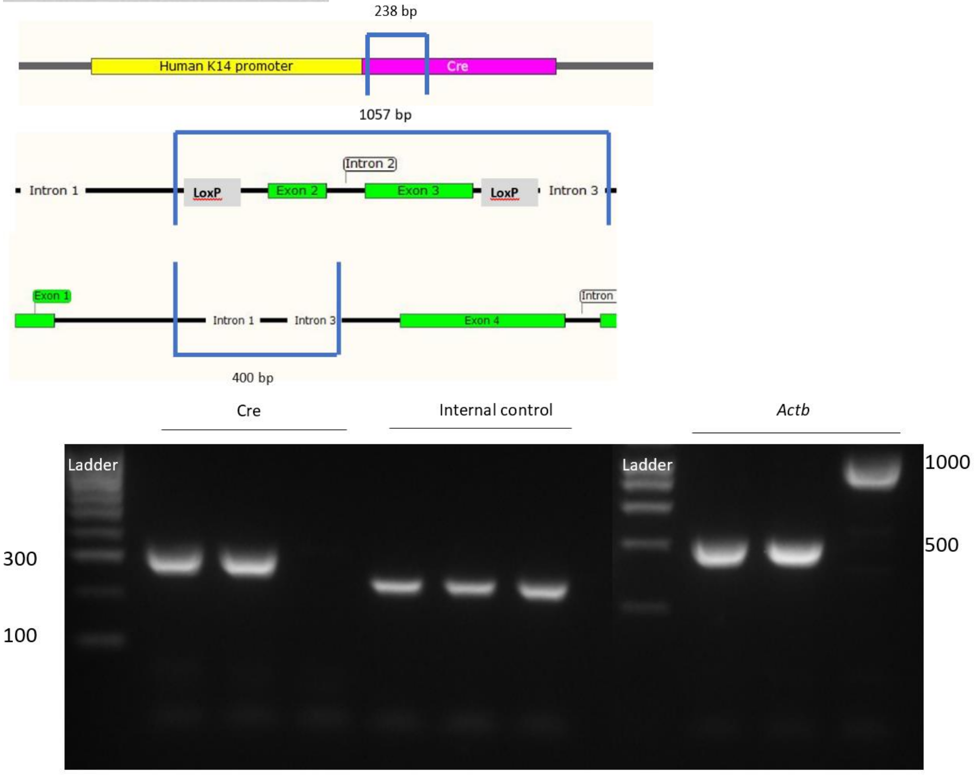

(1) (2) (3) (1) (2) (3) (1) (2)
(3)

**Supplementary Figure S17. Characterisation of the *Actb*$^{+/-}$ D0 pups.**
The thick horizontal line for each measurement group represents mean and the shorter thin horizontal lines represent the SEM. The statistical comparison was done via an individual t-test for each data set investigated and the two tailed p-values are provided for each comparison. N=36 *Actb*$^{+/+}$ and 14 *Actb*$^{+/-}$.

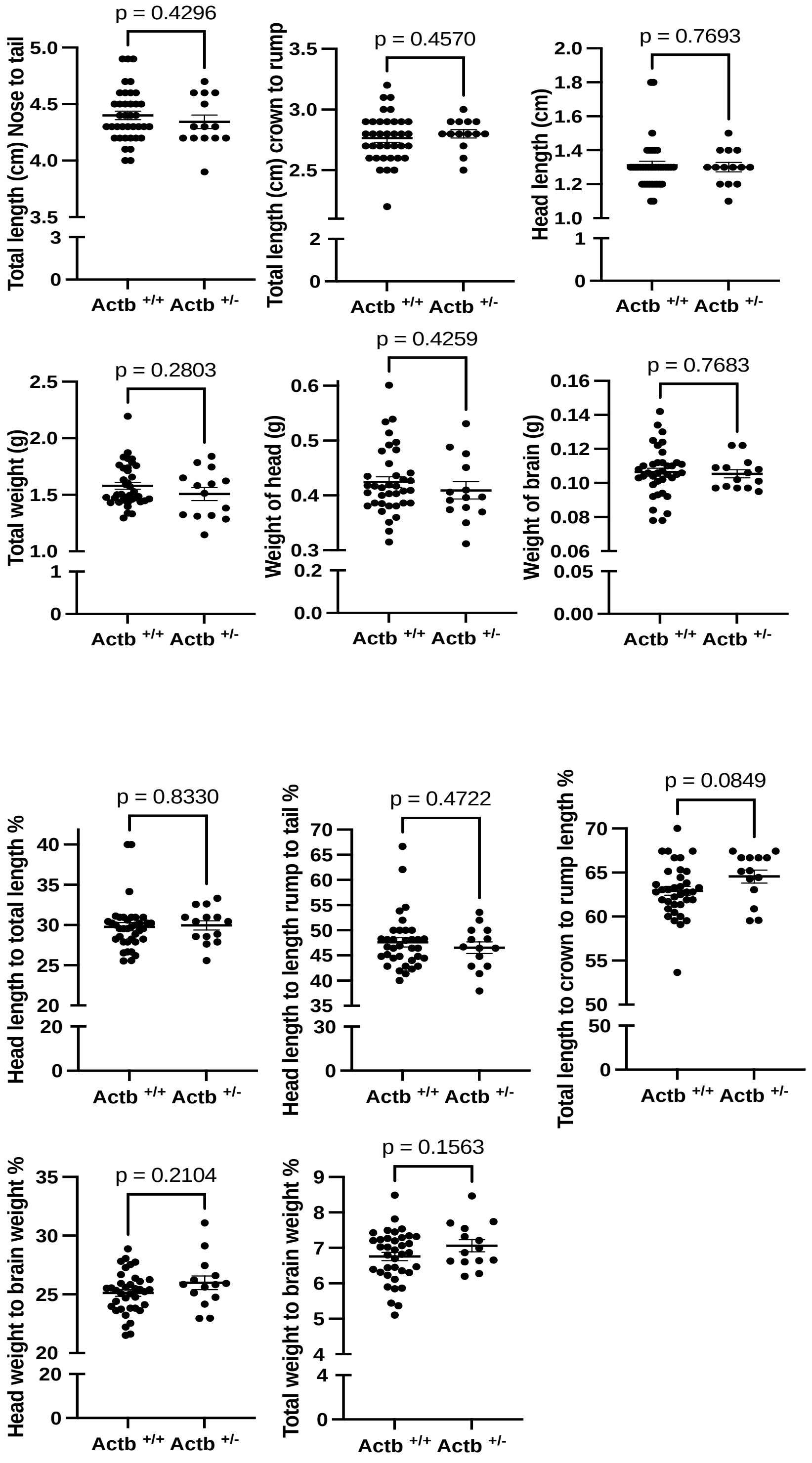

**Supplementary Figure S18. PCA analysis of the D0 male mice brain RNAseq**

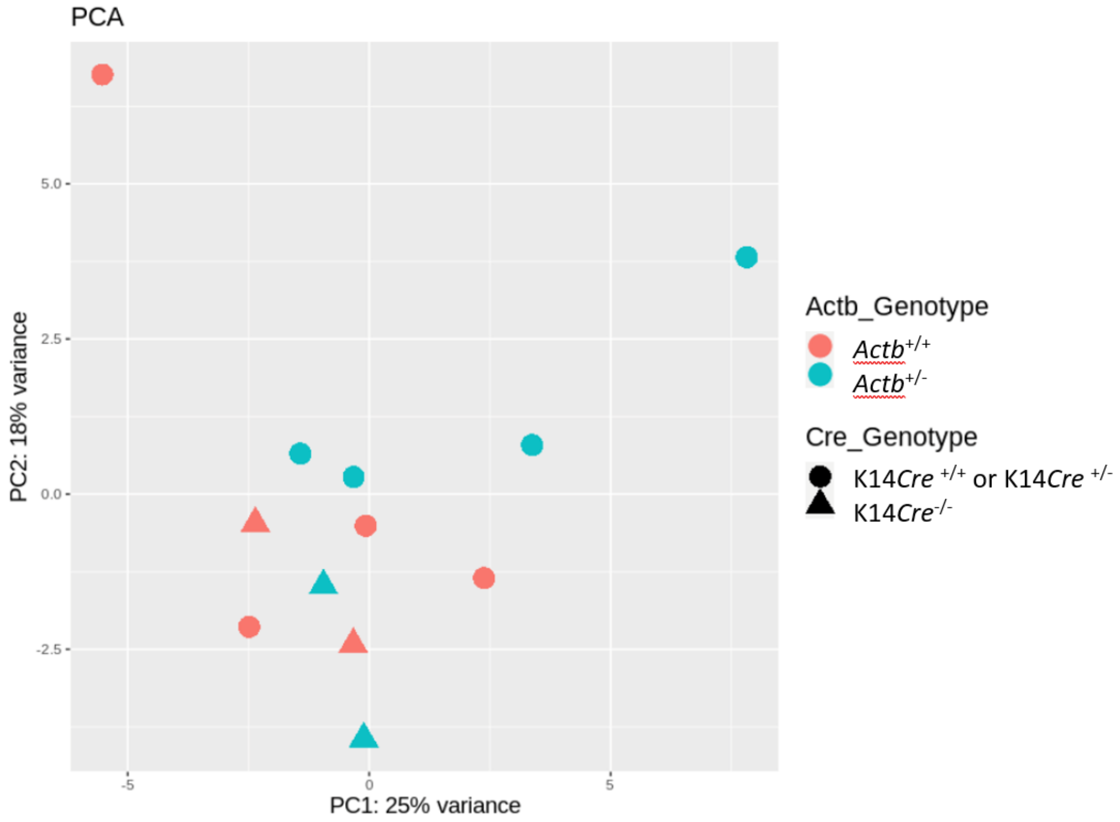

**Supplementary Figure S19. qPCR analysis of the D0 male mice brain RNAseq**

qPCR of male (**top panels**) and female (**bottom panels**) on RNA extracted from D0 neonates. Fold change expression of the genes investigated was calculated with the ΔΔCT method against the housekeeping gene *Eef2* run in triplicate. The thick horizontal line for each experimental group represents mean and the shorter thin horizontal lines represent the SEM. The statistical comparison was done via an individual t-test for each gene investigated and the two tailed p-values are provided for each comparison.

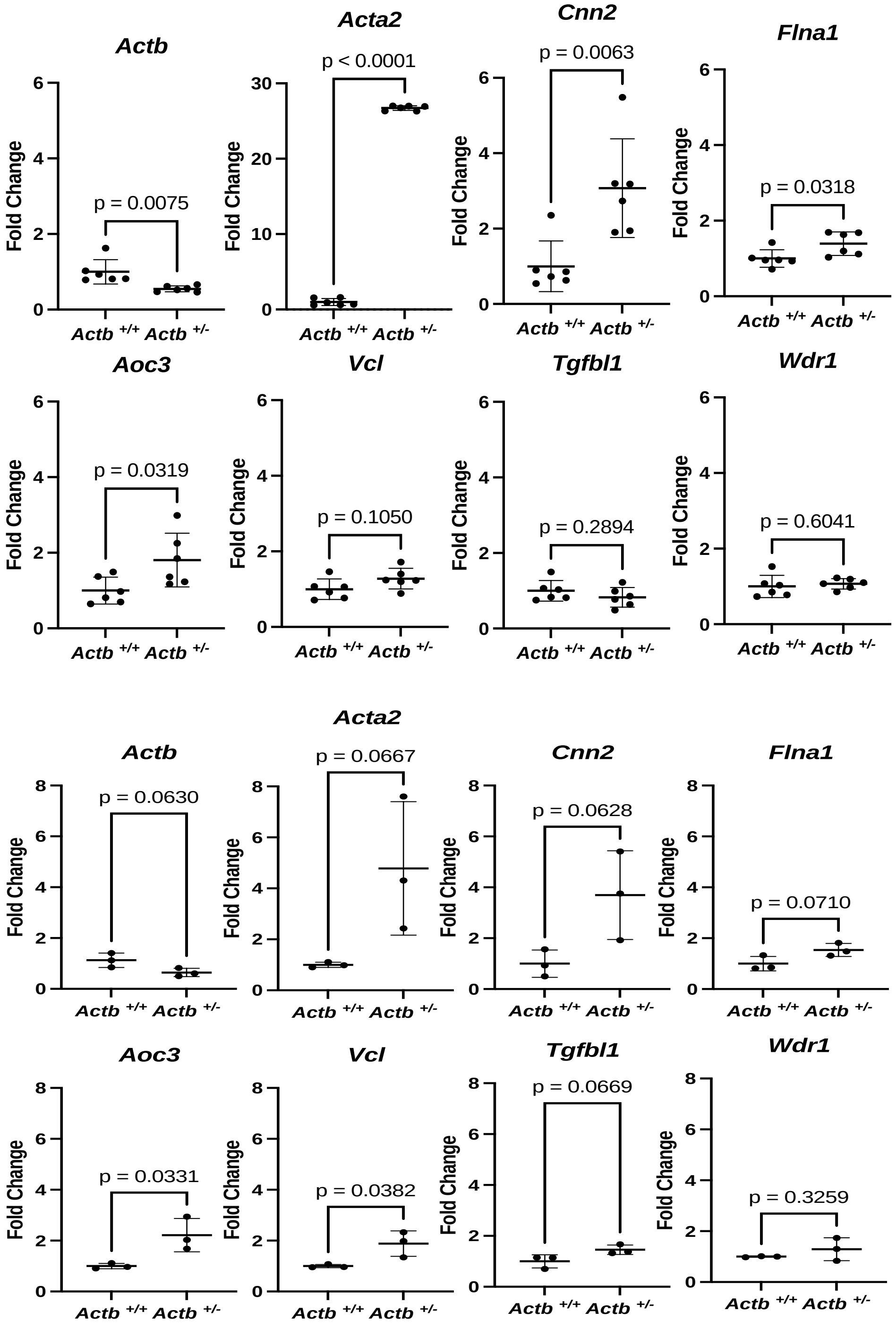

**Supplementary Figure S20. Expression of the neuronal and proliferation markers in the 3D spheroids.**

(A) Immunohistochemical staining of the neuronal spheroid cultures at the end point of the migration assay from the control iPSCs and two BWCFF iPSC lines for Ki-67 (green) and (B) SOX2 (magenta) and TUJ1 (yellow), scale bar 500 µm

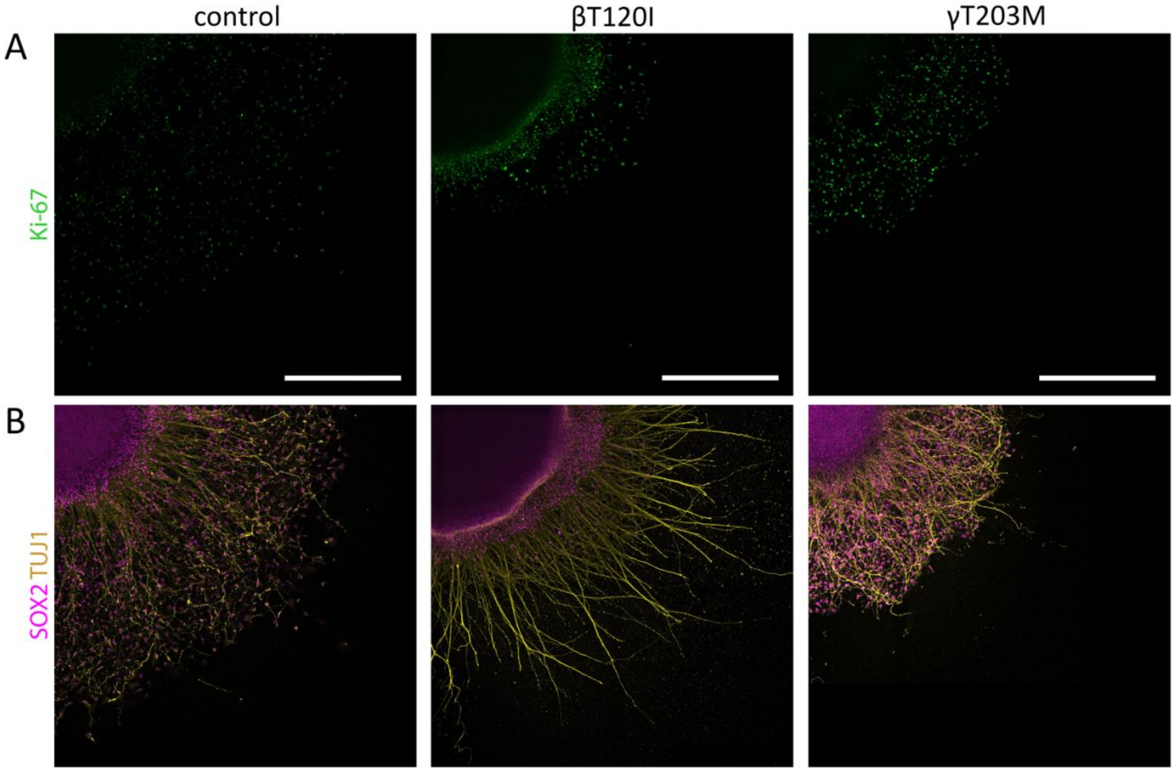

***Supplementary Figure S21. MRI images with cortical malformations typical for BWCFF***
**(A)** T2 weighted axial image demonstrating fronto-temporal pachygyria and prominent perivascular spaces; **(B)** T1 weighted axial image shows anterio predominant pachygyria and a thin band heterotopia in the occipital lobes with prominent perivascular spaces; **(C)** T2 weighted coronal image with the bilateral single periventricular nodules; scale bar 1cm.

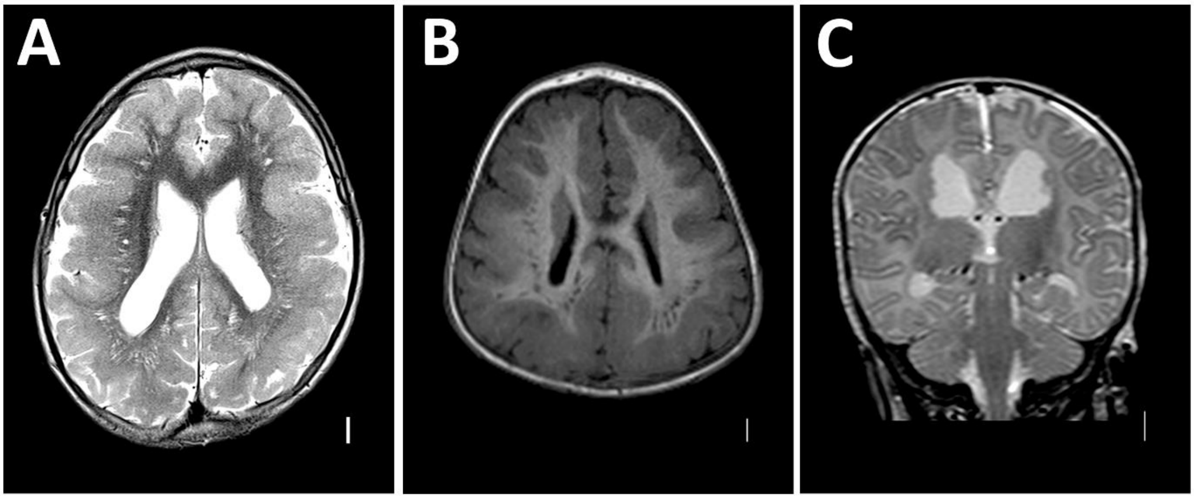

**Supplementary Figure S22. Genomic editing to introduce a BWCFF mutation T120I in *Actb***

**(A)** Guide plasmid vector expressing all elements of CRISPR/Cas9 complex containing the I104-sgRNA1. **(B)** Results of the guide efficiency testing with the wild type *Actb* fragment amplified with the primers I014.6 and I014.7, expected size of the amplicon is 539 bp (lane 1) Lane 2 demonstrates the same amplicon incubated with the guide 81-based RNP complex (expected sized 256/283 bp) as well as the guide 88-based RNP complex (lane 3, expected sizes 269/270 bp) **(C-D)** Example of the initial screening of targeting *Actb* in the ES cells: **(C)** shows PCR with primers I014.7 and I014.9 (mutation-specific primer), positive samples have a mutant allele (D) the same samples as in C amplified with a primer pair I014.6 and I014.7, as above, and cut with restriction enzyme SmaI. SmaI cuts the wild type allele (black arrows), but the restriction site is not present on the homology donor template I014.8. Red arrows indicate samples without an intact wild type allele that cannot be used in the following experiments. **(E-F)** DNA sequencing chromatograms of the selected ES clones, boxes show the end of exon 3.

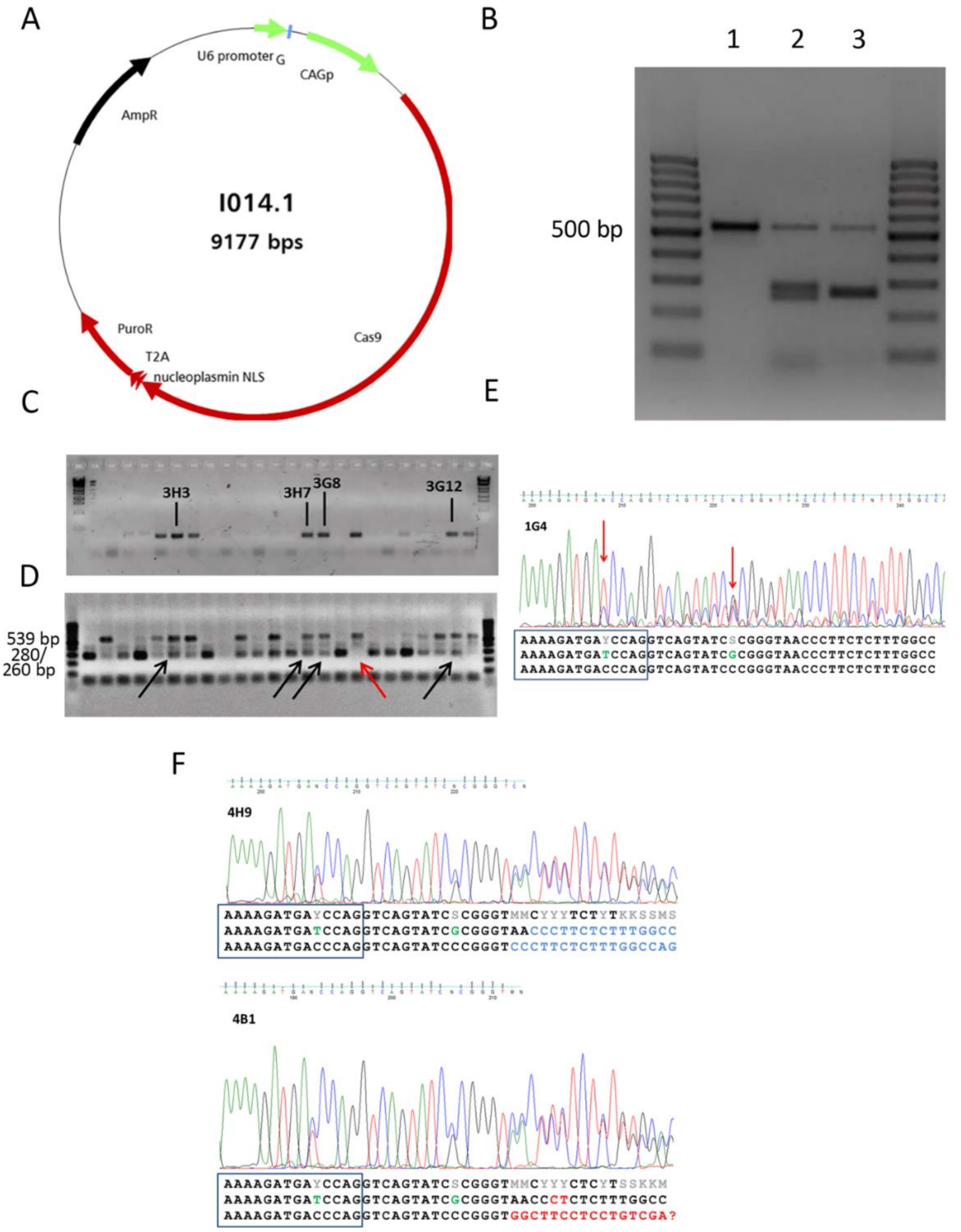

**Supplementary Figure S23. Genomic editing to introduce a BWCFF mutation T120I in *Actg1***

**(A)** Guide plasmid vector expressing all elements of CRISPR/Cas9 complex containing the I104-sgRNA1. **(B)** Results of the guide efficiency testing with the wild type *Actg1* fragment amplified with the primers I018.6 and I018.7, expected size of the amplicon is 609 bp (lane 1) Lane 2 demonstrates the same amplicon incubated with the guide 1 based RNP complex (expected sized 351/258 bp) as well as the guide 2 based RNP complex (lane 3, expected sizes 363/246 bp). **(C)** Example of the initial screening of targeting *Actg1* in the ES cells, where the target *Actg1* region was amplified with I018.6/7 primers and digested with HindIII, expecting a 356 bp and a 253 bp cleavage products; HindIII restriction site was introduced by homology repair template; three candidate clones are indicated with numbers; blue arrow points out a weak signal in the cleavage band and red arrows show the absence of the wild type allele. Only heterozygous clones were picked up for the injections. **(D)** DNA sequencing chromatograms of the selected ES clones, black letters correspond to the coding exonic and grey to the intronic sequence, targeted nucleotide substitution is highlighted in blue and HindIII restriction site in red; all clones show an intronic deletions on the second allele. **(E)** Founder animals from clone 1A4, I018.11 a female 30% chimera with tooth defect, I018.12 30% male chimera also with slant teeth.

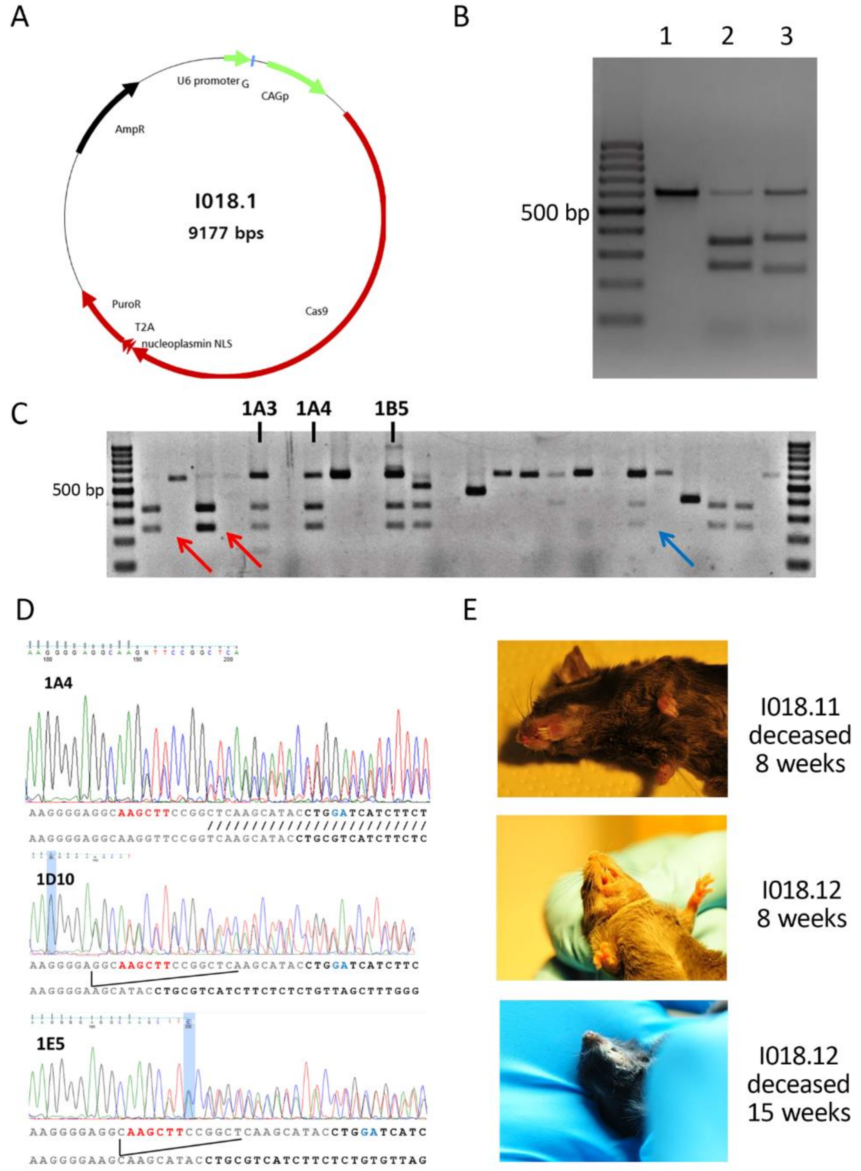

**Supplementary Figure S24. F1 screening for BWCFF mutation in *Actg1***

**(A)** Screening of 5 pups from the clone 1D10 with the HindIII digestion of the PCR product amplified with the primers I018.6/7 demonstrated the HindIII-inserted genotype in two samples (I018.1003 and I018.1004) **(B)** Sequencing of the HindIII digested samples demonstrate the heterozygous target substitution along with the HindIII restriction site (red bars) and an upstream frameshift mutation (read arrow).

A

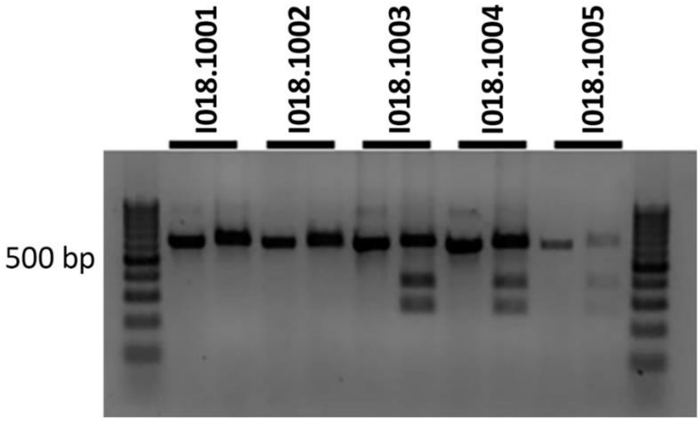

B

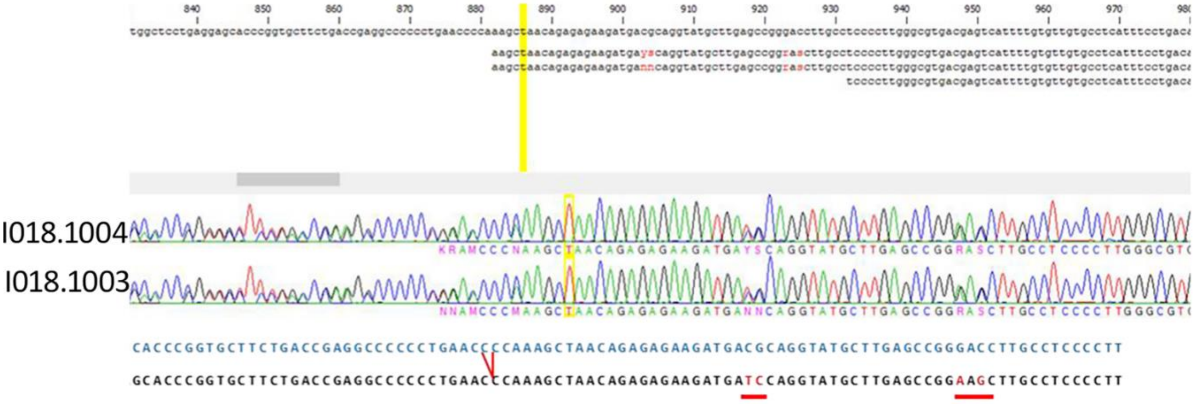

**Supplementary Table 4. List of antibodies**

Primary antibodies

| Target | Specificity | Company | Catalog no. # | RRID | Dilution factor |
|---|---|---|---|---|---|
| β-CYA | Mouse monoclonal $IgG_1$ clone 4C2 | bio-rad | CMCA5775GA | AB_2571580 | 1:50 (IF) 1:750 (WB) |
| g-CYA | Mouse monoclonal $IgG_{2b}$ clone 2A3 | bio-rad | MCA5776GA | AB_2571583 | 1:100 (IF) 1:7500 (WB) |
| Ki-67 | Rabbit polyclonal | abcam | ab15580 | AB_443209 | 1:300 (IF) |
| Tuj1 | Mouse monoclonal | BioLegend | #801201 | AB_2313773 | 1:300 (IF) |
| Pan-Actin | Mouse monoclonal | Novus Biologicals | NB600-535 | AB_2222881 | 1:200 (IF) 1:2500 (WB) |
| SOX2 | Goat polyclonal | R+D Systems | AF2018 | AB_355110 | 1:300 (IF) |
| DAPI | | Roche | 10236276001 | | 1:1000 |
| IRDye® 800CW | | Li-COR | 926-32210 | | 1:15000 |

Secondary antibodies

| Host/ Target | Isotype | Conjugate | Company | Catalog no. # | RRID | Dilution factor |
|---|---|---|---|---|---|---|
| Goat anti Mouse | Mouse IgG, Fcg Subclass 1 Specific | AlexaFluor 488 | Jackson Immunoresearch | 115-545-205 | AB_2338854 | 1:200 |
| Goat anti Mouse | Mouse IgG, Fcg Subclass 2b Specific | CY5 | Jackson Immunoresearch | 115-175-207 | AB_2338717 | 1:50 |
| Donkey anti Mouse | Donkey IgG | AlexaFluor 488 | Thermo Fisher | A-21202 | AB_141607 | 1:500 |
| Donkey anti Rabbit | Donkey IgG | AlexaFluor 555 | Thermo Fisher | A-31572 | AB_162543 | 1:500 |
| Donkey anti Goat | Donkey IgG | AlexaFluor 647 | Thermo Fisher | A-21447 | AB_2535864 | 1:500 |
| Donkey anti Rat | Donkey IgG | AlexaFluor 488 | Thermo Fisher | A-21208 | AB_2535794 | 1:500 |

**NMA clinical consortium**

| Name | Affiliation 1 | Affiliation 2 |
|---|---|---|
| Andrea Accogli | Department of Specialized Medicine, Division of Medical Genetics, McGill University Health Centre, Montreal, Canada | Department of Human Genetics, McGill University, Montreal, Canada |
| Maria Albers | Department of Genetics, University Medical Center Utrecht, Utrecht, Netherlands | |
| Fowzan Alkuraya | Department of Genetics, King Faisal Specialist Hospital and Research Center, Riyadh, Saudi Arabia | |
| Neophytos Apeshiotis | Praxis für Genetik, Eckert-Str. 12, Braunschweig, Germany | |
| Diana Baralle | Faculty of Medicine, University of Southampton, University of Southampton, Southampton, United Kingdom | |
| Carmen Barba | Neuroscience Department, Meyer Children's Hospital IRCCS, viale Pieraccini 24, 50139, Florence, Italy | Department of NEUROFARBA, University of Florence, viale Pieraccini 6, 50139, Florence, Italy |
| Allan Bayat | Department of Epilepsy Genetics and Personalized Medicine, Danish Epilepsy Centre, Dianalund, Denmark | Department of Clinical Genetics, Copenhagen University Hospital, Rigshospitalet, Copenhagen, Denmark |
| Andreas Benneche | Department of Medical Genetics, Haukeland University Hospital, Bergen, Norway | |
| Laura Bernardini | Medical Genetics Unit, IRCCS Casa Sollievo della Sofferenza Foundation, San Giovanni Rotondo (FG), Italy | |
| Saskia Biskup | Zentrum für Humangenetik Tübingen, Tübingen, Germany | |
| Nina Bögershausen | Institute of Human Genetics, University Medical Center Göttingen, Göttingen, Germany | |
| Knut Brockmann | Department of Pediatrics and Adolescent Medicine, University Medical Center Göttingen, Göttingen, Germany | |
| Nicola Brunetti-Pierri | Telethon Institute of Genetics and Medicine (TIGEM), Pozzuoli, Naples, Italy | Department of Translational Medicine, Federico II University, Naples, Italy |
| Peter Burfeind | Institute of Human Genetics, University Medical Center Göttingen, Göttingen, Germany | |
| Ruben Cabanillas | Cabanillas Precision Consulting, Zurich, Switzerland | Translational Medicine, T-Therapeutics, Cambridge, United Kingdom |

| | | |
|---|---|---|
| Patricia Corriols-Noval | Department of Otorhinolaryngology, Hospital Universitario Marqués de Valdecilla, Santander, Spain | |
| Elke de Boer | Department of Human Genetics, Radboudumc, 6500 HB, Nijmegen, Netherlands | |
| Iris de Lange | Department of Genetics, University Medical Center Utrecht, Utrecht, Netherlands | |
| Charulata Deshpande | Manchester Centre for Genomic Medicine, St Mary's Hospital, Manchester University NHS Foundation Trust, Manchester, United Kingdom | |
| Marta Diñeiro | Instituto de Medicina Oncológica y Molecular de Asturias (IMOMA), Oviedo, Spain | |
| Emily Doherty | Carilion Clinic Children's Hospital, Roanoke, United States | |
| Julia Doll | Institut für Humangenetik, Biozentrum, Universität Würzburg, Würzburg, Germany | |
| Sofia Douzgou | Department of Medical Genetics, Haukeland University Hospital, Bergen, Norway | |
| Tracy Dudding-Byth | University of Newcastle, The NSW Genetics of Learning Disability Newcastle, Newcastle, Australia | |
| Nadja Ehmke | Institute of Medical Genetics and Human Genetics, Charité-Universitätsmedizin Berlin, Corporate member of Freie Universität Berlin and Humboldt-Universität zu Berlin, Berlin, Germany | |
| Katherine Fawcett | MRC Computational Genomics Analysis and Training Programme (CGAT), MRC Centre for Computational Biology, MRC Weatherall Institute of Molecular Medicine, John Radcliffe Hospital, Oxford, United Kingdom | Department of Population Health Sciences, University of Leicester, LE1 7RH, Leicester, United Kingdom |
| Carlos R. Ferreira | National Human Genome Research Institute, National Institutes of Health, 20892, Bethesda, United States | |
| Jan Fischer | Institute for Clinical Genetics, Medical Faculty and University Hospital Carl Gustav Carus, TUD Dresden University of Technology, Fetscherstrabe 78, 01311, Dresden, Germany | |
| Joel Fluss | Pediatric Neurology Unit, Paediatrics Subspecialties Service, Geneva Children's Hospital, Geneva, Switzerland | |
| Rocío González-Aguado | Department of Otorhinolaryngology, Hospital Universitario Marqués de Valdecilla, Santander, Spain | |
| Luitgard Graul-Neumann | Institute of Medical Genetics and Human Genetics, Charité-Universitätsmedizin Berlin, Corporate member of Freie Universität Berlin | |

| | | |
|---|---|---|
| | and Humboldt-Universität zu Berlin, Berlin, Germany | |
| Andrew Green | UCD School of Medicine and Medical Science, Children’s Health Ireland (CHI) at Crumlin, Dublin, Ireland | |
| Renzo Guerrini | Neuroscience Department, Meyer Children's Hospital IRCCS, viale Pieraccini 24, 50139, Florence, Italy | Department of NEUROFARBA, University of Florence, viale Pieraccini 6, 50139, Florence, Italy |
| Asya Gusina | Laboratory of Cytogenetic, Molecular Genetic and Morphological Studies, National Research and Applied Medicine Centre 'Mother and Child", Minsk, Belarus | |
| Ute Hehr | Center for Human Genetics, Regensburg, Germany | |
| Maja Hempel | Institute of Human Genetics, Heidelberg University, Heidelberg, Germany | |
| Michaela AH Hofrichter | Institut für Humangenetik, Biozentrum, Universität Würzburg, Würzburg, Germany | |
| Ivan Ivanovski | Medical Genetics Unit, Azienda USL-IRCCS di Reggio Emilia, Reggio Emilia, Italy | Institute of Medical Genetics, University of Zurich, Zürich, Switzerland |
| Wibke G. Janzarik | Department of Neuropediatrics and Muscle Disorders, Center for Pediatrics and Adolescent Medicine, Medical Center, Faculty of Medicine, University of Freiburg, Freiburg, Germany | |
| Diana Johnson | Department of Medical Genetics, National Health Service, NHS, Leeds, United Kingdom | |
| Marieke Joosten | Department of Clinical Genetics, Erasmus MC, Rotterdam, Netherlands | |
| Silke Kaulfub | Institute of Human Genetics, University Medical Center Göttingen, Göttingen, Germany | |
| Hyun Jung Kim | Department of Pediatrics, Eulji General Hospital, College of Medicine, Eulji University, Seoul, Republic of Korea | |
| Tjitske Kleefstra | Department of Human Genetics, Radboudumc, 6500 HB, Nijmegen, Netherlands | Donders Institute for Brain, Cognition and Behaviour, Radboud University, 6500 GL, Nijmegen, Netherlands |
| Eva Klopocki | Institut für Humangenetik, Biozentrum, Universität Würzburg, Würzburg, Germany | |
| Karla Krause | Institute for Clinical Genetics, Medical Faculty and University Hospital Carl Gustav Carus, TUD Dresden University of Technology, Fetscherstrabe 77, 01310, Dresden, Germany | |

| | | |
|---|---|---|
| Alma Kuechler | Institute of Human Genetics, University Hospital Essen, University Duisburg-Essen, 45122, Essen, Germany | |
| Maria Kuzyakova | Institute of Human Genetics, University Medical Center Göttingen, Göttingen, Germany | |
| Martin W. Laass | Department of Pediatrics, Medizinische Fakultät Carl Gustav Carus, TUD Dresden University of Technology, Dresden, Germany | |
| Augusta Lachmeijer | Department of Genetics, University Medical Center Utrecht, Utrecht, Netherlands | |
| Wayne Lam | South East of Scotland Clinical Genetics Service, Edinburgh, United Kingdom | |
| Cha Gon Lee | Department of Pediatrics, Nowon Eulji Medical Center, Eulji University School of Medicine, Seoul, Republic of Korea | |
| Yun Li | Institute of Human Genetics, University Medical Center Göttingen, Göttingen, Germany | |
| Vanesa López-González | Sección de Genética Médica, Servicio de Pediatría, Hospital Clínico Universitario Virgen de la Arrixaca, Murcia, Spain | |
| Karen Low | Department of Clinical Genetics, University Hospitals Bristol NHS Foundation Trust, Bristol, United Kingdom | Centre for Academic Child Health, Bristol Medical School, University of Bristol, Bristol, United Kingdom, |
| Michael Lyons | Greenwood Genetic Center, Greenwood, United States | |
| Carlo Marcelis | Department of Clinical Genetics, Radboud University Medical Center, Nijmegen, Netherlands | |
| Francisco Martinez-Castellano | Unit of Genetics, Hospital Universitari i Politècnic La Fe. Valencia, Valencia, Spain | Genomics Unit, Instituto de Investigación Sanitaria La Fe, 46026, Valencia, Spain |
| Maarten Massink | Department of Genetics, University Medical Center Utrecht, Utrecht, Netherlands | |
| Kay Metcalfe | Manchester Centre for Genomic Medicine, St Mary's Hospital, Manchester University NHS Foundation Trust, Manchester, United Kingdom | |
| Donatella Milani | Fondazione IRCCS Ca' Granda Ospedale Maggiore Policlinico, Milan, Italy | |
| Shahida Moosa | Division of Molecular Biology and Human Genetics, Faculty of Medicine and Health Sciences, Stellenbosch University, Tygerberg, South Africa | Medical Genetics, Tygerberg Hospital, South Africa |
| Manuela Morleo | Telethon Institute of Genetics and Medicine (TIGEM), Pozzuoli, Naples, Italy | Department of Precision Medicine, University of Campania “Luigi Vanvitelli”, Naples, Italy |

| | | |
|---|---|---|
| Teresa Neuhann | Institute of Human Genetics, University Medical Center Göttingen, Göttingen, Germany | |
| Thomas Neumann | Mitteldeutscher Praxisverbund Humangenetik, Halle, Germany | |
| Huu Nguyen | Department of Human Genetics, Ruhr-University Bochum, Bochum, Germany | |
| Vincenzo Nigro | Telethon Institute of Genetics and Medicine (TIGEM), Pozzuoli, Naples, Italy | Department of Precision Medicine, University of Campania “Luigi Vanvitelli”, Naples, Italy |
| Nuha Nimeri | Women’s Wellness and Research Center, NICU, Hamad Medical Corporation, Doha, Qatar | |
| Ewa Obersztyn | Department of Medical Genetics, Institute of Mother and Child, Warsaw, Poland | |
| Anne O'Donnell | Division of Genetics and Genomics, Boston Children's Hospital, Boston, United States | |
| Carmen Orellana | Unit of Genetics, Hospital Universitari i Politècnic La Fe. Valencia, Valencia, Spain | |
| Estrella Pallas | Department of Otorhinolaryngology, Hospital Álvaro Cunqueiro, Vigo, Spain | |
| Hans-Jürgen Pander | Institute of Clinical Genetics, Klinikum Stuttgart, Stuttgart, Germany | |
| Elena Parrini | Neuroscience Department, Meyer Children's Hospital IRCCS, viale Pieraccini 24, 50139, Florence, Italy | |
| Silke Pauli | Institute of Human Genetics, University Medical Center Göttingen, Göttingen, Germany | |
| Michele Pinelli | Department of Molecular Medicine and Medical Biotechnologies, University Federico II, Naples, Italy | Telethon Institute of Genetics and Medicine (TIGEM), Pozzuoli, Naples, Italy |
| Lina Quteineh | Division of Genetic Medicine, Geneva University Hospitals, Geneva, Switzerland | |
| Julia Rankin | Peninsula Clinical Genetics Service, Royal Devon and Exeter NHS Trust, Exeter, United Kingdom | |
| Monica Rosello | Unit of Genetics, Hospital Universitari i Politècnic La Fe. Valencia, Valencia, Spain | |
| Tamanna Roshan Lal | Genetics and Metabolism, Children's National Hospital, Washington, United States | |
| Vincenzo Salpietro | Department of Neuromuscular Disorders, Queen Square Institute of Neurology, University College London, WC1N 3BG, London, United Kingdom | Department of Biotechnological and Applied Clinical Sciences, University of L'Aquila, 67100, L'Aquila, Italy |
| Jens Schallner | Department of Neuropediatrics, TUD Dresden University of Technology, Dresden, Germany | |
| Gregor Schlüter | PRAENATAL, Nürnberg, Germany | |

| | | |
|---|---|---|
| Julia Schmidt | Institute of Human Genetics, University Medical Center Göttingen, Göttingen, Germany | |
| Mariasavina Severino | Neuroradiology Unit, IRCCS Istituto Giannina Gaslini, Genoa, Italy | |
| Vandana Shashi | Department of Pediatrics, Division of Medical Genetics, Duke University Medical Center, Durham, United States | |
| Corinna Siegel | Institute of Human Genetics, Klinikum rechts der Isar, Technical University of Munich, Munich, Germany | Department of Clinical Genetics, MVZ Martinsried, Munich, |
| Margie Sinnema | Department of Clinical Genetics, Maastricht University Medical Center, Maastricht, Netherlands | |
| Anne Slavotinek | Division of Genetics, Department of Pediatrics, University of California, San Francisco, United States | Division of Human Genetics, Cincinnati Children's Hospital, 3333 Burnet Ave, Cincinnati OH 45229, United States |
| Sarah Smithson | Department of Clinical Genetics, University Hospitals Bristol NHS Foundation Trust, Bristol, United Kingdom | |
| Siddharth Srivastava | Department of Neurology, Boston Children's Hospital, Boston, United States | |
| Maja Svrakic | Northwell Health Department of Otolaryngology, New York, United States | |
| Lindsay Swanson | Department of Neurology, Boston Children's Hospital, Boston, United States | |
| Hannah Thomson | Hunter Genetics, The NSW Genetics of Learning Disability Newcastle, Newcastle, Australia | |
| Eduardo Tizzano Ferrari | Àrea de Genètica Clínica i Molecular, Hospital Vall d'Hebrón, Barcelona, Spain | |
| Annalaura Torella | Telethon Institute of Genetics and Medicine (TIGEM), Pozzuoli, Naples, Italy | Department of Precision Medicine, University of Campania “Luigi Vanvitelli”, Naples, Italy |
| Undiagnosed Diseases Network | | |
| Irene Valenzuela Palafoll | Àrea de Genètica Clínica i Molecular, Hospital Vall d'Hebrón, Barcelona, Spain | |
| Yolande van Bever | Department of Clinical Genetics, ErasmusMC University Medical Center Rotterdam, 3015 GD, Rotterdam, Netherlands | |
| Ellen van Binsbergen | Department of Genetics, University Medical Center Utrecht, Utrecht, Netherlands | |

| | | |
|---|---|---|
| Marjon van Slegtenhorst | Department of Clinical Genetics, ErasmusMC University Medical Center Rotterdam, 3015 GD, Rotterdam, Netherlands | |
| Nienke Verbeek | Department of Genetics, University Medical Center Utrecht, Utrecht, Netherlands | |
| Virginie Verhoeven | Department of Clinical Genetics, ErasmusMC University Medical Center Rotterdam, Rotterdam, Netherlands | |
| Barbara Vona | Institute of Human Genetics, University Medical Center Göttingen, Heinrich-Düker-Weg 12, 37073, Göttingen, Germany | Institute for Auditory Neuroscience and InnerEarLab, University Medical Center Göttingen, Robert-Koch-Str. 40, 37075, Göttingen, Germany |
| Dagmar Wahl | Medical Practice for Genetic Counselling, Center for Human Genetics and Laboratory Diagnostics Martinsried, Augsburg, Germany | |
| Luisa Weiss | Center for Human Genetics, Regensburg, Germany | |
| Gökhan Yigit | Institute of Human Genetics, University Medical Center Göttingen, Göttingen, Germany | DZHK (German Center for Cardiovascular Research), partner site Göttingen, Göttingen, Germany |
| Maha Zaki | Clinical Genetics Department, Human Genetics and Genome Research Institute, National Research Centre, Cairo, Egypt | |
